# Prospective analysis of incident disease among individuals of diverse ancestries using genetic and conventional risk factors

**DOI:** 10.1101/2023.10.23.23297414

**Authors:** Wei Wang, Nicholas Eriksson, Matthew McIntyre, Rafaela Bagur Quetglas, Bertram L. Koelsch, David A. Hinds, Stella Aslibekyan, Adam Auton, Michael V. Holmes, Suyash S. Shringarpure, the 23andMe Research Team

**Author notes:** joint senior authors (Michael V. Holmes, Suyash S. Shringarpure). Correspondence to: Dr Wei Wang, PhD Dr Michael V Holmes, MD PhD Dr Suyash S Shringarpure, PhD.

## Abstract

**Background:** Human genetics provides opportunities for enhancing disease prediction through polygenic risk scores (PRS).

**Method:** We used a dataset from 23andMe (6.77M European, 1.30M Latine, and 0.45M African American individuals). Using cross-sectional data for PRS construction and a prospective cohort for evaluation, we estimated PRS-associated cumulative incidences after one year of follow-up for 12 clinical endpoints.

**Results:** The cumulative incidence of disease at one year was consistently higher among individuals in the top 10% of each PRS. Relative risks (RRs) comparing top to bottom 10% of PRS differed across diseases (e.g. European RR 2.12 for eczema vs 12.53 for T2D). Estimates were similar between Europeans and Latines however were more modest for African Americans (e.g. T2D RR 10.92 for Latines vs. 4.00 for African Americans). Clinical manifestation occurred earlier among those in top vs bottom 10% of polygenic risk: 16yrs for hypertension, and 9.5yrs for T2D. Among participants at elevated conventional risk of CHD or T2D, those in the top 10% PRS had a 10-20 fold higher RR of disease incidence vs those not at conventional risk. Among individuals at high polygenic risk of CHD or T2D, favorable lifestyle characteristics associated with 64-73% lower RR of developing disease over 1-year, with cumulative incidence equivalent to the population average.

**Conclusion:** In an ancestrally-diverse cohort, individuals in the top 10% PRS had higher 1-year disease incidence and earlier age of clinical manifestation. PRS provided risk stratification beyond conventional risk factors. Lifestyle characteristics markedly lowered disease incidence among those at elevated polygenic risk.

## 1 Introduction

Elucidating susceptibility to disease is a cornerstone of preventive medicine^1^. Conventional risk prediction algorithms use phenotypic (i.e. non-genotypically measured) variables to estimate risk of disease^2,3^.

The discovery of large numbers of naturally-occuring single-nucleotide polymorphisms (SNPs) associated with disease from increasingly large GWAS together with improved predictive modeling methodologies have led to the creation of polygenic risk scores (PRS) that can accurately predict disease risk^4–9^.

Recent studies have highlighted both the European-centric bias of GWAS^10–12^, meaning most PRS are derived from populations of predominantly European ancestry, and that predictions from PRS are usually not portable across ancestries^13^. This poses a potential roadblock to the equitable application of PRS to disease prediction strategies^8^.

Here, we construct and evaluate PRS for multiple clinical endpoints among individuals of European, Latine, and African American ancestries. We show that PRS predict disease incidence among ancestrally diverse groups, and adds predictive utility beyond conventional risk factors. Lifestyle factors profoundly attenuated risk from polygenic susceptibility to CHD and T2D. Taken together, our findings illustrate multiple public health opportunities for preventing disease through applications of PRS.

## 2 Methods

### Study participants

Participants in this study were selected from the research participant base of 23andMe, a personal genetics company. Participants provided informed consent and participated in the research online, under a protocol approved by the external AAHRPP-accredited IRB, Ethical & Independent Review Services (E&I Review, now Salus IRB). Participants were included in the analysis on the basis of consent status as checked at the time data analyses were initiated. We divided 23andMe research-consented individuals into two non-overlapping datasets: a discovery dataset and a prospective cohort (**Methods S.1, Figure S1**).

### Study endpoints

We collected information on self-reported disease at baseline and during follow-up for 12 endpoints comprising: CHD, T2D, high blood pressure, anemia, blood clots, hypothyroidism, kidney stones, Crohn’s disease, glaucoma, psoriasis and eczema (**Table 1**).

**Table 1.**
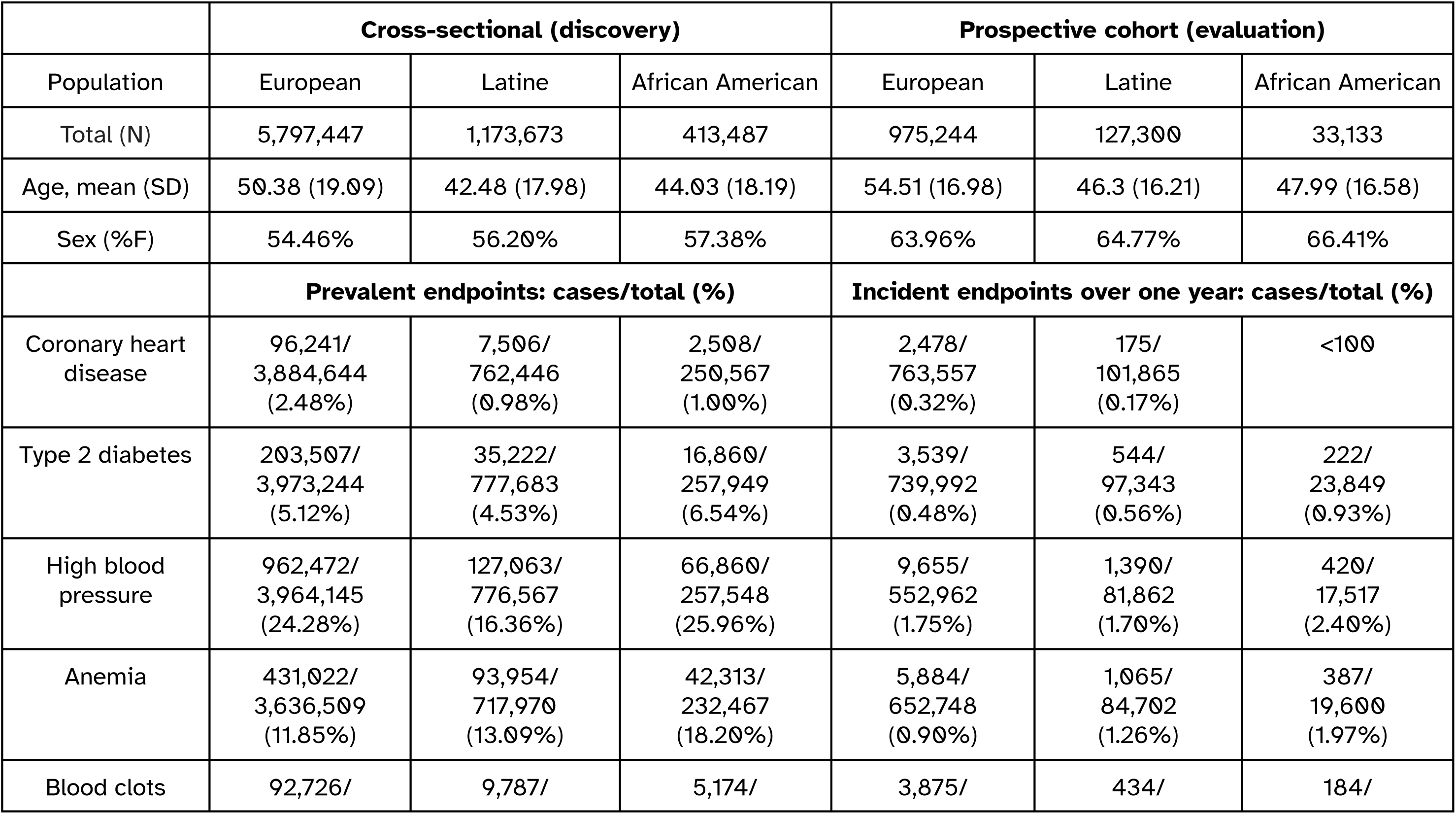
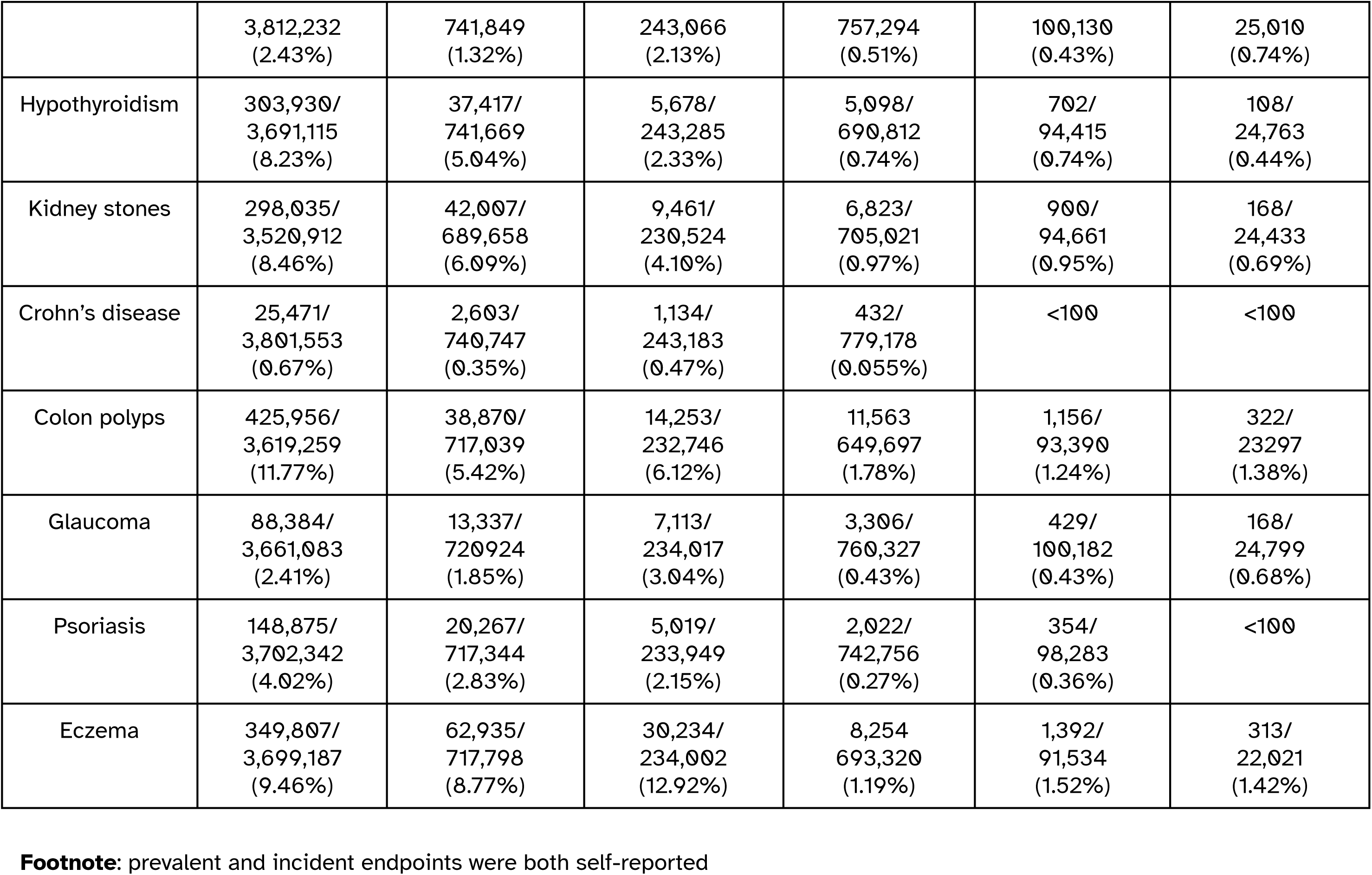
Characteristics of the cross-sectional and prospective cohort datasets from 23andMe used for PRS discovery and evaluation.

### Polygenic risk scores

We conducted GWAS of prevalent disease status in the discovery dataset. GWAS summary statistics were generated using logistic regression assuming an additive model for allelic effects with adjustment for age, sex, principal components (PCs), and genotyping platforms (**Methods S.2**). We conducted GWAS separately in each of three groups defined by genetic ancestry (European, African American, and Latine). We built PRS models by extending the stacked pruning and thresholding (P+T) approach^14^ to multiple populations. We evaluated PRS model performance in a hold-out testing data from the discovery dataset. Further details are provided in the **Methods S.3**.

### Lifestyle risk scores

We used data on 40 lifestyle factors, comprising information on diet, exercise, sleeping, smoking and alcohol consumption. These 40 traits were combined in a Lifestyle Risk Score (LRS), trained separately for CHD and T2D, using a tree-based predictive model (XGboost^15^). We built the lifestyle risk prediction model using self-reported lifestyle variables reported at baseline in the discovery dataset and applied this fitted model to self-reported lifestyle at baseline in individuals in the prospective cohort. Further details are provided in the **Methods S.4**.

### Statistical analysis

We estimated the cumulative incidence of clinical events among participants aged 15-85 yrs in the prospective cohort. Cumulative incidence was calculated as the proportion of individuals that reported incident events as of a given age among all individuals that responded to follow-up questions. The 95% confidence interval (CI) of the cumulative incidence was estimated using bootstrapping. We conducted analyses separately by ancestral groups in order to explore the comparative performance of PRS across ethnically diverse populations. We compared the median age of diagnosis by PRS group. We explored the extent to which PRS added risk stratification among individuals considered to be at conventionally high risk of disease (e.g. individuals at conventional risk of CHD were defined as those who self-reported being hypertensive or taking lipid lowering therapy). Finally, we quantified the cumulative disease incidence among those with high PRS and strata of LRS and compared these individuals to the average incidence of disease in the population (irrespective of PRS or LRS).

In order to ensure we had sufficient data to reliably quantify risk by strata of PRS, we restricted analysis to endpoints for which there were >100 incident events within each ancestral group during the first year of follow-up. We also explored the relationship of PRS with age at self-reported disease onset for prevalent cases.

In this manuscript we abbreviate Latine associated genetic ancestry to Latine ancestry (**Methods S.2**), with similar abbreviations applied to the terms African American ancestry and European ancestry.

## 3 Results

### Characteristics of individuals in the cross-sectional dataset and prospective cohort

The discovery cross-sectional dataset consisted of 7,384,607 individuals and the prospective cohort 1,135,677 individuals. Compared to individuals in the cross-sectional dataset, those in the prospective cohort tended to be older and a greater proportion of them were women (**Table 1**). Among the 7.4M individuals in the cross-sectional dataset, 78.5% were classified as European, 15.9% were classified as Latine, and 5.6% were classified as African American. Conversely, among the 1.1M individuals in the prospective cohort, the corresponding non-European proportions were lower at 11.2% and 2.9%, respectively. In both cross-sectional and prospective cohorts, individuals of Latine ancestry tended to be younger than individuals of African American ancestry, with individuals of European ancestry being the oldest.

The proportion of prevalent cases in the discovery dataset and the number of incident cases during the first year of follow-up in the prospective cohort are reported in **Table 1**. There were 106,255 prevalent and 2,653 incident CHD cases and 255,589 prevalent and 4,305 incident T2D cases.

### Performance of PRS in cross-sectional discovery dataset

PRS varied in predictive performance (**Table S3, Figures S6-S8**): for example, the median of AUC for PRS across the clinical endpoints was 0.65 in the European population, with the worst performance being 0.58 for colon polyps and the best being 0.71 for T2D. The median AUC of clinical endpoints across the three ancestral groups was similar for Europeans (0.65) and Latines (0.63) and lower at 0.58 for African Americans.

### Incident CHD and T2D in individuals of European ancestry

Among Europeans in the prospective cohort, the cumulative incidence of CHD was 0.50% higher (95% CI: [0.43%, 0.56%]) at one year among individuals in the top vs bottom 10% of the PRS, corresponding to a risk ratio of 4.61 (95% CI: [3.79, 5.81]). Those in the high PRS group on average developed CHD 6 years earlier than those in the bottom PRS group (**Table S3**).

The cumulative incidence of T2D was 1.09% higher (95% CI: [1.01%, 1.17%]) at one year among individuals in the top vs bottom 10% of the PRS, with a corresponding risk ratio of 12.53 (95% CI: [9.99, 16.43]). Those in the high PRS group developed T2D on average 10 years earlier than those in the bottom PRS group (**Table S3**).

### Other diseases in individuals of European ancestry

For all endpoints, individuals in the top 10% of disease PRS had a higher cumulative incidence during one-year of prospective follow-up (**Figure 1**), with the average age of onset typically also being younger (**Table S3**). Those in the top vs bottom 10% of PRS developed kidney stones 10.5y earlier, and for hypertension it was 16y earlier.

**Figure 1.**
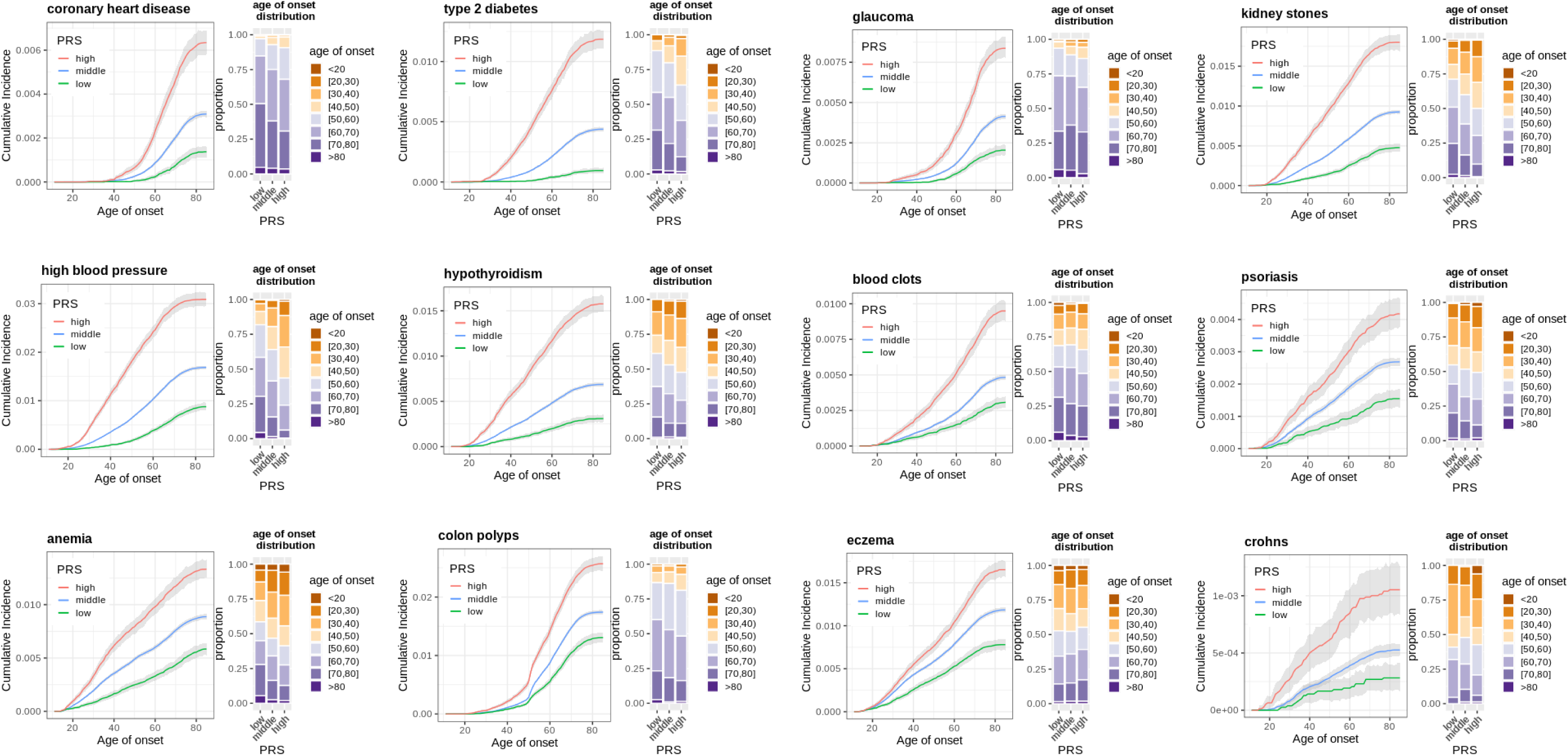
Risk stratification of incident clinical endpoints in Europeans through PRS. Each plot illustrates the cumulative incidence over one year of follow-up among individuals grouped by each disease-specific PRS. Individuals in the top 10% of each PRS are represented by a red line, those in the middle 80% by a blue line and those in the bottom 10% by a green line. The grey shading around each line indicates the 95% CI from bootstrapping. The age of onset distribution for each of the three PRS-defined strata is displayed to the right of each cumulative incidence plot.

### Performance of PRS in individuals of Latine and African American ancestry

**Figure 2** illustrates risk stratification of PRS in Latines and African Americans. The incidence of diseases differed by ancestry e.g. T2D incidence was higher in Latine and African American individuals as compared to Europeans and anemia incidence was up to 2-fold higher in African Americans vs Europeans. **Table S3** and **Figure S9** displays the RR comparing top and bottom 10% of the PRS for multiple diseases among the three ancestrally-defined groups in the prospective dataset (for incident cases) as well as a held-out portion of the discovery dataset (for prevalent cases). There were similar estimates between Europeans and Latines, while RRs tended to be closer to the null for African Americans.

**Figure 2.**
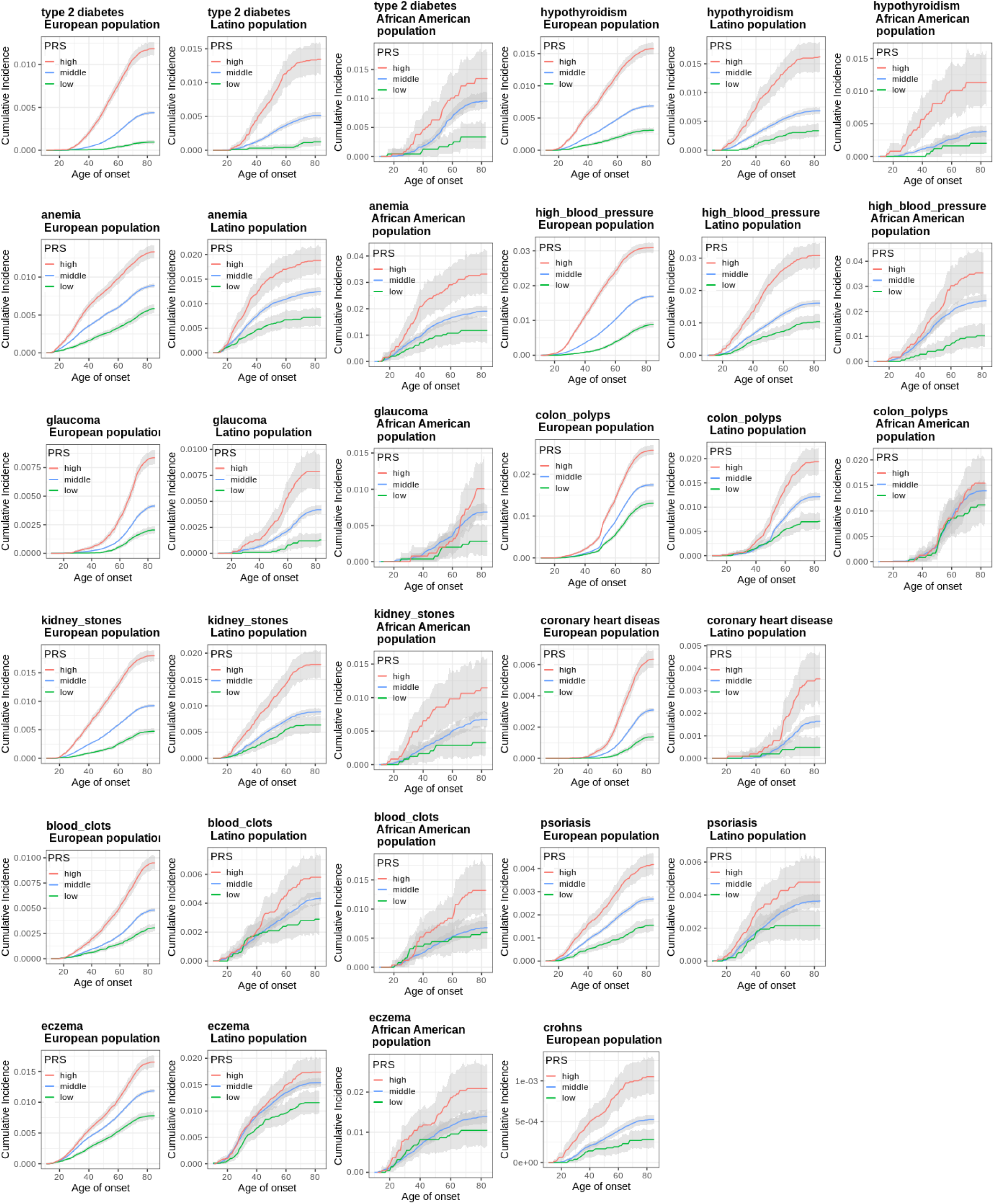
Risk stratification of incident clinical endpoints across different ancestral groups through PRS. Each plot illustrates the cumulative incidence over one year of follow-up among individuals grouped by each disease-specific PRS. Individuals in the top 10% of each PRS are represented by a red line, those in the middle 80% by a blue line and those in the bottom 10% by a green line. The grey shading around each line indicates the 95% CI from bootstrapping.

Owing to comparably fewer cases, the relationship of PRS with risk of incident glaucoma, colon polyps and blood clots in African Americans was less clear than for other endpoints (**Figure 2**). Given the larger number of cases, we identified strong evidence of PRS-derived risk stratification for these conditions using prevalent data (**Figure S2**). For incidence cases where there were <100 events, we explored the relationship of PRS with prevalent cases: for prevalent CHD and psoriasis among African Americans and for Crohn’s among Latine individuals (**Figure S2**), again identifying risk stratification. **Figure S3** provides a visual comparison for 9 endpoints in African Americans where there are data from both incident and prevalent sources, showing the improvement in risk stratification arising from larger numbers of cases.

### Performance of PRS in individuals at high risk of CHD and T2D

Individuals who reported presence of hypertension and/or receiving treatment with lipid lowering therapies at baseline had a cumulative incidence of CHD that was 5.69-fold (95% CI: [5.17, 6.25]) greater than others in the prospective cohort. Among these individuals with conventional cardiovascular risk factors, the cumulative incidence of CHD in the high PRS group was an additional 1.70-fold higher and in individuals with both high conventional and high polygenic risk, the RR was 9.68-fold (95% CI: [8.50, 10.95]) higher than individuals who did not report hypertension/lipid lowering therapy (**Figure 3**).

**Figure 3.**
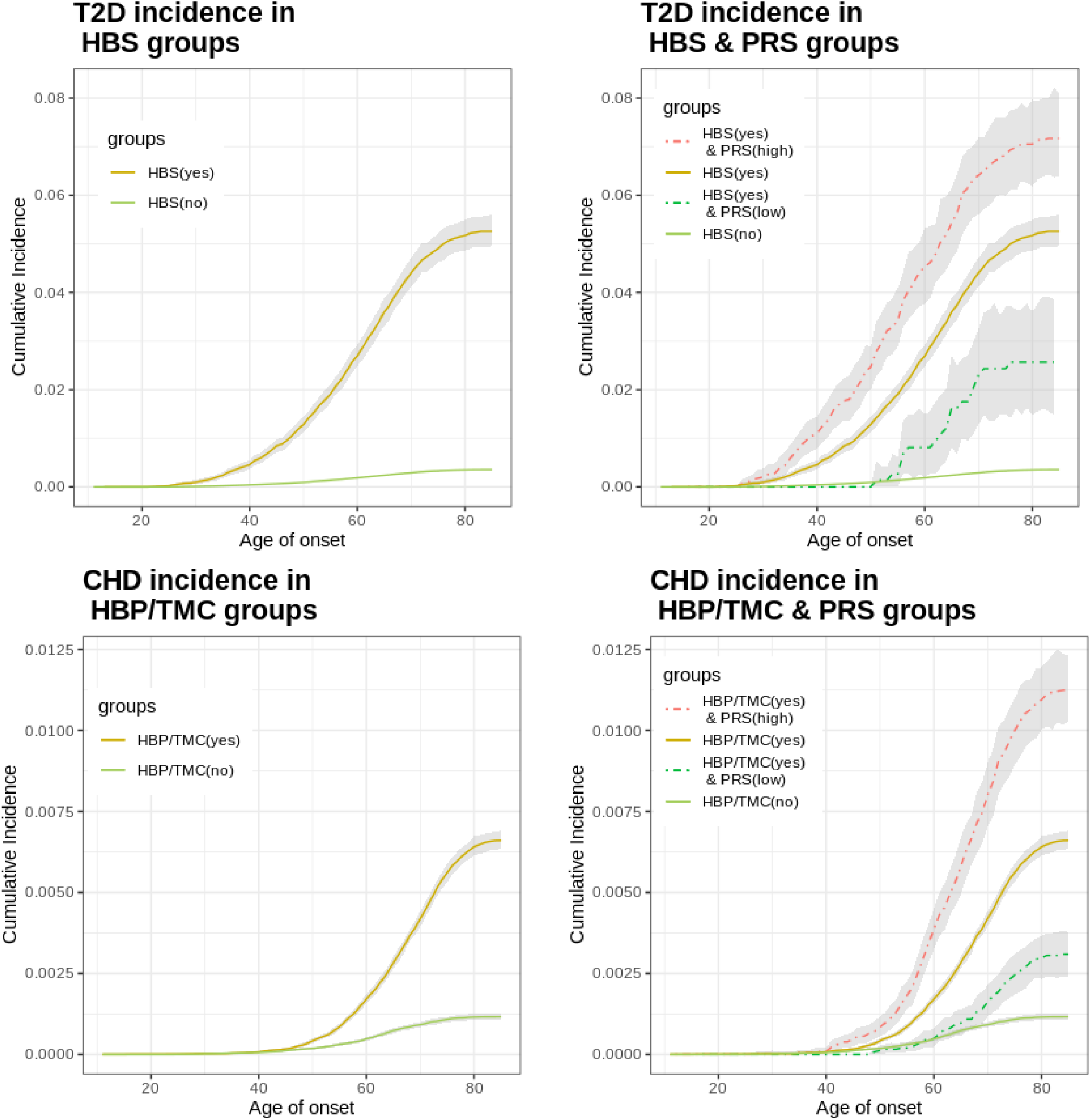
Risk stratification of incident T2D and CHD using PRS among individuals considered conventionally to be at high risk. The plots on the left hand side illustrate the higher cumulative incidence arising from conventional (phenotypically-defined) definitions of high risk. For T2D, this constitutes individuals who self-reported elevated blood sugar at baseline. For CHD, high risk comprises individuals who reported presence of hypertension and/or receiving treatment with lipid lowering therapy. The plots on the right hand side demonstrate the additive effect of PRS onto conventional risk factors. The red dotted line indicates those with the conventional risk factors who are also in the top 10% of the disease-relevant PRS whereas the green dotted line represents individuals with the conventional risk factor who are in the bottom 10% of the PRS. The grey shading around each line indicates the 95% CI from bootstrapping. HBP: high blood pressure; TMC: took medication for cholesterol

Individuals who reported presence of elevated blood glucose at baseline had a cumulative incidence of T2D that was 14.85-fold (95% CI: [13.84, 15.99]) greater than other individuals in the prospective cohort. Among these individuals with elevated blood glucose, the cumulative incidence of T2D was an additional 1.36-fold higher in those who were in the high T2D PRS. Among those with elevated blood glucose and high T2D PRS, the RR was 20.24-fold (95% CI: [17.71, 23.09]) higher than individuals who did not report elevated blood glucose (**Figure 3**).

### Effect of lifestyle on disease incidence among those at high polygenic risk

Among individuals in the top 10% of the CHD PRS, those who also had favorable lifestyle characteristics had a 73% relative risk reduction of disease, which yielded a cumulative incidence that was lower than the whole prospective cohort irrespective of genotype or lifestyle (**Figure 4**).

**Figure 4.**
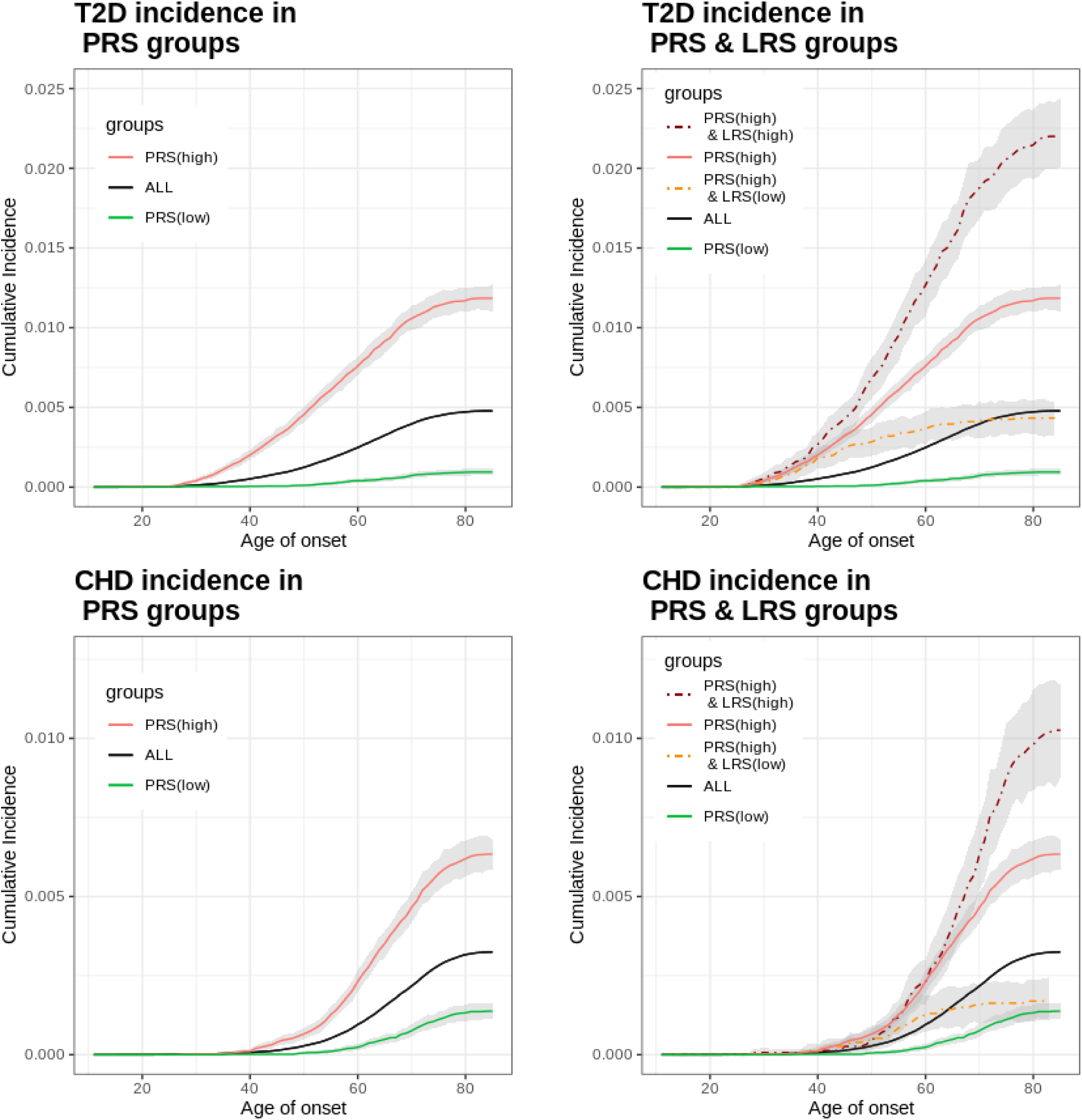
Impact of lifestyle factors on incident T2D and CHD among individuals at high polygenic risk. The plots on the left hand side illustrate the cumulative incidence of disease in everyone (black line), in those in the top 10% of the disease-relevant PRS (red line) and those in the bottom 10% (green line). The “all” group is represented rather than the middle 80% as in previous figures since we want to be able to illustrate the effect of lifestyle on changing risk as compared to the population average. The right hand plots demonstrate the impact of lifestyle factors among individuals at high polygenic risk of disease. The dark red dotted line represents those individuals in the top 10% of the disease-relevant PRS who also have lifestyle factors associated with higher risk of disease (in the top 20% of the disease-relevant LRS). The orange dotted line represents individuals in the top 10% of the disease-relevant PRS who have lifestyle factors associated with lower risk of disease (in the bottom 20% of the disease-relevant LRS). The grey shading around each line indicates the 95% CI from bootstrapping

For T2D, individuals in the top 10% of the T2D PRS who had a favorable lifestyle had a 64% relative risk reduction, yielding a cumulative incidence of T2D comparable to the entire prospective cohort irrespective of genotype or lifestyle (**Figure 4**).

The top four variables accounting for 47% of feature importance for the CHD lifestyle risk score were smoking, exercise, alcohol and salt. For T2D, the corresponding variables were exercise, alcohol, red meat and sugar, accounting for 53% of feature importance (**Figure S5**).

## 4 Discussion

In this study, we used a contemporaneous, diverse, US dataset to construct *de novo* PRS and evaluated predictive performance for incident disease. Across 12 clinical endpoints, individuals in the top decile of PRS had a higher incidence and, typically, an earlier age of onset. PRSs provided additional risk stratification beyond conventional risk factors, highlighting their incremental utility. Lifestyle factors diminished the excess risk arising from complex disease genetics for T2D and CHD, which represent the leading causes of morbidity and mortality worldwide.

To our knowledge, the sample size of our study for evaluating PRS and disease incidence is the largest, and most diverse, to date including a discovery dataset of over 7M (including 1.1M Latine and 0.4M African American) and a prospective cohort of over 1M (including 127K Latine and 33K African American). This allowed us to provide reliable measures of disease incidence stratified by PRS during one year of follow-up and longer (**Figure S4**).

A major addition of this study to the field is the derivation and evaluation of PRS for incident disease in non-European populations. While the C-statistic was consistent between Europeans and Latines, estimates for African Americans were typically lower. The RR estimates of top vs bottom 10% PRS were broadly similar for Europeans and Latines yet estimates for African Americans were more modest (**Figure S9**). In Thompson *et al.*^16^ the effect size of PRS for disease risk was consistently lower in non-Europeans, likely reflective of PRS being constructed from predominantly European ancestry individuals. Using an optimized PRS from a chronic kidney disease GWAS consortium, Khan *et al.*^17^ identified weaker disease associations of PRS in African Americans. Ge *et al*.^18^ used a Bayesian trans-ancestral approach to PRS derivation yielding comparable RRs of PRS across ancestrally-defined groups. Our findings provide empirical evidence in support of the critical value of ancestrally-diverse datasets, as evidenced by the weaker estimates of predictive utility seen in African American individuals.

Whether PRS are informative beyond conventionally-measured risk factors is a commonly-posed question^12^. Measures related to blood pressure and blood lipids feature in both QRISK3^19^ and Pooled Cohort Equations^20^ for estimating CVD risk. In our dataset, hypertensive individuals or those receiving lipid lowering therapy were at 5.69 fold higher risk of incident CHD than others. Those also in the top 10% CHD PRS had an even higher RR of 9.68. Individuals reporting high blood glucose were at 14.85-fold higher risk of T2D, and the RR was 20.24 among those also in the top 10% of the T2D PRS. These findings indicate the potential value for PRS in identifying high-risk groups, who may benefit from enhanced clinical care (e.g. closer monitoring, earlier intervention^21^).

We found strong effects of lifestyle on modifying the risk arising from polygenic susceptibility to disease. This is in keeping with prior studies examining CHD^22,23^, while adding important new insights from a contemporaneous US dataset and finding similar results for T2D. The main lifestyle constituents for CHD (smoking, exercise, alcohol and salt intake) and T2D (exercise, alcohol, red meat and sugar intake) include several that have established roles in disease etiology^24–26^. Our findings point to a potential strategy for PRS to advocate lifestyle changes and counter excess risk of disease.

Our study has several strengths. First, in evaluating performance in a prospective cohort, we emulate how PRS would be used clinically to predict future events. Second, the ancestral diversity builds on previously-conducted studies that are predominantly European. Third, exploring multiple incident diseases highlights the unique value proposition of PRS. Fourth, that lifestyle characteristics profoundly impact the risk of polygenic susceptibility to disease reinforces that genetics are not deterministic. Fifth, in addition to identifying individuals at elevated risk, PRS also impacted age of clinical onset: 16 years earlier for hypertension and 10 years for T2D. Predicting age of onset and detecting disease earlier in the natural course of clinical manifestation potentially avoids disease-related sequelae manifest in morbidity and premature death. Finally, our PRSs provide strong risk stratification for events occurring within only one year of follow-up, illustrating their immediate clinical relevance in a contemporaneous adult population.

Our study also has several limitations. First, all participants sought direct-to-consumer genotyping, which might impact external validity. Second, lifestyle traits were not reported in all individuals, potentially leading to bias, and being self-reported, may be prone to measurement error. Third, self-reported incident case counts may be influenced by some study participants not being blinded to selected PRS results^27^. Fourth, fewer individuals of non-European ancestry meant limited cases for some endpoints.

In summary, using the largest and most ancestrally diverse datasets of its kind, we used PRS to characterize the cumulative incidence of multiple diseases. PRS identified individuals at higher risk of incident diseases within one year across multiple ancestries, and had additional risk stratification utility among individuals conventionally thought to be at high risk of disease. Lifestyle factors ameliorated the excess polygenic risk of disease for T2D and CHD, two of the leading causes of morbidity and mortality in the US and worldwide. Our findings add important credibility to the utility of PRS among individuals of diverse ancestries in the short-term.

## Data Availability

To be updated at the time of publication.

## Acknowledgements

We would like to thank the research participants and employees of 23andMe for making this work possible.

Wei Wang, Nicholas Eriksson, Matthew McIntyre, Rafaela Bagur Quetglas, Bertram L. Koelsch, David A. Hinds, Stella Aslibekyan, 23andMe Research Team, Adam Auton, Michael V. Holmes, Suyash S. Shringarpure are employed by and hold stock or stock options in 23andMe, Inc.

## Notes

### Competing Interest Statement

W.W., N.E., M.M., R.B.Q., B.L.K, D.A.H, S.A., A.A., M.V.H. and S.S.S. are current or former employees of 23andMe and hold stock or stock options in 23andMe.

### Funding Statement

This study did not receive any funding.

### Author Declarations

Participants in this study were selected from the research participant base of 23andMe, a personal genetics company. Participants provided informed consent and participated in the research online, under a protocol approved by the external AAHRPP-accredited IRB, Ethical & Independent Review Services (E&I Review, now Salus IRB). Participants were included in the analysis on the basis of consent status as checked at the time data analyses were initiated.

## References

1. Ockene JK, Edgerton EA, Teutsch SM, et al. Integrating Evidence-Based Clinical and Community Strategies to Improve Health. Am J Prev Med 2007;32(3):244–52.

2. Khera R, Pandey A, Ayers CR, et al. Performance of the Pooled Cohort Equations to Estimate Atherosclerotic Cardiovascular Disease Risk by Body Mass Index. Jama Netw Open 2020;3(10):e2023242.

3. Lindström J, Tuomilehto J. The Diabetes Risk Score. Diabetes Care 2003;26(3):725–31.

4. Vilhjalmsson B, Yang J, Finucane HK, et al. Modeling Linkage Disequilibrium Increases Accuracy of Polygenic Risk Scores. Biorxiv 2015;015859.

5. Mak TSH, Porsch RM, Choi SW, Zhou X, Sham PC. Polygenic scores via penalized regression on summary statistics. Genet Epidemiol 2017;41(6):469–80.

6. Mars N, Koskela JT, Ripatti P, et al. Polygenic and clinical risk scores and their impact on age at onset and prediction of cardiometabolic diseases and common cancers. Nat Med 2020;26(4):549–57.

7. Khera AV, Chaffin M, Aragam KG, et al. Genome-wide polygenic scores for common diseases identify individuals with risk equivalent to monogenic mutations. Nat Genet 2018;50(9):1219–24.

8. Martin AR, Kanai M, Kamatani Y, Okada Y, Neale BM, Daly MJ. Clinical use of current polygenic risk scores may exacerbate health disparities. Nat Genet 2019;51(4):584–91.

9. Claussnitzer M, Cho JH, Collins R, et al. A brief history of human disease genetics. Nature 2020;577(7789):179–89.

10. Sirugo G, Williams SM, Tishkoff SA. The Missing Diversity in Human Genetic Studies. Cell 2019;177(1):26–31.

11. Whose genomics? Nat Hum Behav 2019;3(5):409–10.

12. O’Sullivan JW, Raghavan S, Marquez-Luna C, et al. Polygenic Risk Scores for Cardiovascular Disease: A Scientific Statement From the American Heart Association. Circulation 2022;146(8):e93–118.

13. Duncan L, Shen H, Gelaye B, et al. Analysis of polygenic risk score usage and performance in diverse human populations. Nat Commun 2019;10(1):3328.

14. Privé F, Vilhjálmsson BJ, Aschard H, Blum MGB. Making the Most of Clumping and Thresholding for Polygenic Scores. Am J Hum Genetics 2019;105(6):1213–21.

15. Krishnapuram B, Shah M, Smola A, et al. XGBoost: A Scalable Tree Boosting System. Arxiv 2016;785–94.

16. Thompson DJ, Wells D, Selzam S, et al. UK Biobank release and systematic evaluation of optimised polygenic risk scores for 53 diseases and quantitative traits. Medrxiv 2022;2022.06.16.22276246.

17. Khan A, Turchin MC, Patki A, et al. Genome-wide polygenic score to predict chronic kidney disease across ancestries. Nat Med 2022;28(7):1412–20.

18. Ge T, Irvin MR, Patki A, et al. Development and validation of a trans-ancestry polygenic risk score for type 2 diabetes in diverse populations. Genome Med 2022;14(1):70.

19. Hippisley-Cox J, Coupland C, Brindle P. Development and validation of QRISK3 risk prediction algorithms to estimate future risk of cardiovascular disease: prospective cohort study. Bmj 2017;357:j2099.

20. Stone NJ, Robinson JG, Lichtenstein AH, et al. 2013 ACC/AHA Guideline on the Treatment of Blood Cholesterol to Reduce Atherosclerotic Cardiovascular Risk in Adults A Report of the American College of Cardiology/American Heart Association Task Force on Practice Guidelines. J Am Coll Cardiol 2014;63(25):2889–934.

21. Taylor R, Ramachandran A, Yancy WS, Forouhi NG. Nutritional basis of type 2 diabetes remission. Bmj 2021;374:n1449.

22. Khera AV, Emdin CA, Drake I, et al. Genetic Risk, Adherence to a Healthy Lifestyle, and Coronary Disease. New Engl J Medicine 2016;375(24):2349–58.

23. Hasbani NR, Ligthart S, Brown MR, et al. American Heart Association’s Life’s Simple 7: Lifestyle Recommendations, Polygenic Risk, and Lifetime Risk of Coronary Heart Disease. Circulation 2021;145(11):808–18.

24. Cifu AS, Davis AM. Prevention, Detection, Evaluation, and Management of High Blood Pressure in Adults. Jama 2017;318(21):2132.

25. Ockene IS, Miller NH. Cigarette Smoking, Cardiovascular Disease, and Stroke: A Statement for Healthcare Professionals From the American Heart Association. Circulation 1997;96(9):3243–7.

26. Marshall JA, Bessesen DH. Dietary Fat and the Development of Type 2 Diabetes. Diabetes Care 2002;25(3):620–2.

27. Ashenhurst JR*, Zhan J*, Multhaup ML, et al. White Paper 23-21 A Generalized Method for the Creation and Evaluation of Polygenic Scores. Available from: https://permalinks.23andme.com/pdf/23_21-PRSMethodology_May2020.pdf

