## Supplementary materials for "Prospective analysis of incident disease among individuals of diverse ancestries using genetic and conventional risk factors"

* joint senior authors

### Table of Contents

**23andMe Research Team** 4
**Supplementary Methods** 5
- **S.1 Discovery cohort and prospective cohort** 5
- **S.2 GWAS information** 5
- **S.3 PRS model** 8
- **S.4 Lifestyle Risk Score model** 9
- **S.5 Incidence cases/controls** 9

**Supplementary Results** 10
- **S.1 Disease incidence in Europeans** 10

**Supplementary Figures** 12
- **Fig S1. Schematic of the dataset and analysis.** 12
- **Fig S2.A. Risk stratification of prevalent clinical endpoints through PRS.** 13
- **Fig S2.B Risk stratification of prevalent clinical endpoints through PRS.** 14
- **Fig S2.C Risk stratification of prevalent clinical endpoints through PRS.** 15
- **Fig S3 Risk stratification of prevalent and incident clinical endpoints through PRS in**

African American population. 16
Fig S4.A Incidence of endpoints by strata defined by PRS during 3 years follow-up 17
Fig S4.B Incidence of endpoints by strata defined by PRS during 3 years follow-up 18
Fig S4.C Incidence of endpoints by strata defined by PRS during 3 years follow-up 19
Fig S4.D Incidence of endpoints by strata defined by PRS during 3 years follow-up 20
Fig S5.A Feature importance plot from CHD LRS model. 21
Fig S5.B Feature importance plot from T2D LRS model. 22
Fig S6.A PRS performance for anemia in testing data from discovery dataset (European population) 23
Fig S6.B PRS performance for blood clots in testing data from discovery dataset (European population) 24
Fig S6.C PRS performance for coronary heart disease in testing data from discovery dataset (European population) 25
Fig S6.D PRS performance for colon polyps in testing data from discovery dataset (European population) 26
Fig S6.E PRS performance for Crohn's in testing data from discovery dataset (European population) 27
Fig S6.F PRS performance for eczema in testing data from discovery dataset (European population) 28
Fig S6.G PRS performance for glaucoma in testing data from discovery dataset (European population) 29
Fig S6.H PRS performance for high blood pressure in testing data from discovery dataset (European population) 30
Fig S6.I PRS performance for hypothyroidism in testing data from discovery dataset (European population) 31
Fig S6.J PRS performance for kidney stones in testing data from discovery dataset (European population) 32
Fig S6.K PRS performance for psoriasis in testing data from discovery dataset (European population) 33
Fig S6.L PRS performance for type 2 diabetes in testing data from discovery dataset (European population) 34
Fig S7.A PRS performance for anemia in testing data from discovery dataset (Latine population) 35
Fig S7.B PRS performance for blood clots in testing data from discovery dataset (Latine population) 36
Fig S7.C PRS performance for coronary heart disease in testing data from discovery dataset (Latine population) 37
Fig S7.D PRS performance for colon polyps in testing data from discovery dataset (Latine population) 38
Fig S7.E PRS performance for Crohn's in testing data from discovery dataset (Latine population) 39
Fig S7.F PRS performance for eczema in testing data from discovery dataset (Latine population) 40

**Fig S7.G PRS performance for glaucoma in testing data from discovery dataset (Latine population)** **41**
**Fig S7.H PRS performance for high blood pressure in testing data from discovery dataset (Latine population)** **42**
**Fig S7.I PRS performance for hypothyroidism in testing data from discovery dataset (Latine population)** **43**
**Fig S7.J PRS performance for kidney stones in testing data from discovery dataset (Latine population)** **44**
**Fig S7.K PRS performance for psoriasis in testing data from discovery dataset (Latine population)** **45**
**Fig S7.L PRS performance for type 2 diabetes in testing data from discovery dataset (Latine population)** **46**
**Fig S8.A PRS performance for anemia in testing data from discovery dataset (African American population)** **47**
**Fig S8.B PRS performance for blood clots in testing data from Discovery dataset (African American population)** **48**
**Fig S8.C PRS performance for coronary heart disease in testing data from discovery dataset (African American population)** **49**
**Fig S8.D PRS performance for colon polyps in testing data from discovery dataset (African American population)** **50**
**Fig S8.E PRS performance for eczema in testing data from discovery dataset (African American population)** **51**
**Fig S8.F PRS performance for glaucoma in testing data from discovery dataset (African American population)** **52**
**Fig S8.G PRS performance for high blood pressure in testing data from Discovery dataset (African American population)** **53**
**Fig S8.H PRS performance for hypothyroidism in testing data from Discovery dataset (African American population)** **54**
**Fig S8.I PRS performance for kidney stones in testing data from discovery dataset (African American population)** **55**
**Fig S8.J PRS performance for psoriasis in testing data from discovery dataset (African American population)** **56**
**Fig S8.K PRS performance for type 2 diabetes in testing data from discovery dataset (African American population)** **57**
**Fig S9. PRS performance for diseases across multiple ancestries.** **58**
**Supplementary Tables** **59**
**Table S1. Clinical endpoint definitions in the discovery dataset** **59**
**Table S2. Clinical endpoint definition from baseline and during follow-up** **65**
**Table S3. Performance of PRS for incident endpoints across different ancestries** **68**
**Table S4. Lifestyle traits constituting the lifestyle risk score (LRS)** **70**
**References** **76**

### 23andMe Research Team

The following members of the 23andMe Research Team contributed to this study:

Stella Aslibekyan, Adam Auton, Elizabeth Babalola, Robert K. Bell, Jessica Bielenberg, Katarzyna Bryc, Emily Bullis, Daniella Coker, Devika Dhamija, Sayantan Das, Sarah L. Elson, Nicholas Eriksson, Teresa Filshtein, Alison Fitch, Pierre Fontanillas, Will Freyman, Julie M. Granka, Karl Heilbron, Alejandro Hernandez, Barry Hicks, David A. Hinds, Michael V. Holmes, Ethan M. Jewett, Yunxuan Jiang, Katelyn Kukar, Alan Kwong, Keng-Han Lin, Bianca A. Llamas, Maya Lowe, Matthew H. McIntyre, Steven J. Micheletti, Meghan E. Moreno, Priyanka Nandakumar, Dominique T. Nguyen, Jared O'Connell, Aaron A. Petrakovitz, G. David Poznik, Alexandra Reynoso, Morgan Schumacher, Anjali J. Shastri, Janie F. Shelton, Jingchunzi Shi, Suyash Shringarpure, Qiaojuan Jane Su, Susana A. Tat, Vinh Tran, Joyce Y. Tung, Xin Wang, Wei Wang, Catherine H. Weldon, Peter Wilton, Corinna D. Wong.

### Supplementary Methods

#### S.1 Discovery cohort and prospective cohort

Individuals in both the discovery dataset and prospective cohort provided details on prevalent disease status in an initial “Health Profile” survey: questions were framed around current or past disease status, e.g. “Have you ever been diagnosed with or treated for any of the following conditions?”. Following completion of baseline information, participants were contacted on an annual basis to answer a “Health Update” survey, with questions framed around changes to disease status in the past year, e.g. “In the past 12 months, have you been newly diagnosed with any of the following conditions by a medical professional?”. Individuals who answered the Health Update survey in the first three instances were placed in the prospective cohort, and the remainder became the discovery cohort. From these surveys, we obtained information on prevalent and incident disease. Furthermore, we identified additional prevalent cases using information from other surveys (see **Table S1, S2** for details). In the discovery cohort, we randomly divided the discovery cohort into three non-overlapping data sets, GWAS-training data, PRS-training data and testing data with sample size proportions as 70%, 20% and 10%. We conducted GWAS using the GWAS-training data, fitted PRS models using the PRS-training data, and used the testing data to evaluate the out-of-sample prediction performance in the discovery cohort (**Figure S1**).

#### S.2 GWAS information

DNA extraction and genotyping were performed on saliva samples by Clinical Laboratory Improvement Amendments-certified and College of American Pathologists-accredited clinical laboratories of Laboratory Corporation of America. Participants were genotyped on one of five genotyping platforms. The v1 and v2 platforms were based on the Illumina HumanHap550+ BeadChip, including about 25,000 custom variants selected by 23andMe, with a total of about 560,000 SNPs. The v3 platform was based on the

Illumina OmniExpress+ BeadChip, with custom content to improve the overlap with our v2 array, with a total of about 950,000 variants. The v4 platform was a fully customized array, including a lower redundancy subset of v2 and v3 variants with additional coverage of lower-frequency coding variation, and about 570,000 variants. The v5 platform, in current use, is an Illumina Infinium Global Screening Array (~640,000 variants) supplemented with ~50,000 variants of custom content. This array was specifically designed to better capture global worldwide genetic diversity and to help standardize the platform for genetic research. Samples that failed to reach 98.5% call rate were re-genotyped. Participants whose analyses failed repeatedly were re-contacted by 23andMe customer service to provide additional samples. A total of number of 1,522,458 variants have been genotyped across the five genotyping platforms.

Participant genotype data were imputed using an imputation panel combining two independent reference panels: the publicly available Human Reference Consortium[1] (HRC) panel and the 23andMe reference panel, which was built by 23andMe using internal and external cohorts. We phased and imputed data for each genotyping platform separately.

23andMe ancestry classifier algorithm determines participant ancestries through an analysis of local ancestry[2]. It first partitions phased genotyped data into short windows of about 300 SNPs. Within each window, we use a support vector machine (SVM) to classify individual haplotypes into one of 45 worldwide reference populations. The SVM classifications are then fed into a hidden Markov model (HMM) that accounts for switch errors and incorrect assignments, and gives probabilities for each reference population in each window. Finally, we used simulated admixed individuals to recalibrate the HMM probabilities so that the reported assignments are consistent with the simulated admixture proportions. We are aggregating the probabilities of the 45 reference populations into ancestry groups (for example, European genetic ancestry: European + Middle Eastern > 0.97, European > 0.90; African American + Latine genetic ancestries:

European + African + East Asian + Native American + Middle Eastern > 0.90, African + Native American > 0.01). African Americans and Latine are admixed with broadly varying contributions from Europe, Africa and the Americas. Therefore, no single threshold of genome-wide ancestry will be able to effectively classify African Americans and Latine. However, the distributions of the length of segments of European, African and American ancestry are very different between African Americans and Latine, because of distinct admixture timing between the three ancestral populations in the two ethnic groups. Therefore, we trained a logistic classifier that takes one customer's length histogram of segments of African, European and American ancestry, and predict whether the customer is likely African American or Latine.

A maximal set of unrelated individuals is chosen for each GWAS analysis using a segmental identity-by-descent (IBD) estimation algorithm[3]. Participants are defined as related if they shared more than 700 cM IBD, including regions where the two individuals share either one or both genomic segments IBD. This level of relatedness (roughly 20% of the genome) corresponds approximately to the minimal expected sharing between first cousins in an outbred population. When selecting individuals for case/control phenotype analyses, the selection process is designed to maximize case sample size by preferentially retaining cases over controls. Specifically, if both an individual case and an individual control are found to be related, then the case is retained in the analysis.

Variants that were only genotyped on the “V1” platform were flagged due to small sample size, and variants on chrM or chrY, because many of these are not currently called reliably. Using trio data, variants that failed a test for parent–offspring transmission were also flagged; specifically, the child’s allele count was regressed against the mean parental allele count and variants with fitted $\beta < 0.6$ and $P < 10^{-20}$ for a test of $\beta < 1$ were flagged. Variants with a Hardy–Weinberg $P < 10^{-20}$ in Europeans, or a call rate of <90%, were also flagged. Genotyped variants were also tested for batch effects and variants with $P < 10^{-50}$ by analysis of variance of genotypes against a factor dividing genotyping date into 20 roughly equal-sized buckets were flagged. For imputed GWAS results, we flagged SNPs with an imputation $r^2$<0.5, as well as SNPs that had strong evidence of a platform batch effect. The batch effect test is an F test from an ANOVA of the SNP dosages against a factor splitting participants into 20 bins by imputation date; we flagged results with $P<10^{-50}$. Each variant flagged by QC on genotyped or imputation data were excluded from the GWAS analysis.

#### S.3 PRS model

We built PRS based on GWAS summary statistics and individual level data from the PRS-training data portion of the discovery cohort (**Figure S1**). Using the GWAS summary statistics from each population, we calculated pruning and thresholding (P+T) scores using PLINK[4] for samples in the PRS-training data. The P+T scores were calculated using different P-value thresholds and LD pruning parameters. We filtered the GWAS summary statistics based on minor allele frequency (MAF), excluding variants with MAF less than 0.1%. We added additional variant screening based on imputation $r^2$, and set different thresholds for imputation $r^2$ as 0.5 and 0.95. The different P-values thresholds are set as 1e-1, 1e-2, 1e-3, 1e-4, 1e-5, 1e-6, 1e-7, 1e-8, 1e-9 and 1e-10, and the LD $r^2$ values in the LD pruning were set as 0.01, 0.05, 0.1, 0.2, 0.5 and 0.8. In the LD pruning step, the pruning distance was set as 500Kb, which means that the variants within 500Kb of the index variant would be considered for pruning. As such, we had a total of 120 (the total combinations of 2 imputation $r^2$ parameters, 10 P-value thresholding parameters and 6 LD pruning parameters) different parameter settings for the P+T PRS model in each population. We applied the stacked P+T approach using the Elastic-Net model with 10-fold cross validation to build the PRS model based on 360 P+T PRS scores across three populations. The stacked P+T approach on single population was proposed by Prive et al 2019[5]. In the testing data, we evaluated the PRS model performance based on area under the receiver operating characteristics curve (AUC) and the disease prevalence in groups binned according to PRS percentile in the testing dataset. These metrics were tested in a hold-out testing data from the discovery cohort on both PRS and PRS adjusted by age, sex, and the first 5 PCs (**Figure S6-S8, Table S3**).

#### S.4 Lifestyle Risk Score model

To construct the Lifestyle Risk Scores (LRS), we applied XGboost[6], a tree-based predictive model, on 40 lifestyle factors, for each disease outcome. The lifestyle factors are based on the first answers to the survey questions on diet, exercise, sleeping, smoking and alcohol consumption (Table S4). We built the lifestyle risk prediction model in the discovery dataset and applied the model to samples in the prospective cohort. In the discovery dataset, we trained the LRS model with 24 parameters settings (step size: 0.05, 0.1, 0.25, 0.5; maximum depth of a tree: 2, 3, 5; minimum loss reduction to split a leaf node: 0.1, 0.5; subsample ratio of the training instances: 0.75; subsample ratio of columns in constructing each tree: 0.6), we choose the optimal parameter setting based on AUC performance in testing data from the discovery dataset.

#### S.5 Incidence cases/controls

We defined incident cases in each follow-up year as individuals with “No” answers from the survey questions in all the previous instances and reported “Yes” to the follow-up questions from the health update surveys in the current instance (individuals who skipped a survey instance were excluded). We defined the controls of the incidence data as individuals with “No” answers in all instances from health history surveys. For example, 1st-year T2D incident cases were defined as individuals who reported “No” to the survey question, “Have you ever been diagnosed with or treated for any of the following conditions; type 2 diabetes”, during the enrollment, and reported “Yes” in the follow up questions from health update survey to update the disease status of T2D in the first following year. Controls were defined as those who reported “No” to both questions (Detailed survey questions are in **Table S2**).

We also conducted analyses among individuals at higher risk of incident disease denoted by conventional risk factors. For example, we calculated the incidence of T2D among individuals with self-reported high blood sugar at baseline. The cases at high risk of conventional (non-genetically measured) risk factors are defined as individuals who reported ‘No’ answers for T2D questions but with “Yes” answers for the high blood sugar questions in the health history surveys during enrollment (baseline), and reported “Yes” answers for the T2D follow-up questions in the health update survey, which means the samples had been diagnosed with high blood sugar but not with T2D during enrollment and had been diagnosed with T2D in the following years. The controls at high risk of conventional (non-genetically measured) risk factors were defined as samples with ‘No’ answers for T2D questions but with ‘Yes’ answers for the high blood sugar questions in the health history surveys, and still with ‘No’ answers for the T2D follow-up questions in the health update survey, which means the samples maintained the prediabetic status without any progression in the following years.

Analyses were conducted using R 3.6.3 and Python 2.7.9.

### Supplementary Results

#### S.1 Disease incidence in Europeans

*Incident CHD in Europeans*

Among Europeans in the prospective cohort, the cumulative incidence of CHD was 0.32% (95% CI: [0.31%, 0.34%]). For those in the top 10% of the CHD PRS, the incidence was 0.63% (95% CI: [0.57%, 0.70%]), and among those in the bottom 10% of the PRS it was 0.14% (95% CI: [0.12%, 0.17%]) (**Figure 1**). Comparing top to bottom deciles of the PRS this corresponds to an absolute difference of 0.50% (95% CI: [0.43%, 0.56%]) and a risk ratio of 4.61 (95% CI: [3.79, 5.81]). Those in the high PRS group on average developed CHD at 64 (14) yrs and those in the bottom PRS group at 70 (12) yrs, corresponding to an earlier  age of onset of 6 yrs (**Table S3**).

The relationship between age of onset and whether individuals were in the low, middle and high groups of the PRS is presented in the bar charts adjacent to the cumulative incidence plots in **Figure 1** in the main manuscript. For CHD, this demonstrates a step-wise shift in age across PRS groups, with a greater representation of cases among those younger than 60 yrs in the high PRS group.

*Incident T2D in individuals of European ancestry*

The cumulative incidence of T2D within the prospective cohort was 0.48% (95% CI: [0.46%, 0.49%]). Among those in the top 10% of the T2D PRS, it was 1.19% (95% CI: [1.11%, 1.27%]) and in those in the bottom 10% PRS it was 0.095% (95% CI: [0.07%, 0.12%]) (**Figure 1**). This corresponded to an absolute difference of 1.09% (95% CI: [1.01%, 1.17%]) and a relative risk of 12.53 (95% CI: [9.99, 16.43]) comparing high to low PRS groups. The average age of diagnosis was 60 (median; IQR 18) yrs with those in the high PRS group on average developing T2D at 55 (21) yrs and those in the bottom PRS group at 64.5 (14) yrs, corresponding to an average earlier age of onset of 9.5 yrs (**Table S3**). As for CHD, there was a stepwise change to an earlier average age of onset from low to middle to high groups of PRS.

*Age of diagnosis*

The relationship of PRS with age of diagnosis was less clear for eczema and Crohn's disease: it is possible that there were insufficient incident cases of disease and/or the underlying epidemiology of age of onset (e.g. a bimodal distribution) obscured our ability to identify an age effect of PRS for these health conditions.

### Supplementary Figures

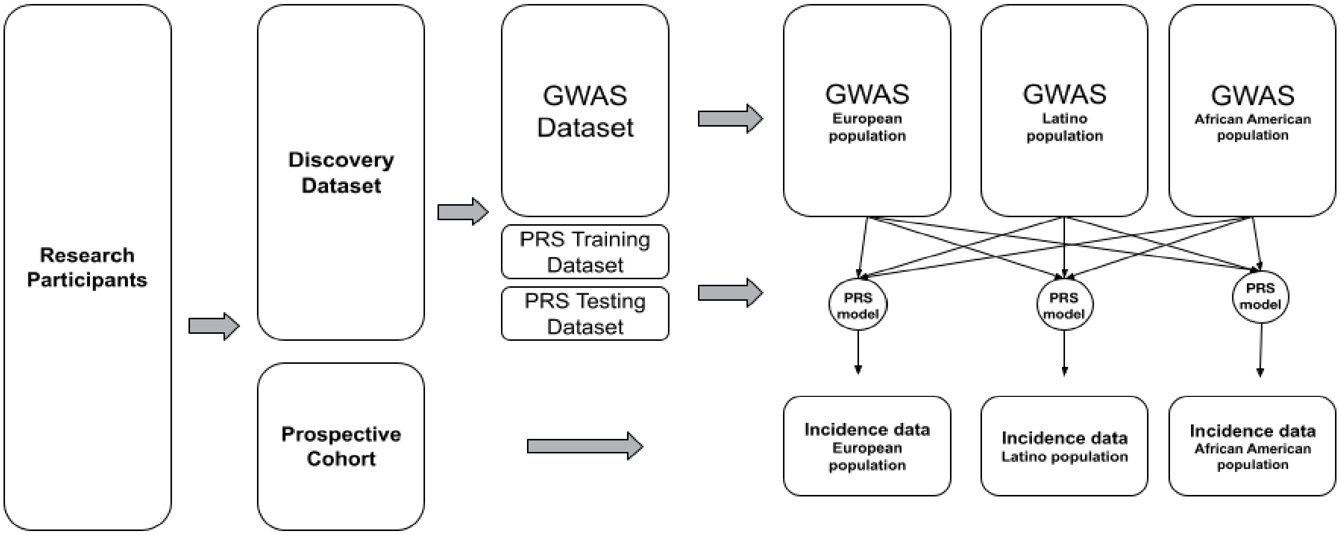


**Fig S1. Schematic of the dataset and analysis.**
Research participants were divided into discovery dataset and prospective cohort. GWAS and PRS were conducted in subsets of the discovery dataset, from which PRS were constructed based on summary statistics across three populations and applied to the prospective cohort, in order to evaluate the performance of PRS for incident disease.

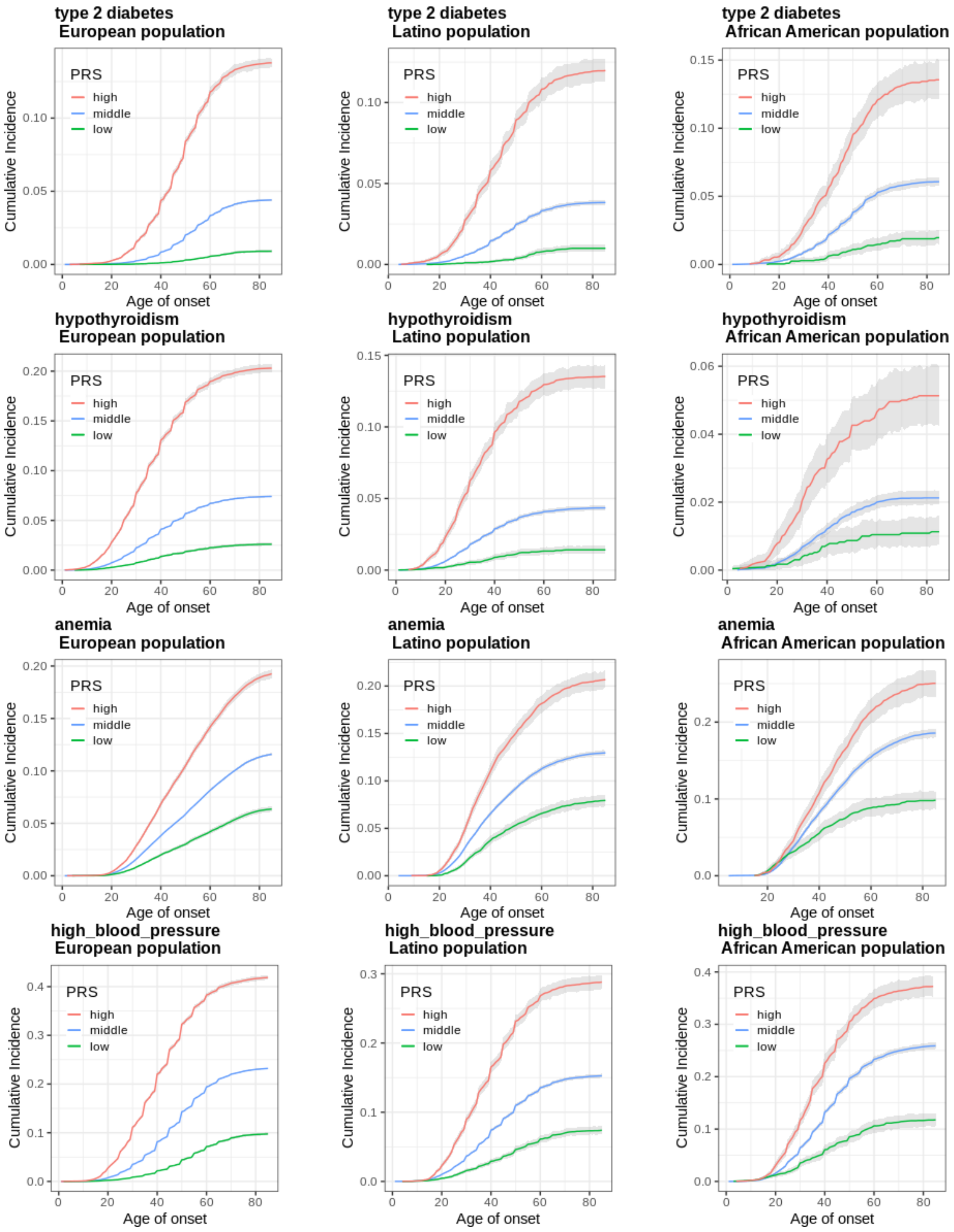


**Fig S2.A. Risk stratification of prevalent clinical endpoints through PRS.**

Each plot illustrates the (cumulative) prevalence among individuals from testing data of Discovery dataset and grouped by each disease-specific PRS. Individuals in the top 10% of each PRS are represented by a red line, those in the middle 80% by a blue line and those in the bottom 10% by a green line. The gray shading around each line indicates the 95% CI from bootstrapping.

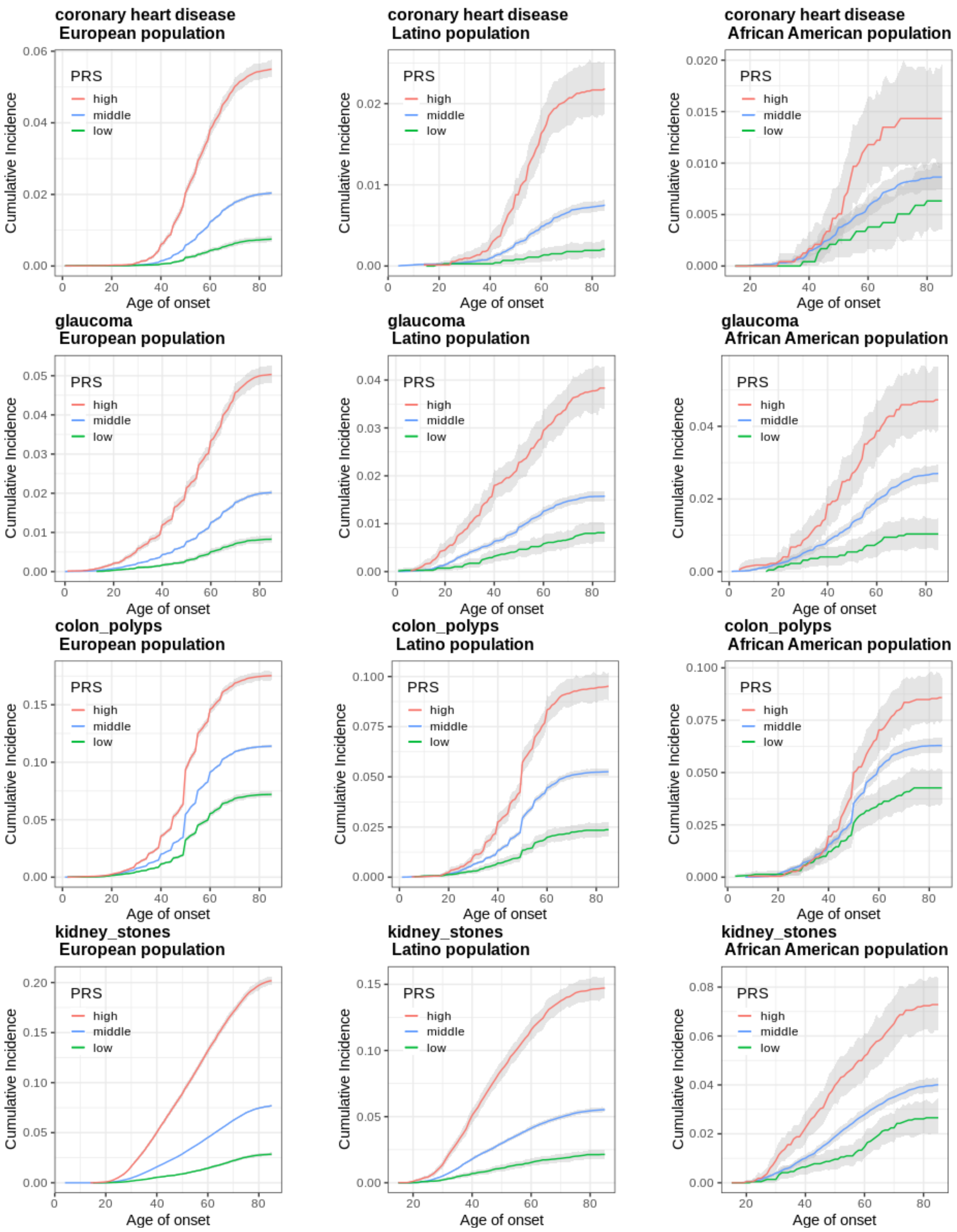


**Fig S2.B Risk stratification of prevalent clinical endpoints through PRS.**

Each plot illustrates the (cumulative) prevalence among individuals from testing data of Discovery dataset and grouped by each disease-specific PRS. Individuals in the top 10% of each PRS are represented by a red line, those in the middle 80% by a blue line and those in the bottom 10% by a green line. The gray shading around each line indicates the 95% CI from bootstrapping.

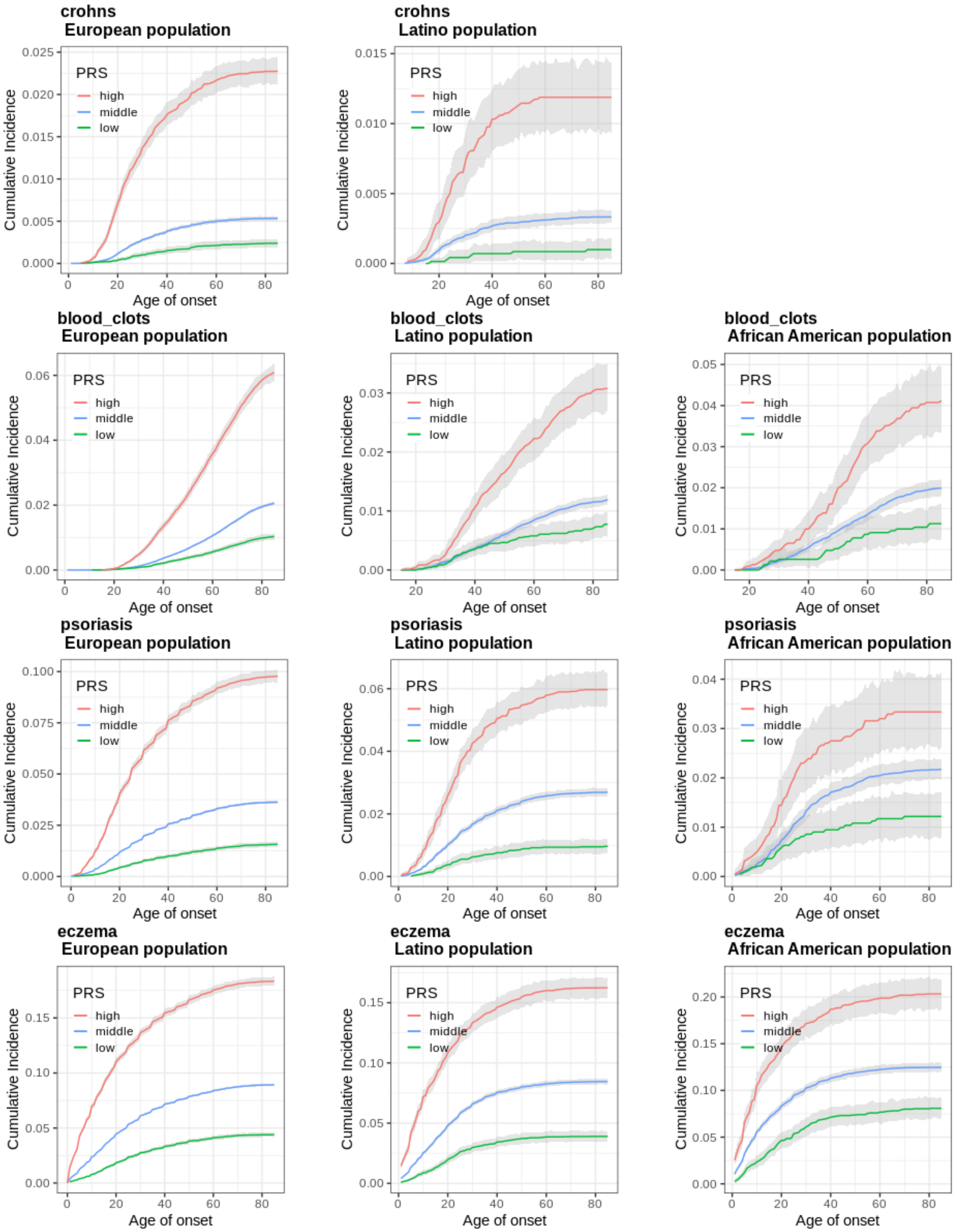


**Fig S2.C Risk stratification of prevalent clinical endpoints through PRS.**
Each plot illustrates the (cumulative) prevalence among individuals from testing data of Discovery dataset and grouped by each disease-specific PRS. Individuals in the top 10% of each PRS are represented by a red line, those in the middle 80% by a blue line and those in the bottom 10% by a green line. The gray shading around each line indicates the 95% CI from bootstrapping.

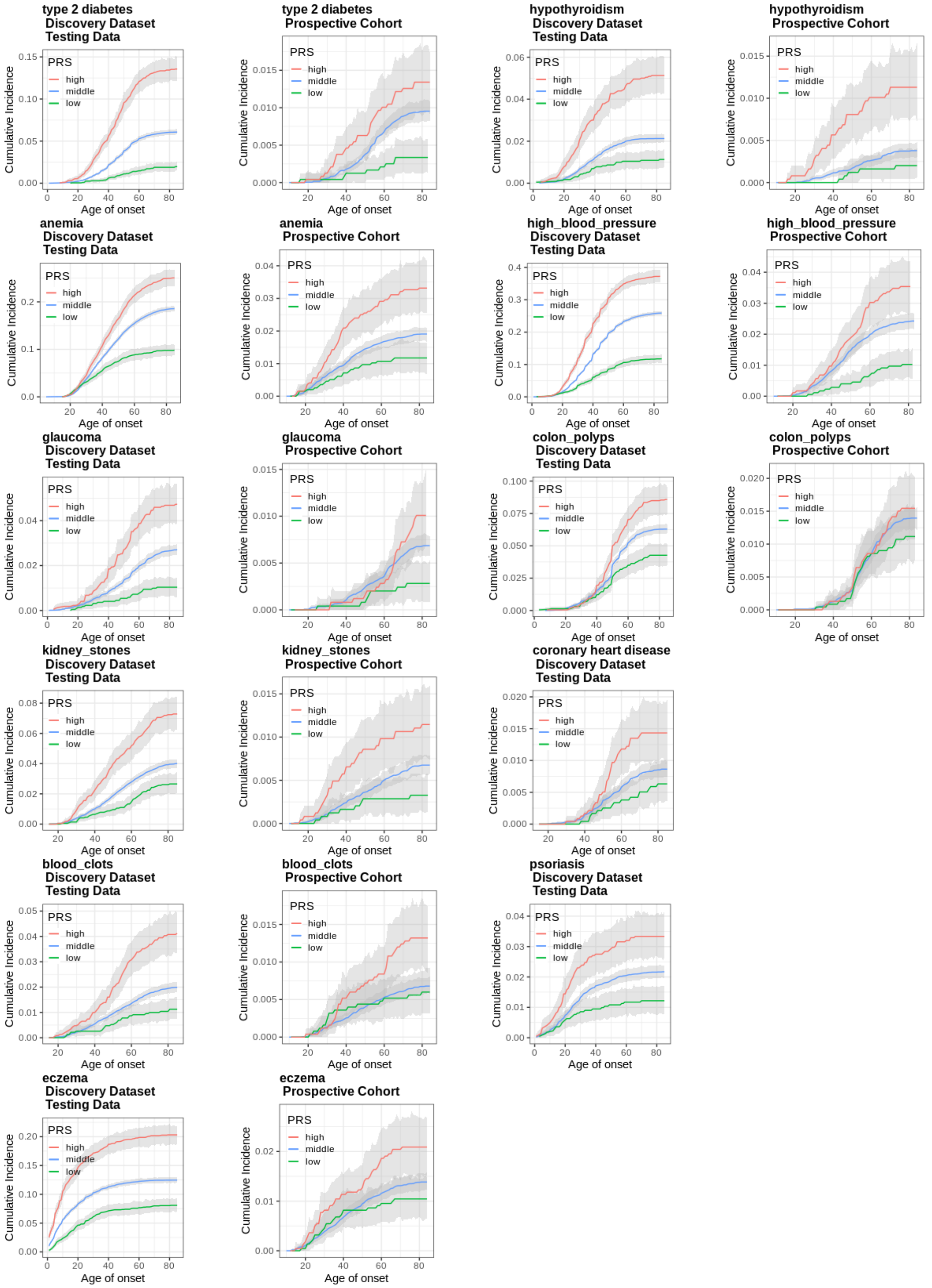


**Fig S3 Risk stratification of prevalent and incident clinical endpoints through PRS in African American population.**

Individuals in the top 10% of each PRS are represented by a red line, those in the middle 80% by a blue line and those in the bottom 10% by a green line. The gray shading around each line indicates the 95% CI from bootstrapping.

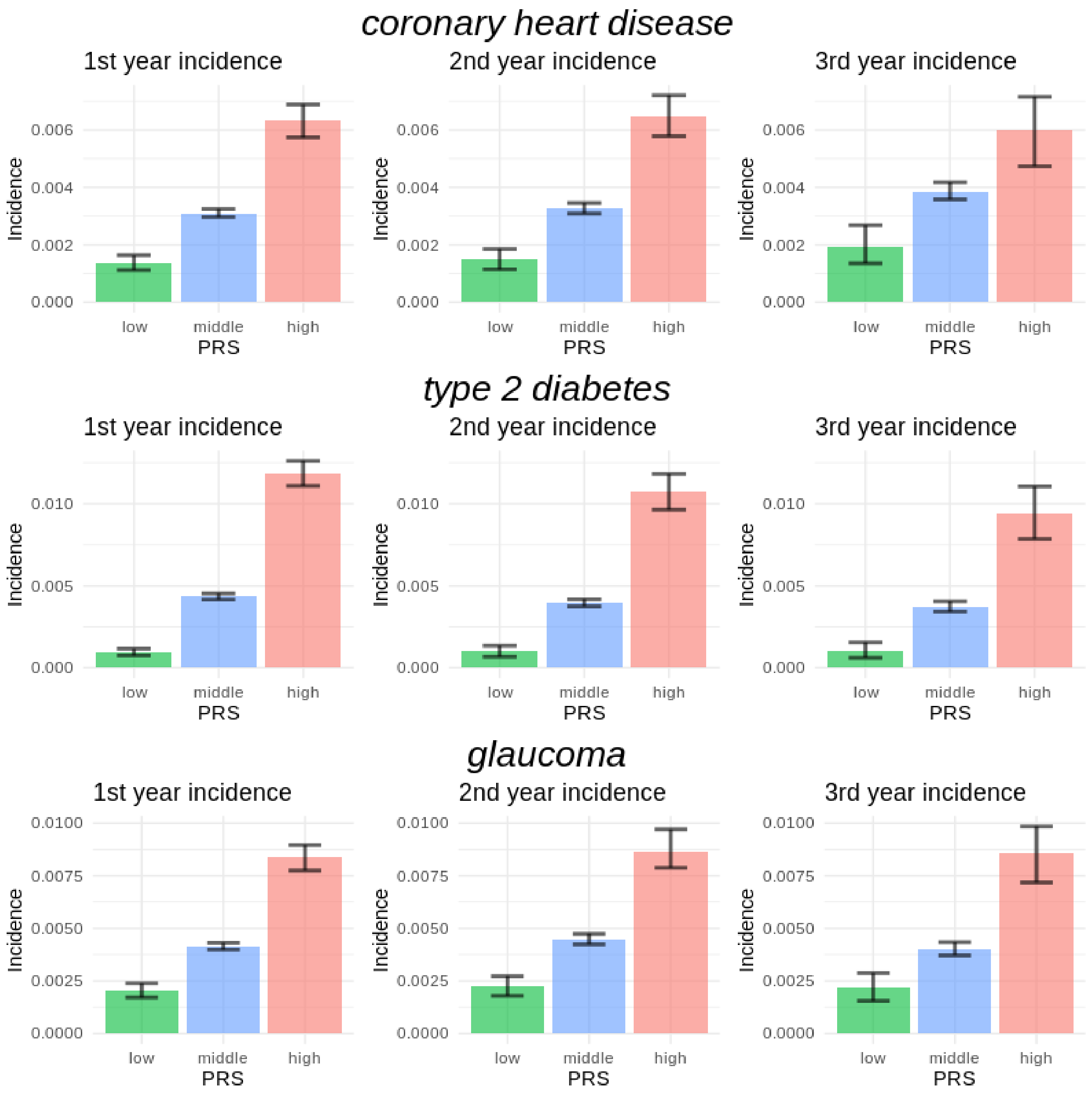


**Fig S4.A Incidence of endpoints by strata defined by PRS during 3 years follow-up**
Each plot illustrates the incidence over the 1st, 2nd, and 3rd year of follow-up among individuals grouped by each disease-specific PRS. Individuals in the top 10% of each PRS are represented by a red bar, those in the middle 80% by a blue bar and those in the bottom 10% by a green bar. The error bars indicate the 95% CI from bootstrapping.

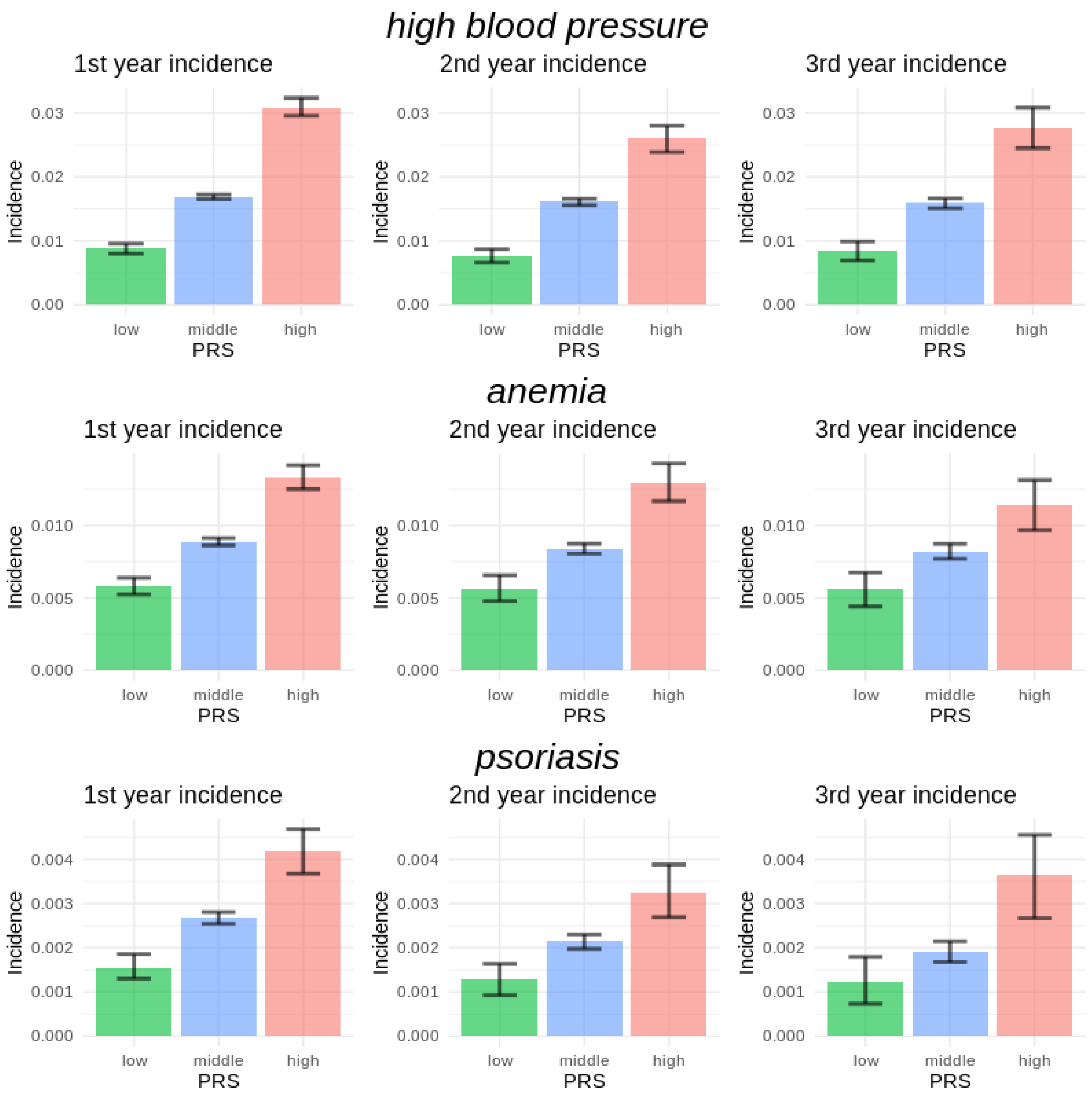


**Fig S4.B Incidence of endpoints by strata defined by PRS during 3 years follow-up**

Each plot illustrates the incidence over the 1st, 2nd, and 3rd year of follow-up among individuals grouped by each disease-specific PRS. Individuals in the top 10% of each PRS are represented by a red bar, those in the middle 80% by a blue bar and those in the bottom 10% by a green bar. The error bars indicate the 95% CI from bootstrapping.

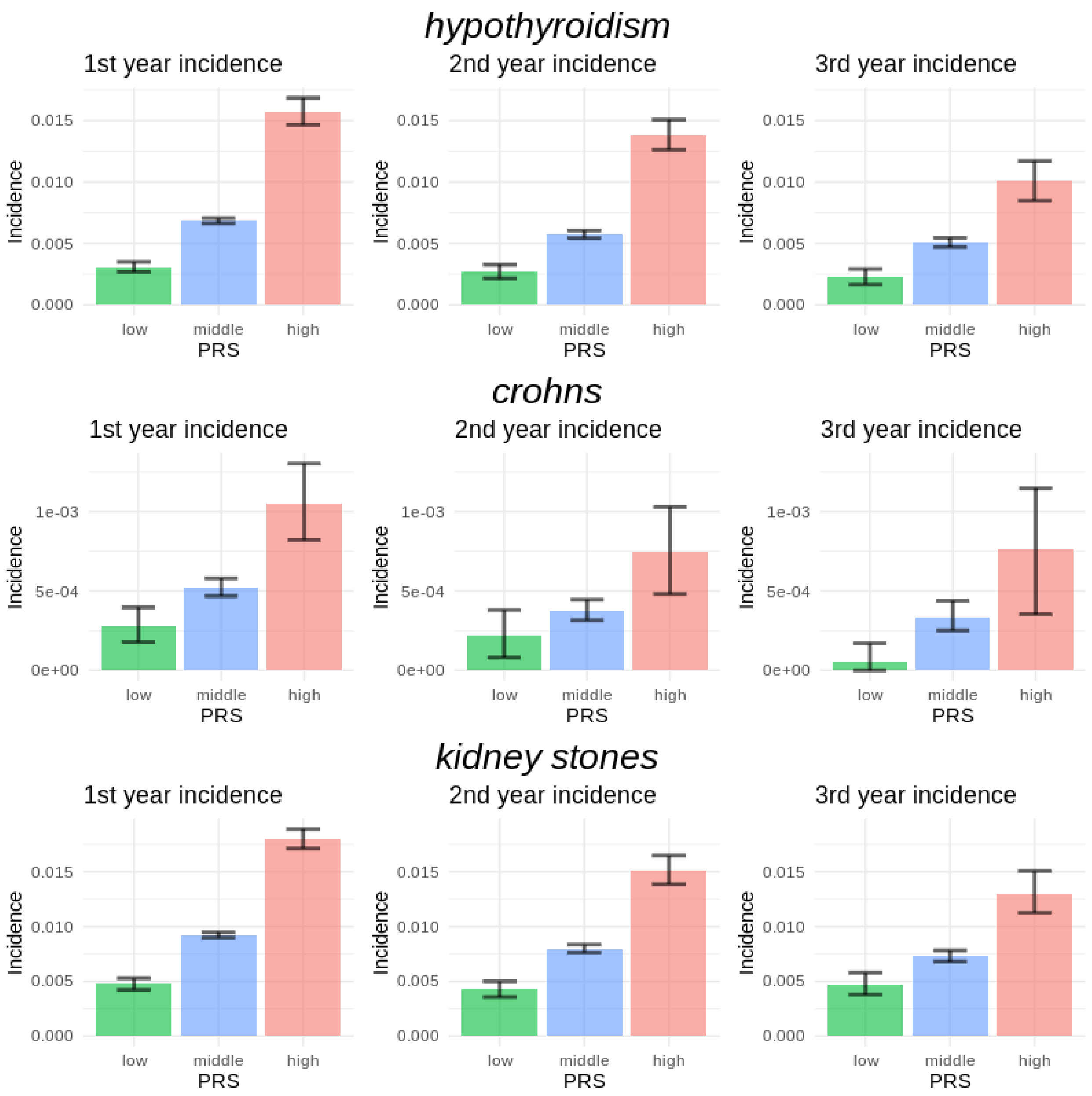


**Fig S4.C Incidence of endpoints by strata defined by PRS during 3 years follow-up**
Each plot illustrates the incidence over the 1st, 2nd, and 3rd year of follow-up among individuals grouped by each disease-specific PRS. Individuals in the top 10% of each PRS are represented by a red bar, those in the middle 80% by a blue bar and those in the bottom 10% by a green bar. The error bars indicate the 95% CI from bootstrapping.

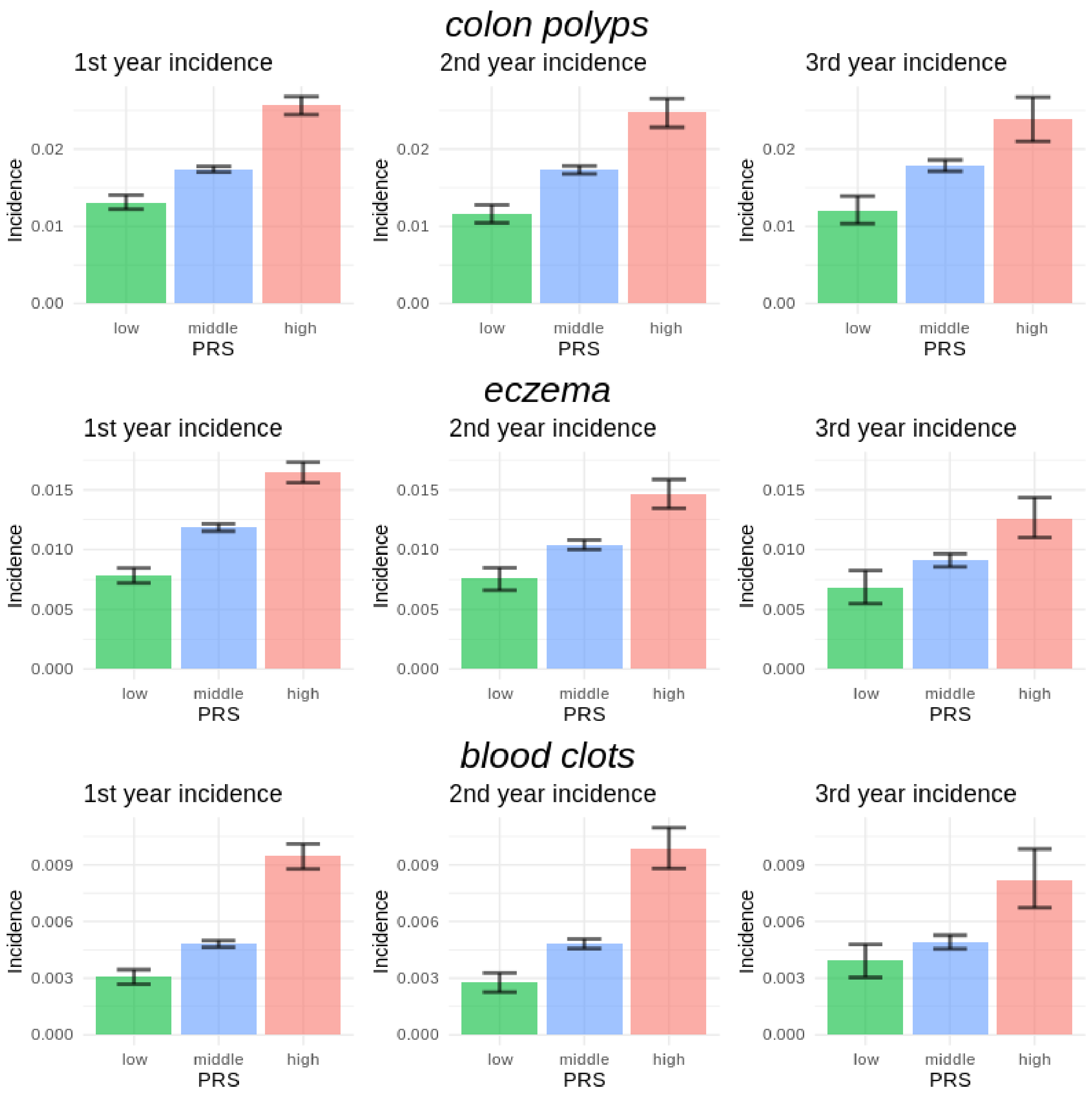


**Fig S4.D Incidence of endpoints by strata defined by PRS during 3 years follow-up**

Each plot illustrates the incidence over the 1st, 2nd, and 3rd year of follow-up among individuals grouped by each disease-specific PRS. Individuals in the top 10% of each PRS are represented by a red bar, those in the middle 80% by a blue bar and those in the bottom 10% by a green bar. The error bars indicate the 95% CI from bootstrapping.

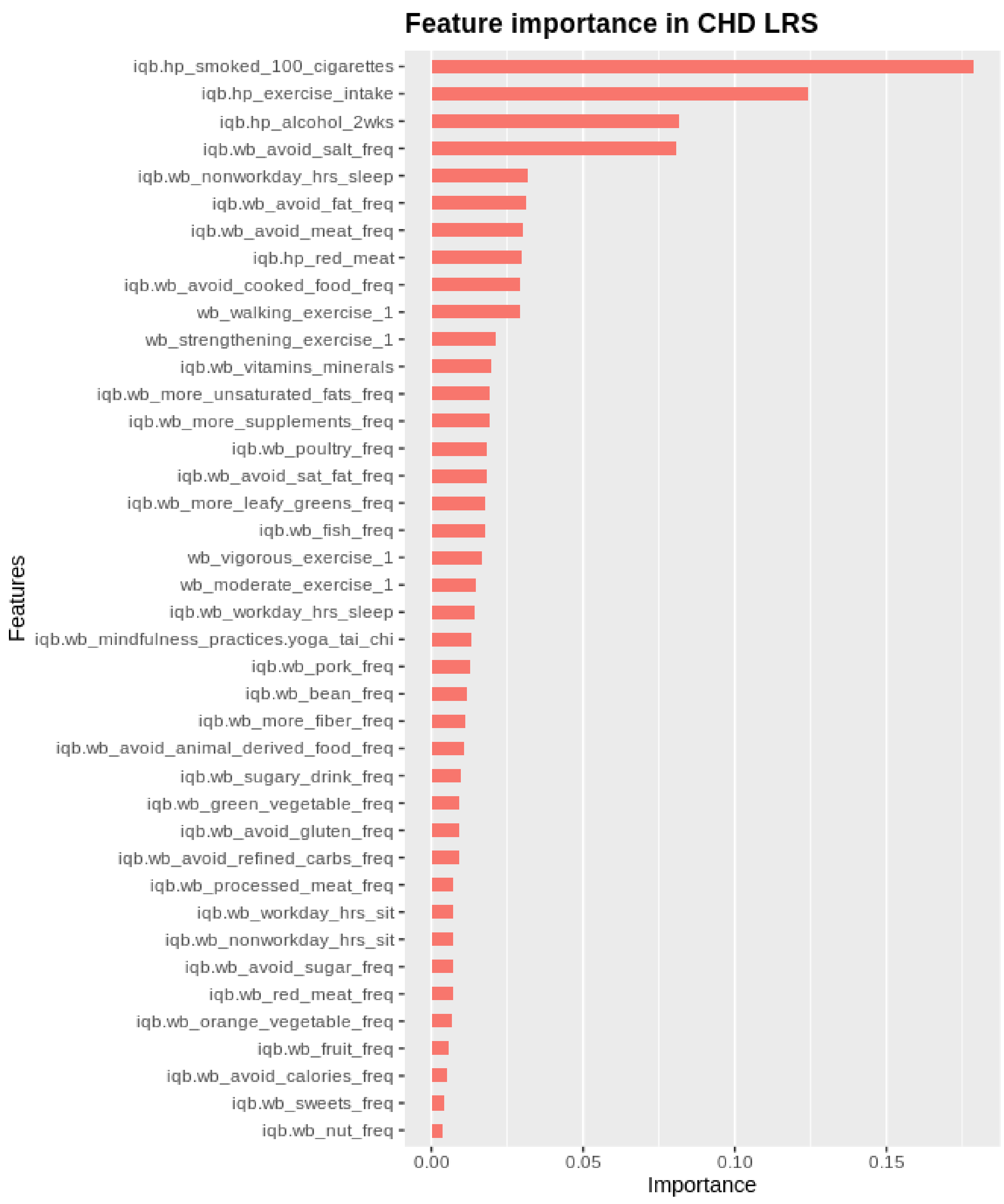


**Fig S5.A Feature importance plot from CHD LRS model.**

Each bar indicates the feature importance from the XGboost model for CHD LRS, and “Gain” was used as the measure of the feature importance.

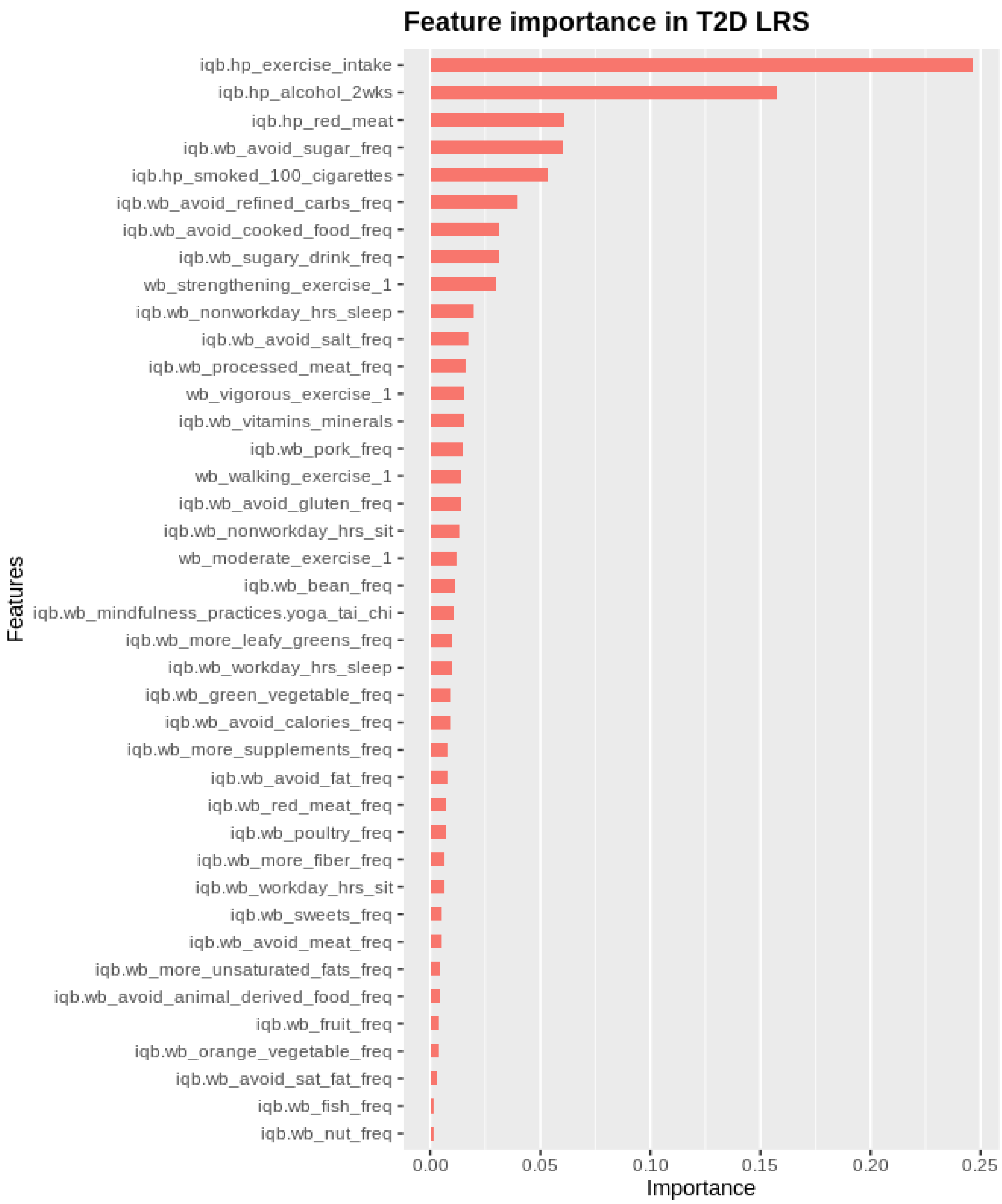


**Fig S5.B Feature importance plot from T2D LRS model.**

Each bar indicates the feature importance from the XGboost model for T2D LRS, and “Gain” was used as the measure of the feature importance.

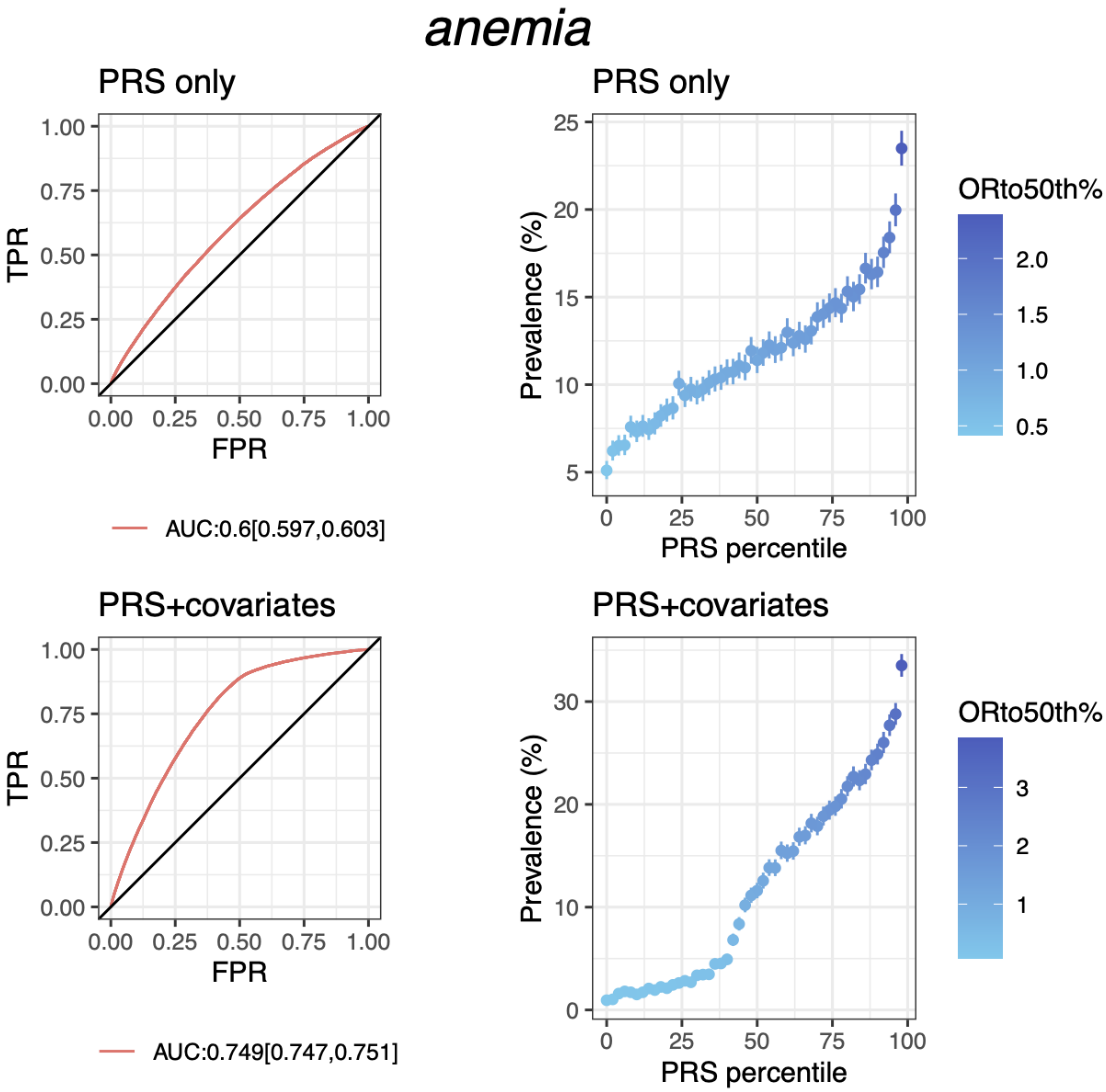


**Fig S6.A PRS performance for anemia in testing data from discovery dataset (European population)**

Plots on the left column show the receiver operating characteristics curve and AUC with 95% CI; plots on the right column show the disease prevalence in groups binned according to PRS (or PRS+covariates) percentile, where ORto50th% shows the odd ratio of each bin over the 25th bin (50 bins in total). PRS+covariates is the model with PRS, age, sex and PCs.

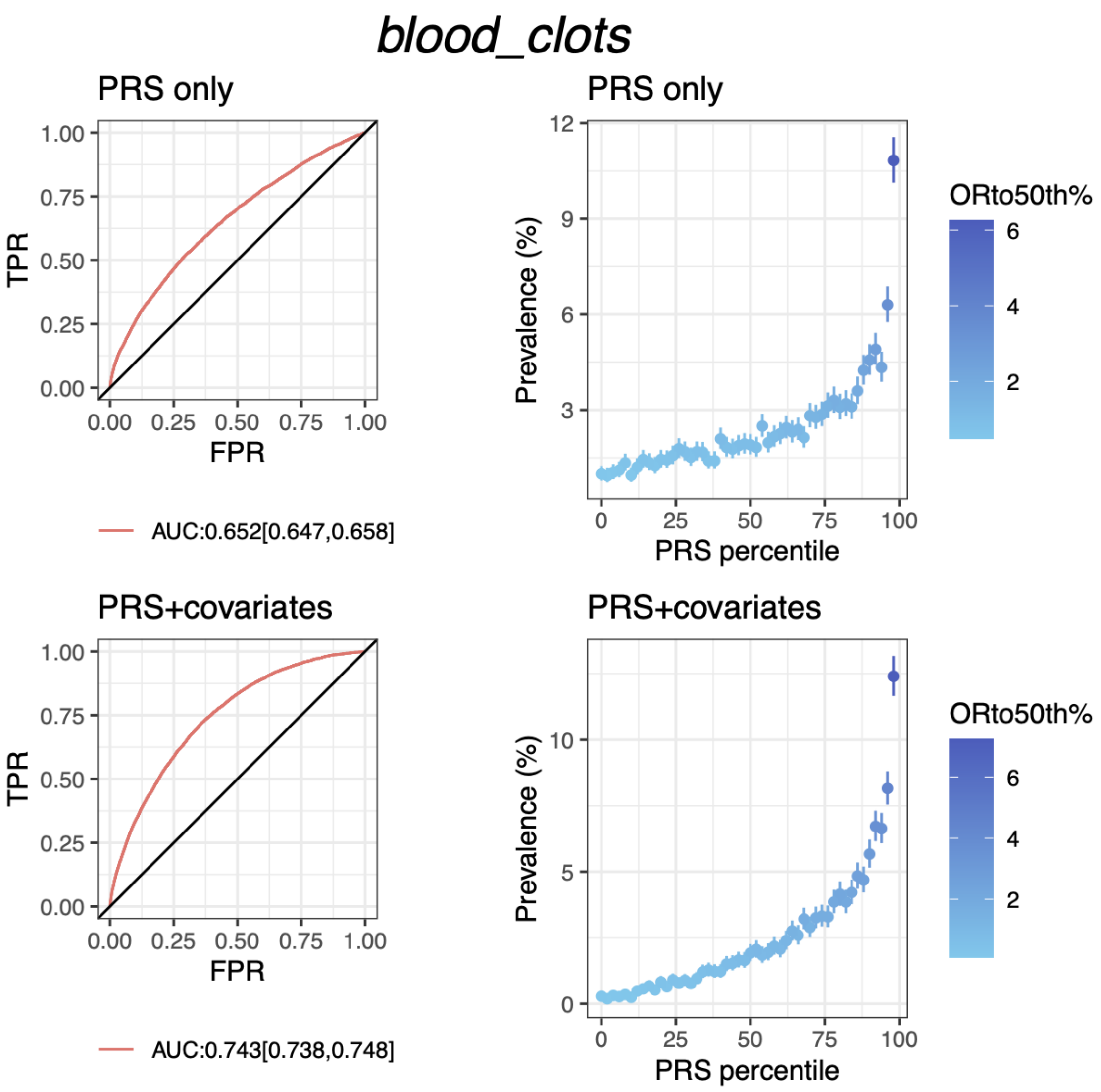


**Fig S6.B PRS performance for blood clots in testing data from discovery dataset (European population)**

Plots on the left column show the receiver operating characteristics curve and AUC with 95% CI; plots on the right column show the disease prevalence in groups binned according to PRS (or PRS+covariates) percentile, where ORto50th% shows the odd ratio of each bin over the 25th bin (50 bins in total). PRS+covariates is the model with PRS, age, sex and PCs.

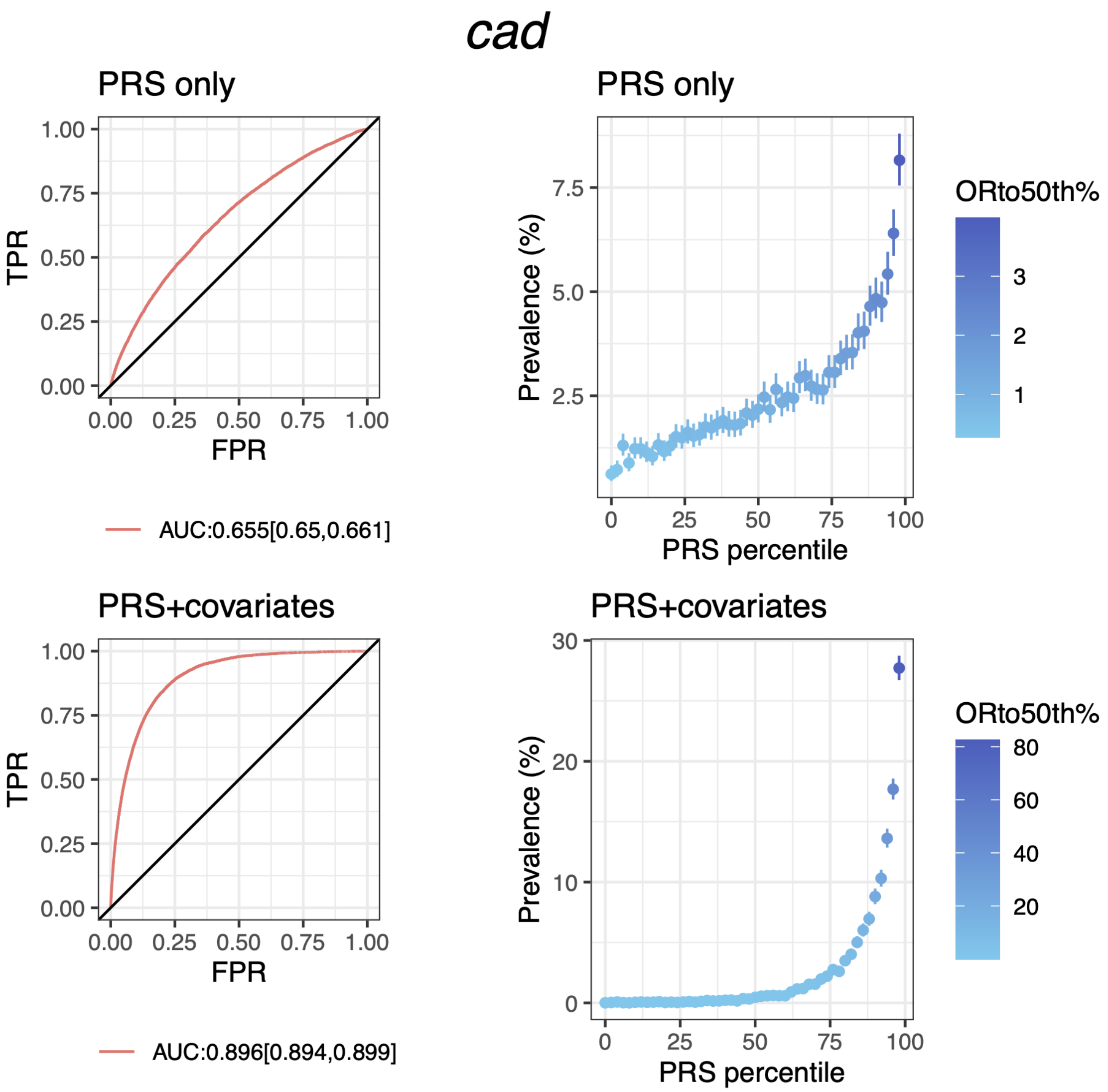


**Fig S6.C PRS performance for coronary heart disease in testing data from discovery dataset (European population)**

Plots on the left column show the receiver operating characteristics curve and AUC with 95% CI; plots on the right column show the disease prevalence in groups binned according to PRS (or PRS+covariates) percentile, where ORto50th% shows the odd ratio of each bin over the 25th bin (50 bins in total). PRS+covariates is the model with PRS, age, sex and PCs.

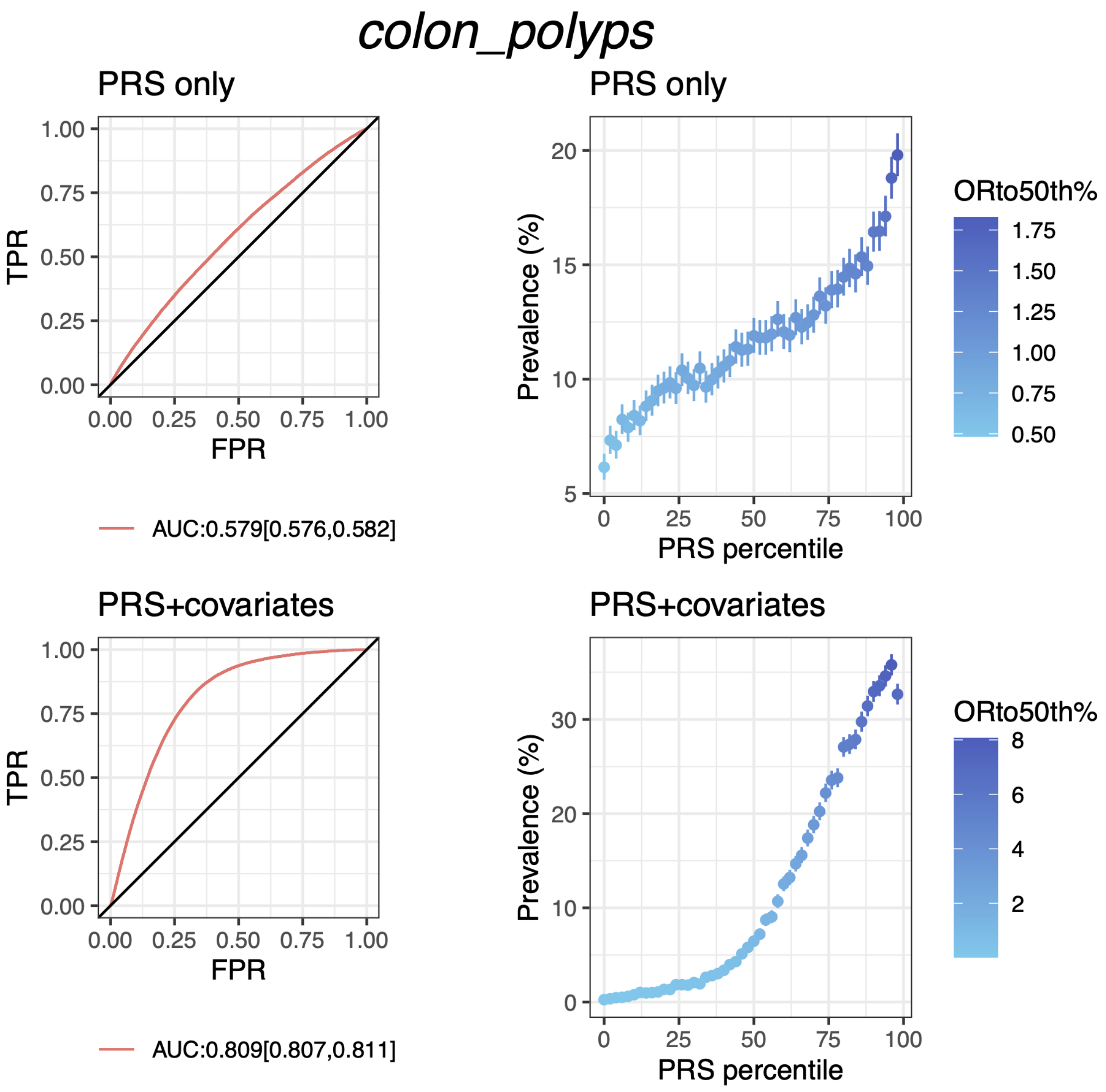


**Fig S6.D PRS performance for colon polyps in testing data from discovery dataset (European population)**

Plots on the left column show the receiver operating characteristics curve and AUC with 95% CI; plots on the right column show the disease prevalence in groups binned according to PRS (or PRS+covariates) percentile, where ORto50th% shows the odd ratio of each bin over the 25th bin (50 bins in total). PRS+covariates is the model with PRS, age, sex and PCs.

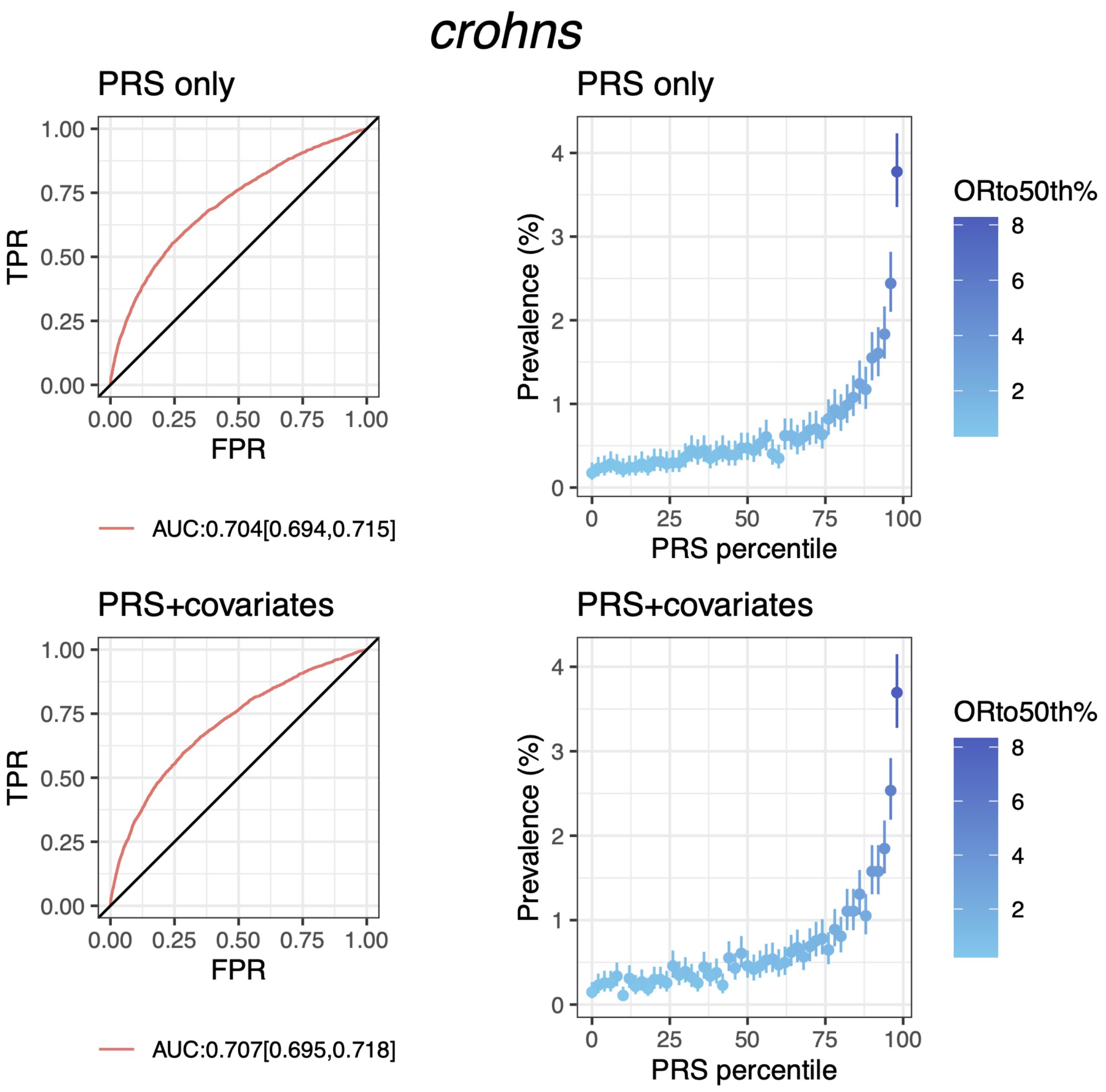


**Fig S6.E PRS performance for Crohn's in testing data from discovery dataset (European population)**

Plots on the left column show the receiver operating characteristics curve and AUC with 95% CI; plots on the right column show the disease prevalence in groups binned according to PRS (or PRS+covariates) percentile, where ORto50th% shows the odd ratio of each bin over the 25th bin (50 bins in total). PRS+covariates is the model with PRS, age, sex and PCs.

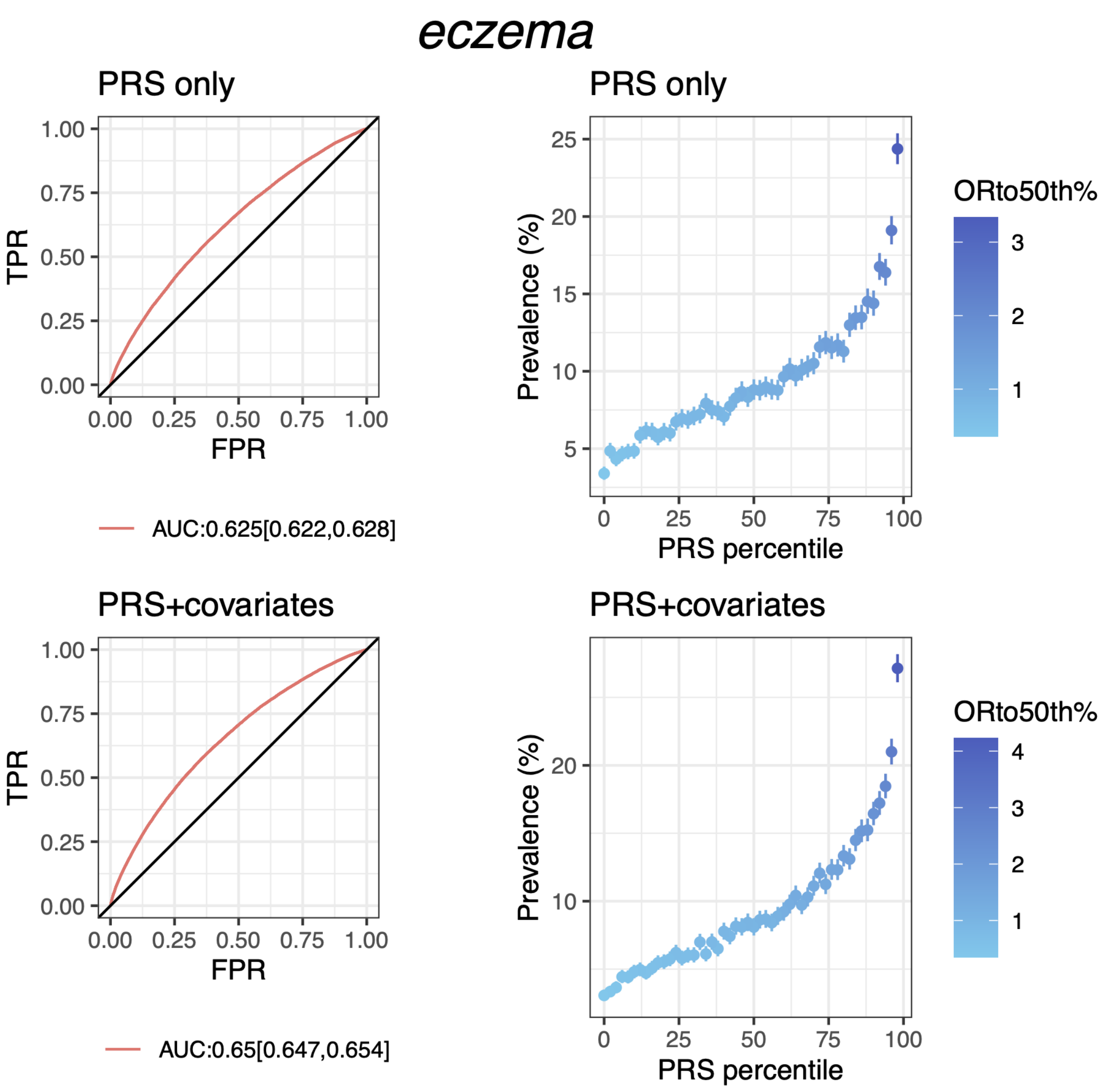


**Fig S6.F PRS performance for eczema in testing data from discovery dataset (European population)**

Plots on the left column show the receiver operating characteristics curve and AUC with 95% CI; plots on the right column show the disease prevalence in groups binned according to PRS (or PRS+covariates) percentile, where ORto50th% shows the odd ratio of each bin over the 25th bin (50 bins in total). PRS+covariates is the model with PRS, age, sex and PCs.

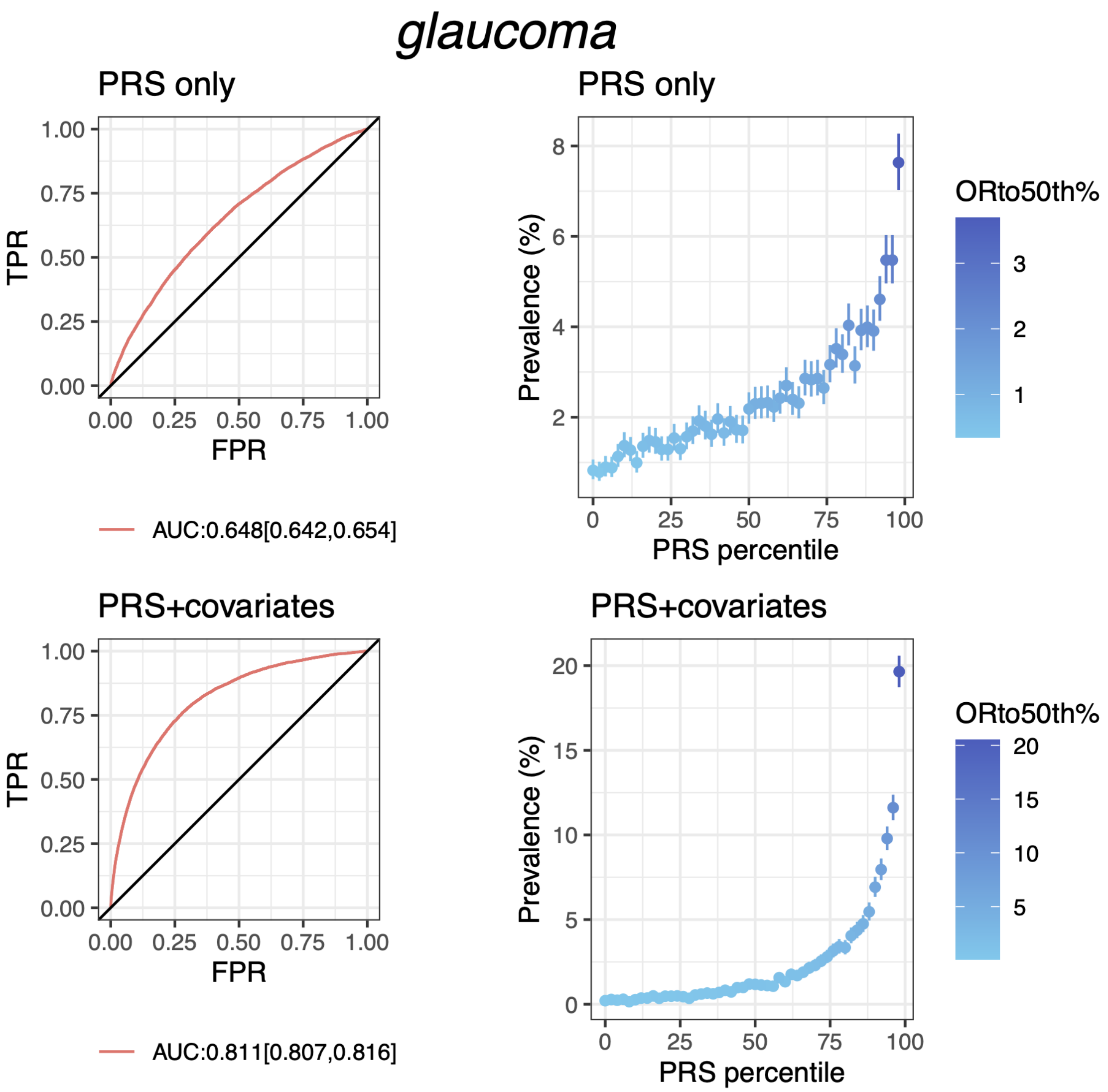


**Fig S6.G PRS performance for glaucoma in testing data from discovery dataset (European population)**

Plots on the left column show the receiver operating characteristics curve and AUC with 95% CI; plots on the right column show the disease prevalence in groups binned according to PRS (or PRS+covariates) percentile, where ORto50th% shows the odd ratio of each bin over the 25th bin (50 bins in total). PRS+covariates is the model with PRS, age, sex and PCs.

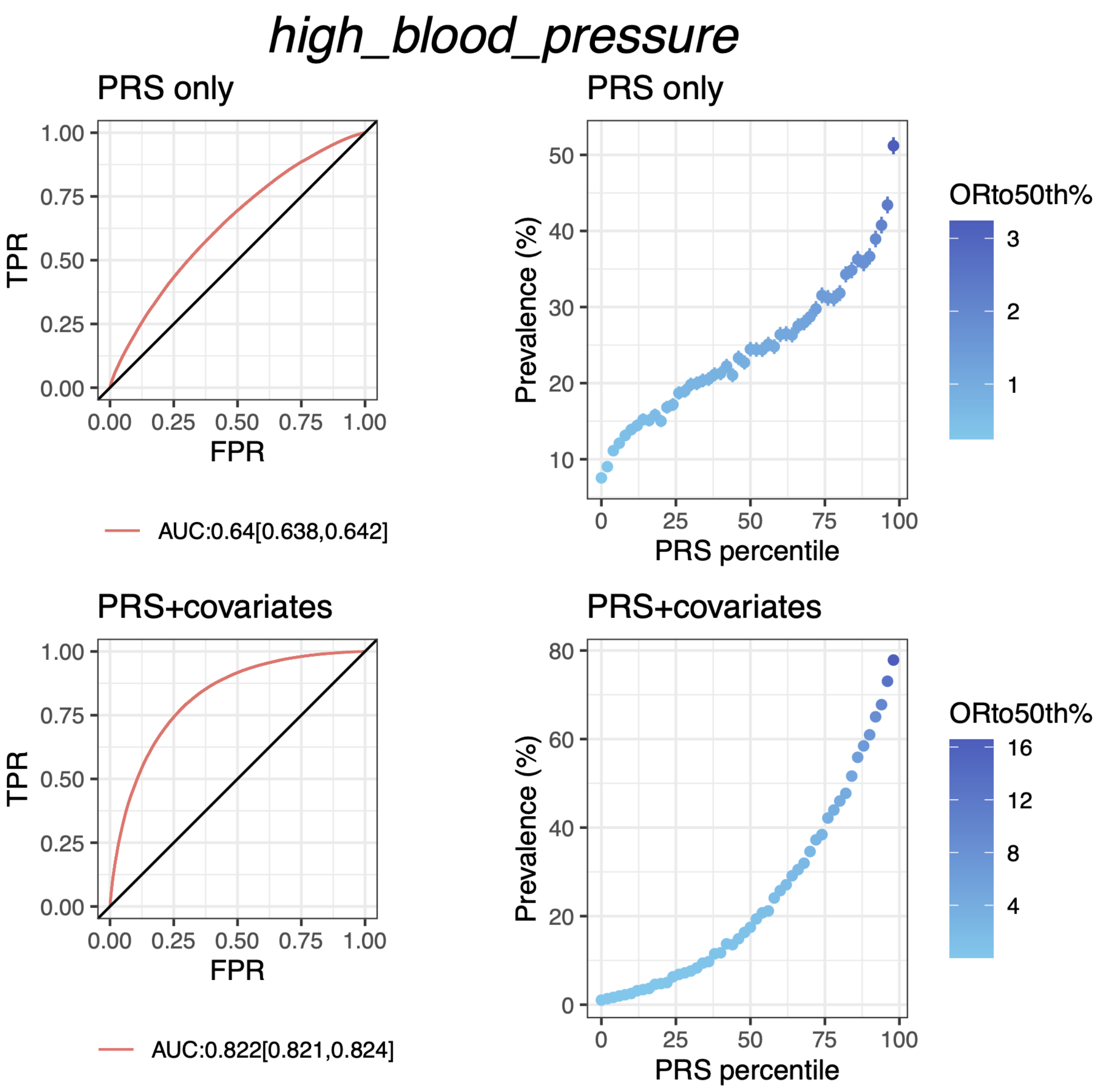


**Fig S6.H PRS performance for high blood pressure in testing data from discovery dataset (European population)**

Plots on the left column show the receiver operating characteristics curve and AUC with 95% CI; plots on the right column show the disease prevalence in groups binned according to PRS (or PRS+covariates) percentile, where ORto50th% shows the odd ratio of each bin over the 25th bin (50 bins in total). PRS+covariates is the model with PRS, age, sex and PCs.

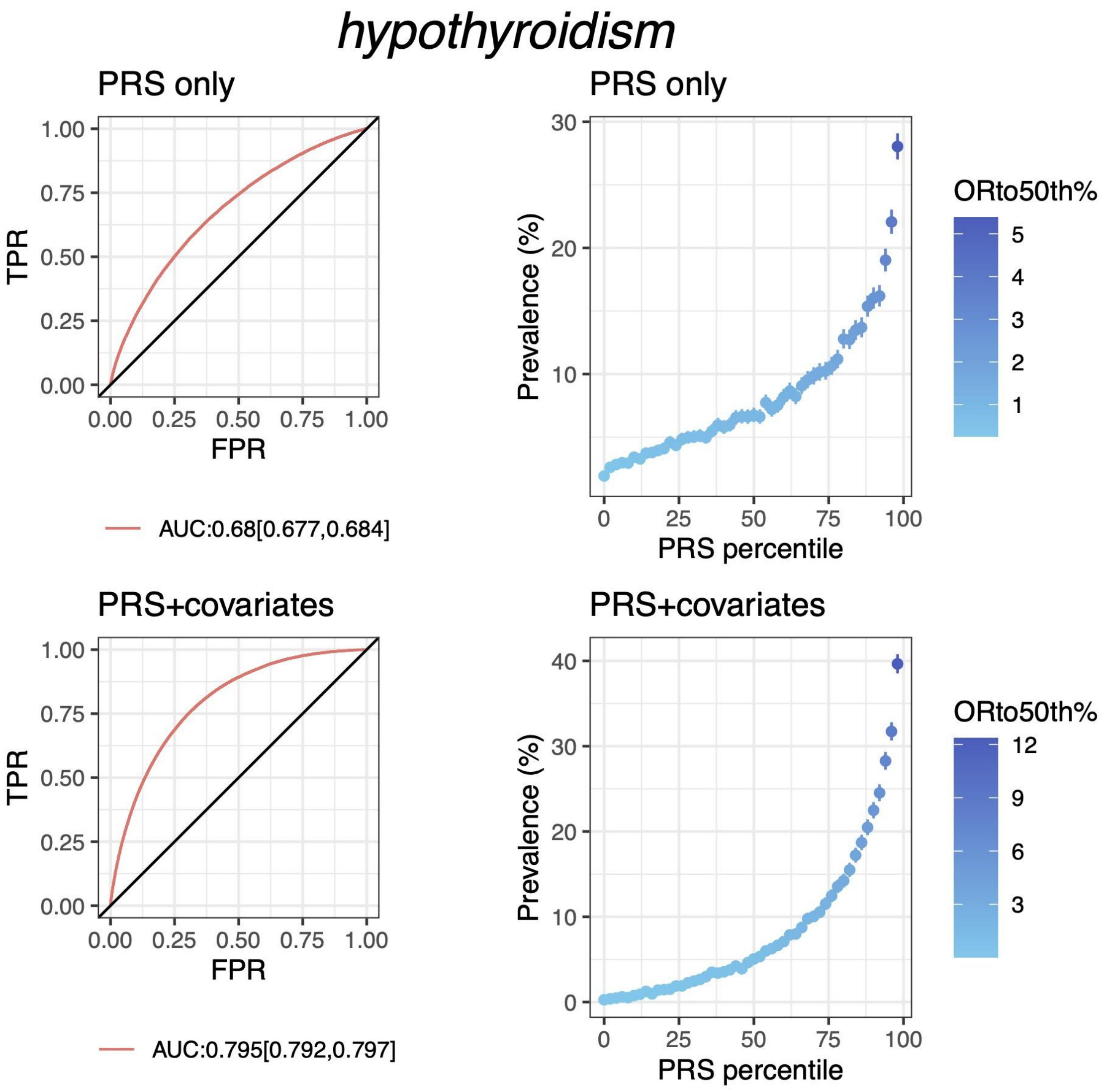


**Fig S6.I PRS performance for hypothyroidism in testing data from discovery dataset (European population)**

Plots on the left column show the receiver operating characteristics curve and AUC with 95% CI; plots on the right column show the disease prevalence in groups binned according to PRS (or PRS+covariates) percentile, where ORto50th% shows the odd ratio of each bin over the 25th bin (50 bins in total). PRS+covariates is the model with PRS, age, sex and PCs.

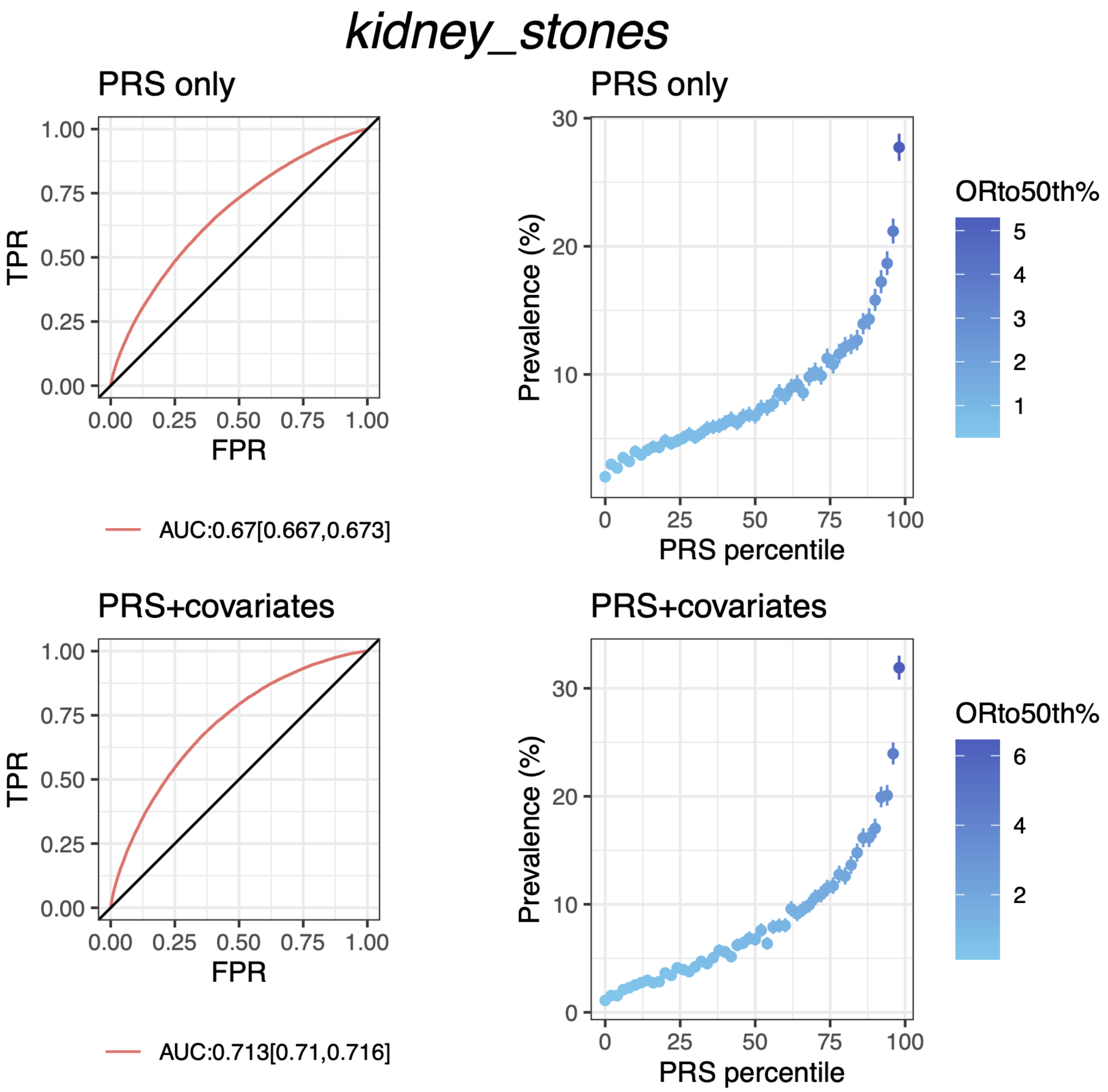


**Fig S6.J PRS performance for kidney stones in testing data from discovery dataset (European population)**

Plots on the left column show the receiver operating characteristics curve and AUC with 95% CI; plots on the right column show the disease prevalence in groups binned according to PRS (or PRS+covariates) percentile, where ORto50th% shows the odd ratio of each bin over the 25th bin (50 bins in total). PRS+covariates is the model with PRS, age, sex and PCs.

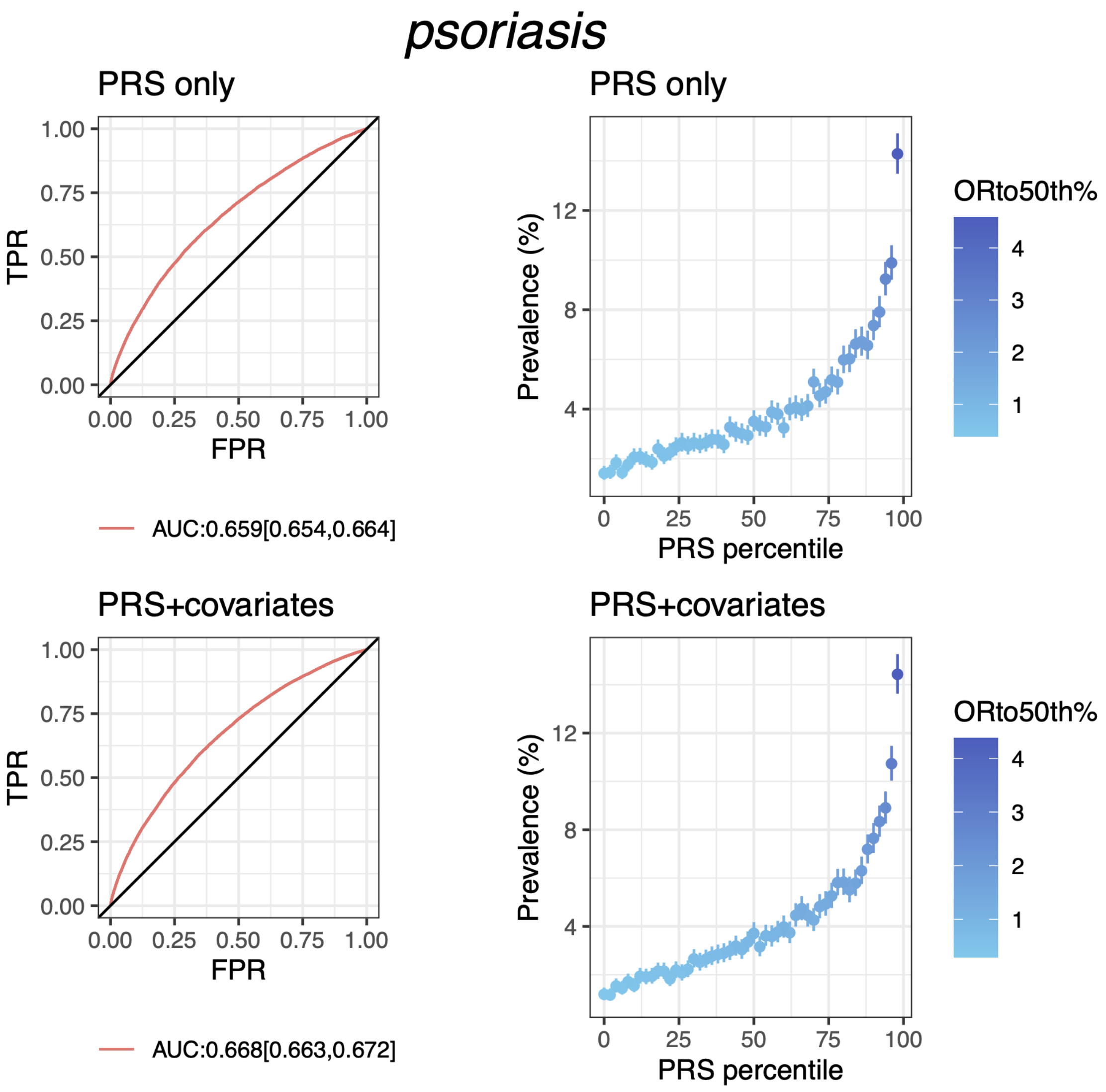


**Fig S6.K PRS performance for psoriasis in testing data from discovery dataset (European population)**

Plots on the left column show the receiver operating characteristics curve and AUC with 95% CI; plots on the right column show the disease prevalence in groups binned according to PRS (or PRS+covariates) percentile, where ORto50th% shows the odd ratio of each bin over the 25th bin (50 bins in total). PRS+covariates is the model with PRS, age, sex and PCs.

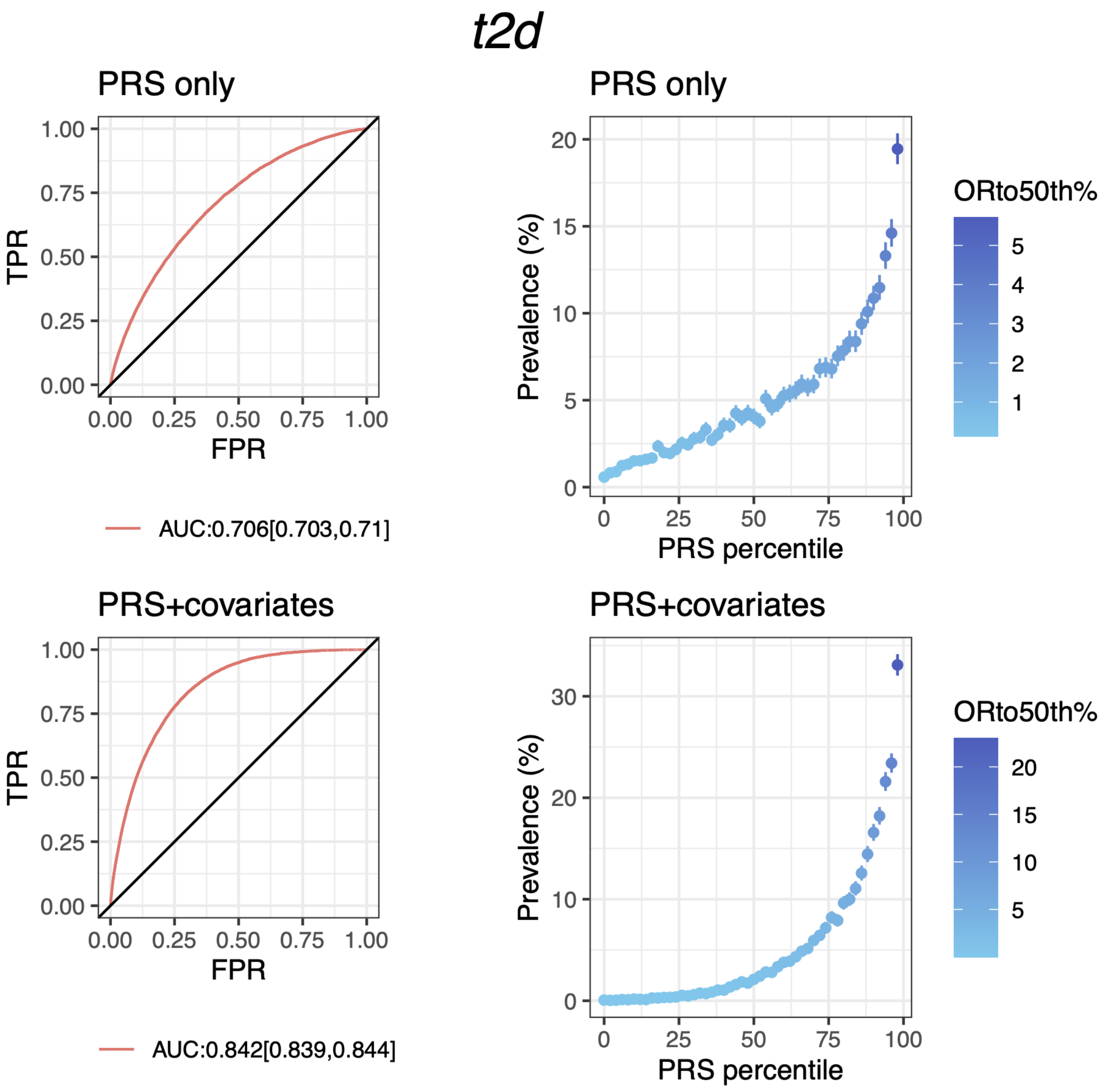


**Fig S6.L PRS performance for type 2 diabetes in testing data from discovery dataset (European population)**

Plots on the left column show the receiver operating characteristics curve and AUC with 95% CI; plots on the right column show the disease prevalence in groups binned according to PRS (or PRS+covariates) percentile, where ORto50th% shows the odd ratio of each bin over the 25th bin (50 bins in total). PRS+covariates is the model with PRS, age, sex and PCs.

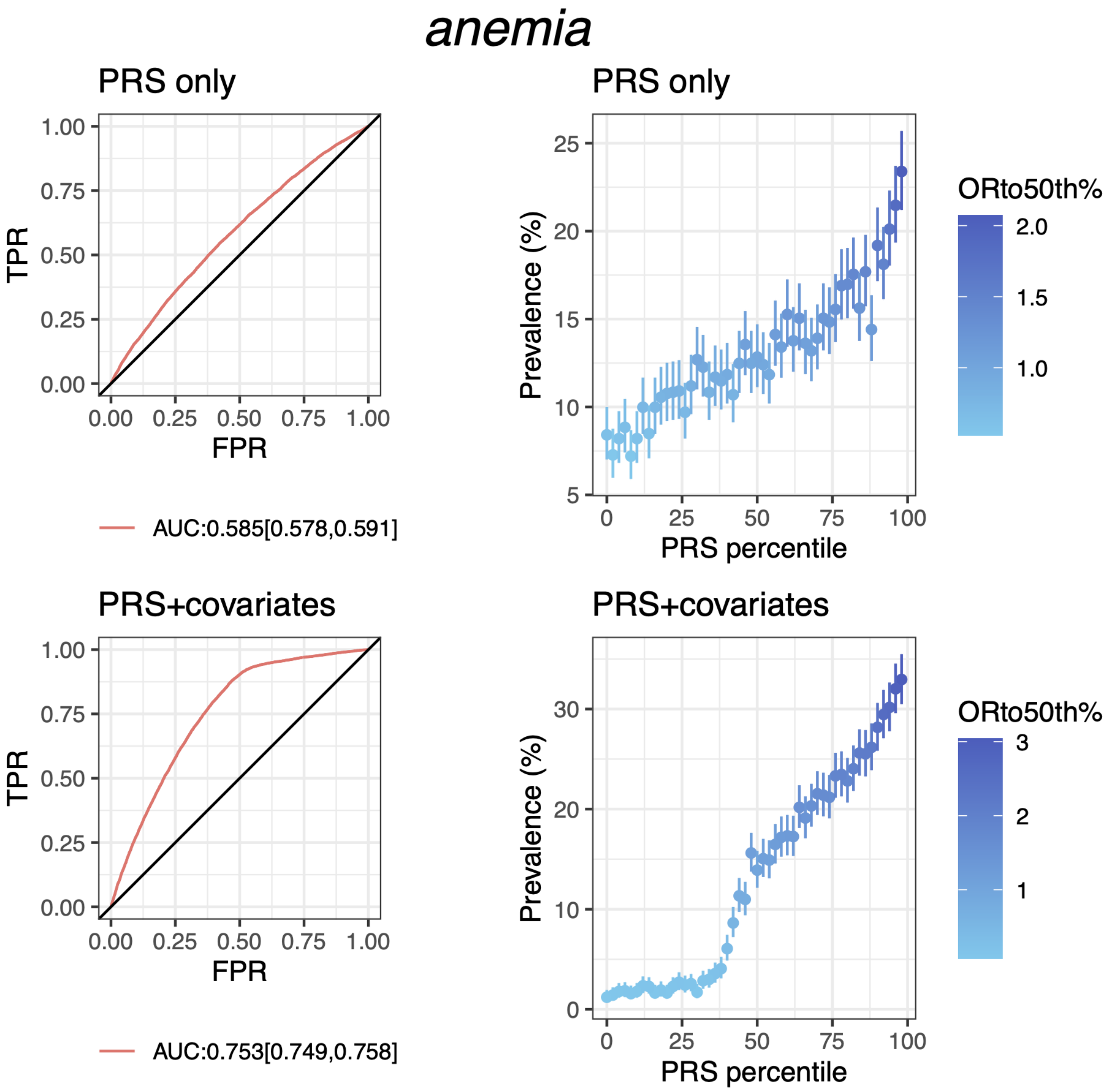


**Fig S7.A PRS performance for anemia in testing data from discovery dataset (Latine population)**

Plots on the left column show the receiver operating characteristics curve and AUC with 95% CI; plots on the right column show the disease prevalence in groups binned according to PRS (or PRS+covariates) percentile, where ORto50th% shows the odd ratio of each bin over the 25th bin (50 bins in total). PRS+covariates is the model with PRS, age, sex and PCs.

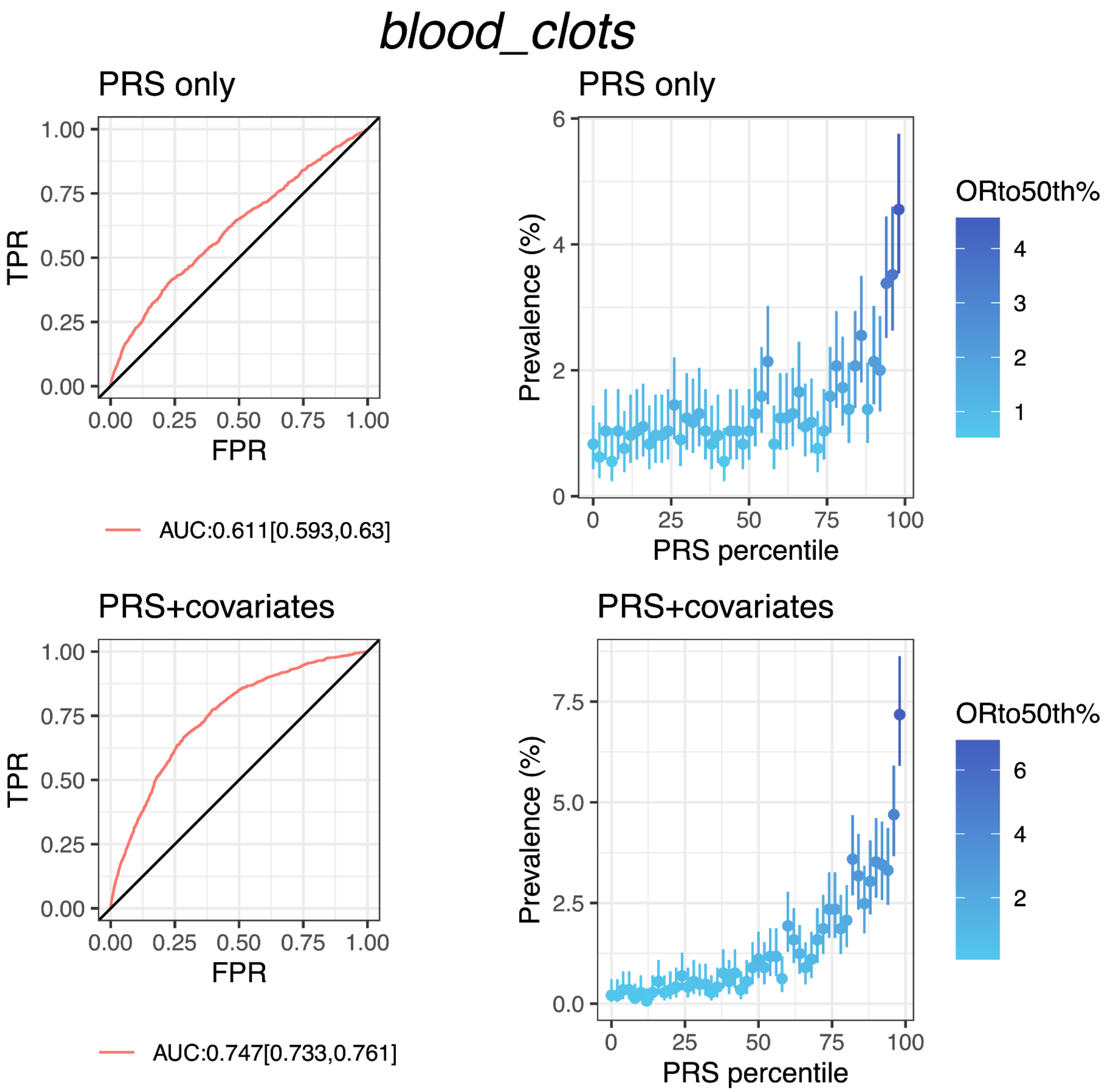


**Fig S7.B PRS performance for blood clots in testing data from discovery dataset (Latine population)**

Plots on the left column show the receiver operating characteristics curve and AUC with 95% CI; plots on the right column show the disease prevalence in groups binned according to PRS (or PRS+covariates) percentile, where ORtoth50% shows the odd ratio of each bin over the 25th bin (50 bins in total). PRS+covariates is the model with PRS, age, sex and PCs.

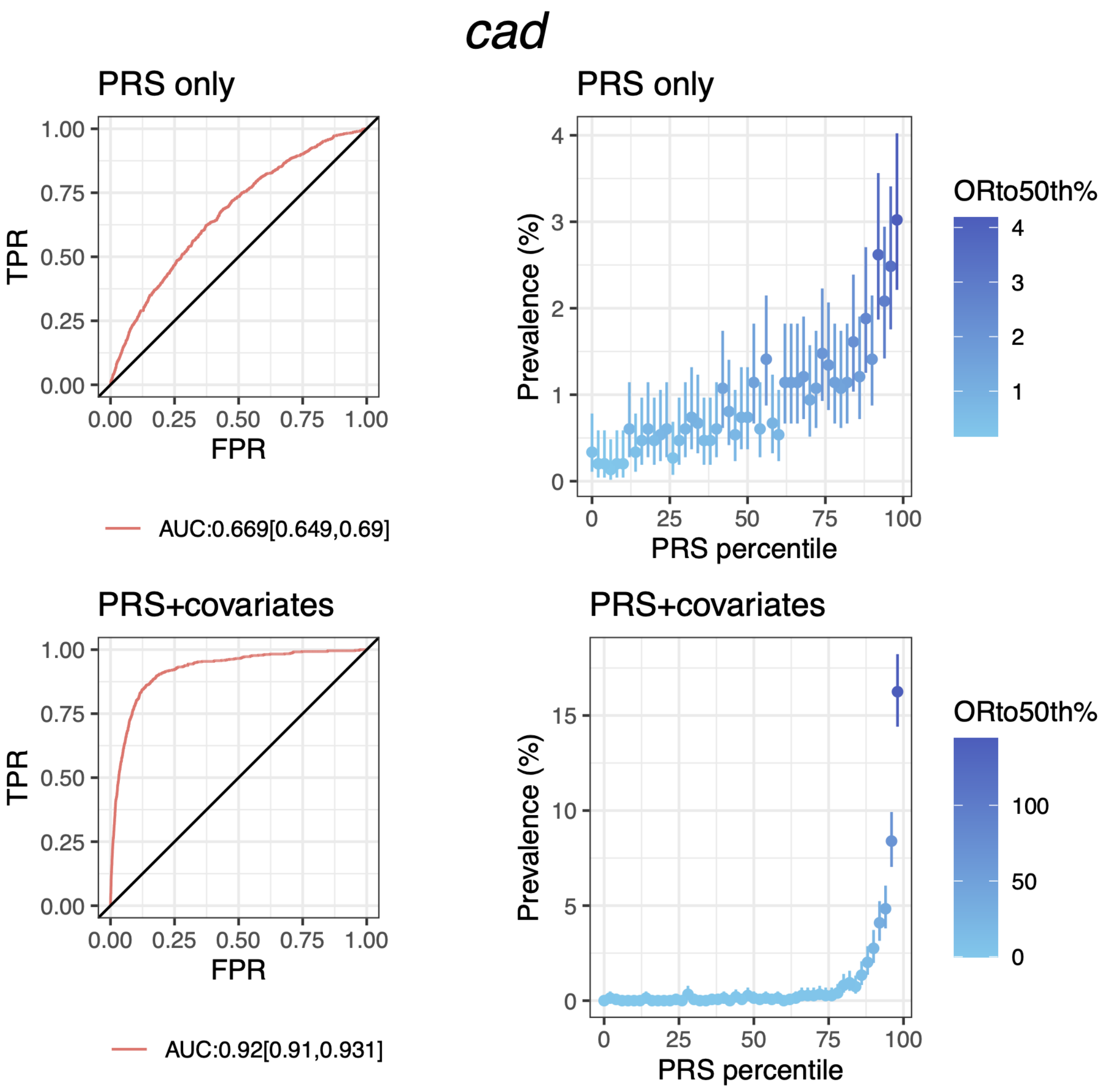


**Fig S7.C PRS performance for coronary heart disease in testing data from discovery dataset (Latine population)**

Plots on the left column show the receiver operating characteristics curve and AUC with 95% CI; plots on the right column show the disease prevalence in groups binned according to PRS (or PRS+covariates) percentile, where ORto50th% shows the odd ratio of each bin over the 25th bin (50 bins in total). PRS+covariates is the model with PRS, age, sex and PCs.

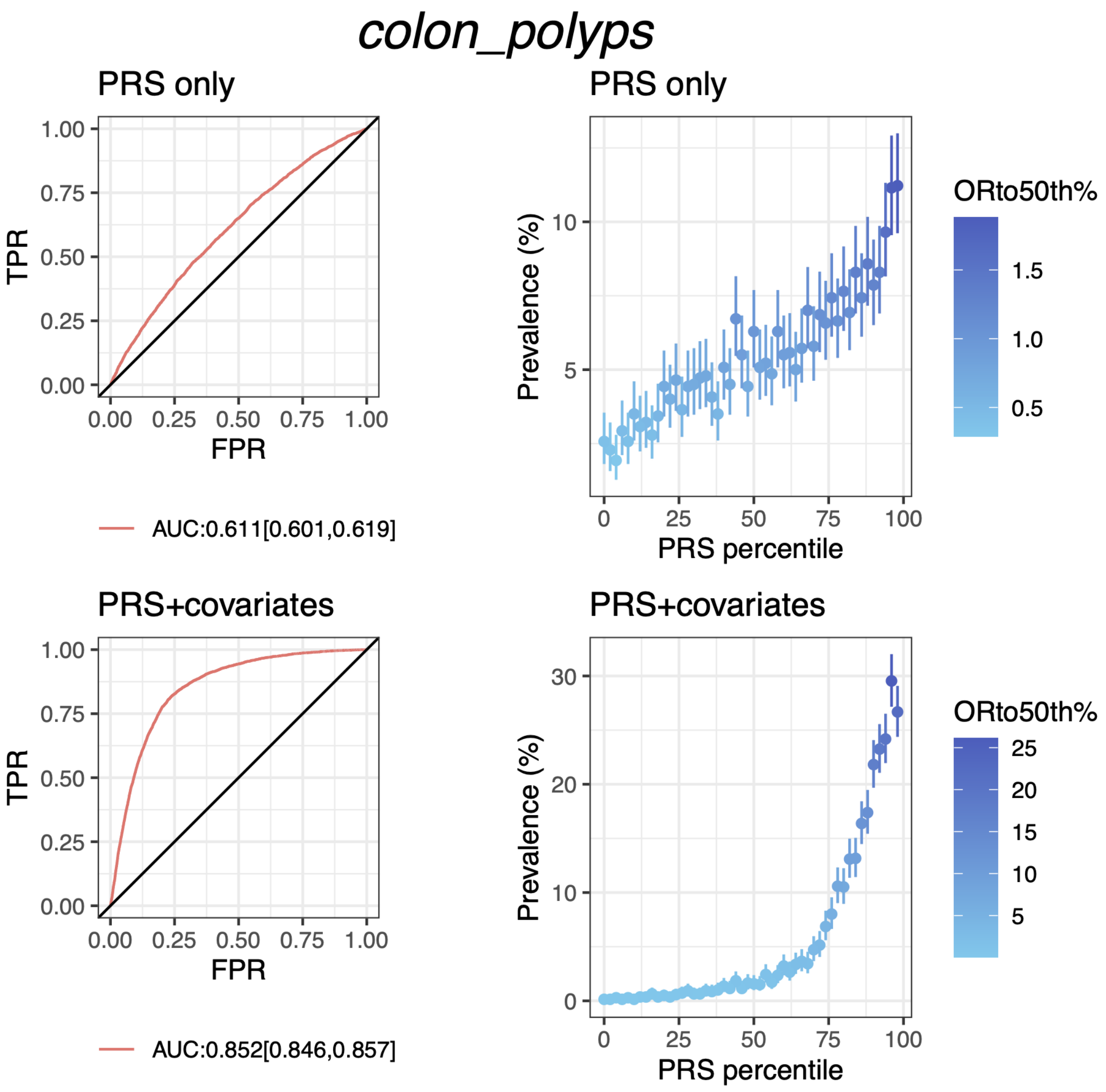


**Fig S7.D PRS performance for colon polyps in testing data from discovery dataset (Latine population)**

Plots on the left column show the receiver operating characteristics curve and AUC with 95% CI; plots on the right column show the disease prevalence in groups binned according to PRS (or PRS+covariates) percentile, where ORto50th% shows the odd ratio of each bin over the 25th bin (50 bins in total). PRS+covariates is the model with PRS, age, sex and PCs.

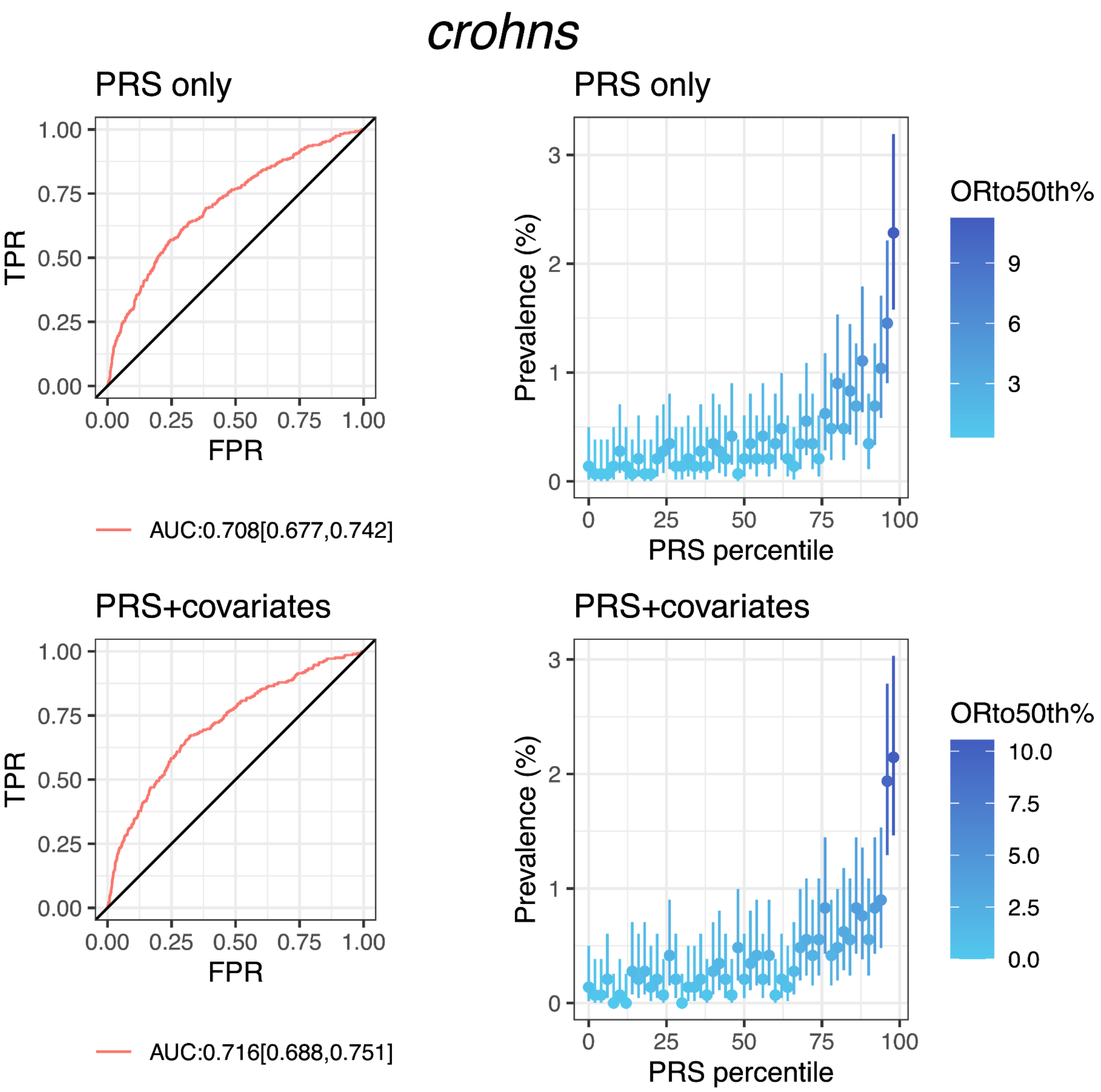


**Fig S7.E PRS performance for Crohn's in testing data from discovery dataset (Latine population)**

Plots on the left column show the receiver operating characteristics curve and AUC with 95% CI; plots on the right column show the disease prevalence in groups binned according to PRS (or PRS+covariates) percentile, where ORto50th% shows the odd ratio of each bin over the 25th bin (50 bins in total). PRS+covariates is the model with PRS, age, sex and PCs.

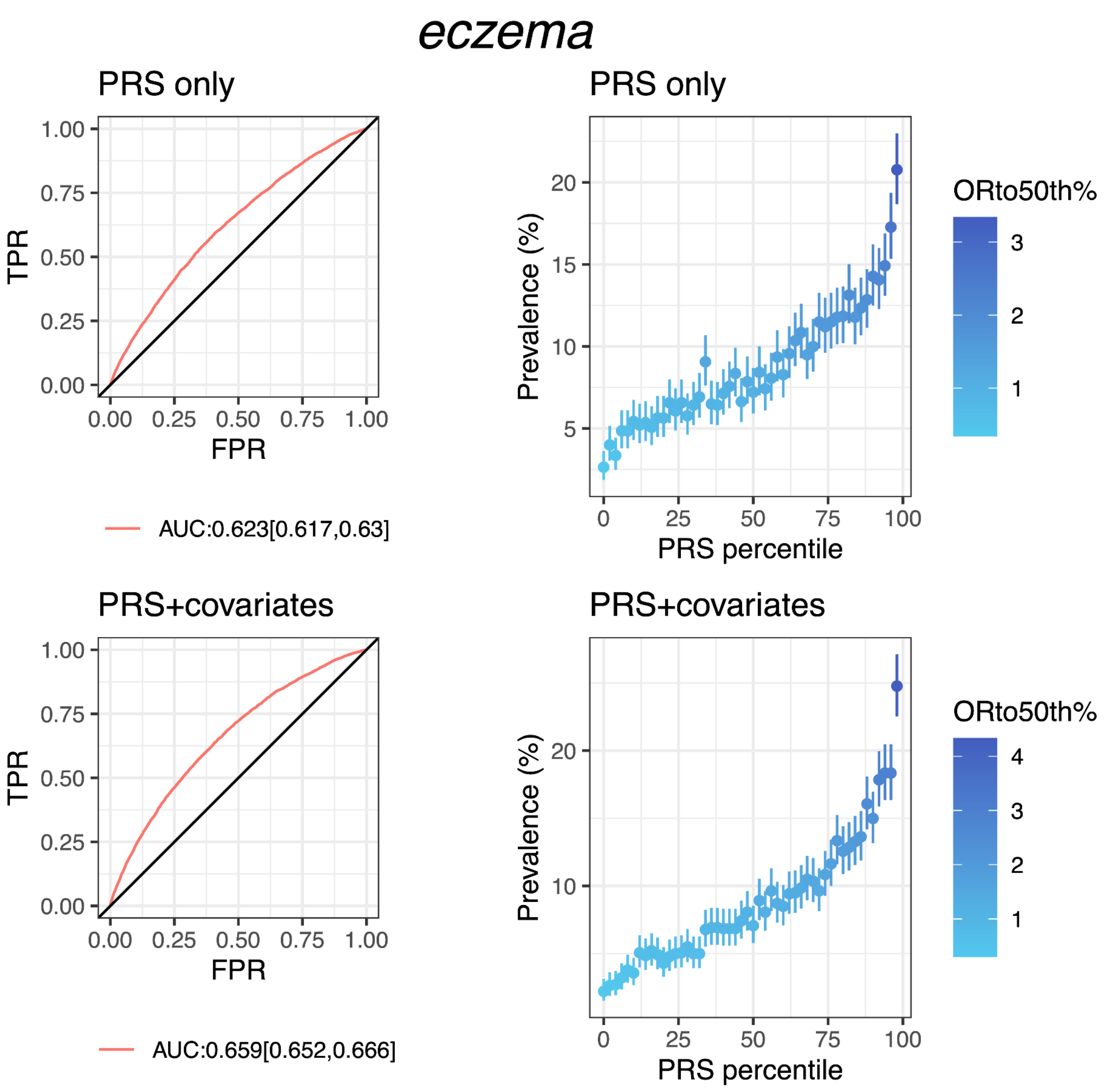


**Fig S7.F PRS performance for eczema in testing data from discovery dataset (Latine population)**

Plots on the left column show the receiver operating characteristics curve and AUC with 95% CI; plots on the right column show the disease prevalence in groups binned according to PRS (or PRS+covariates) percentile, where ORto50th% shows the odd ratio of each bin over the 25th bin (50 bins in total). PRS+covariates is the model with PRS, age, sex and PCs.

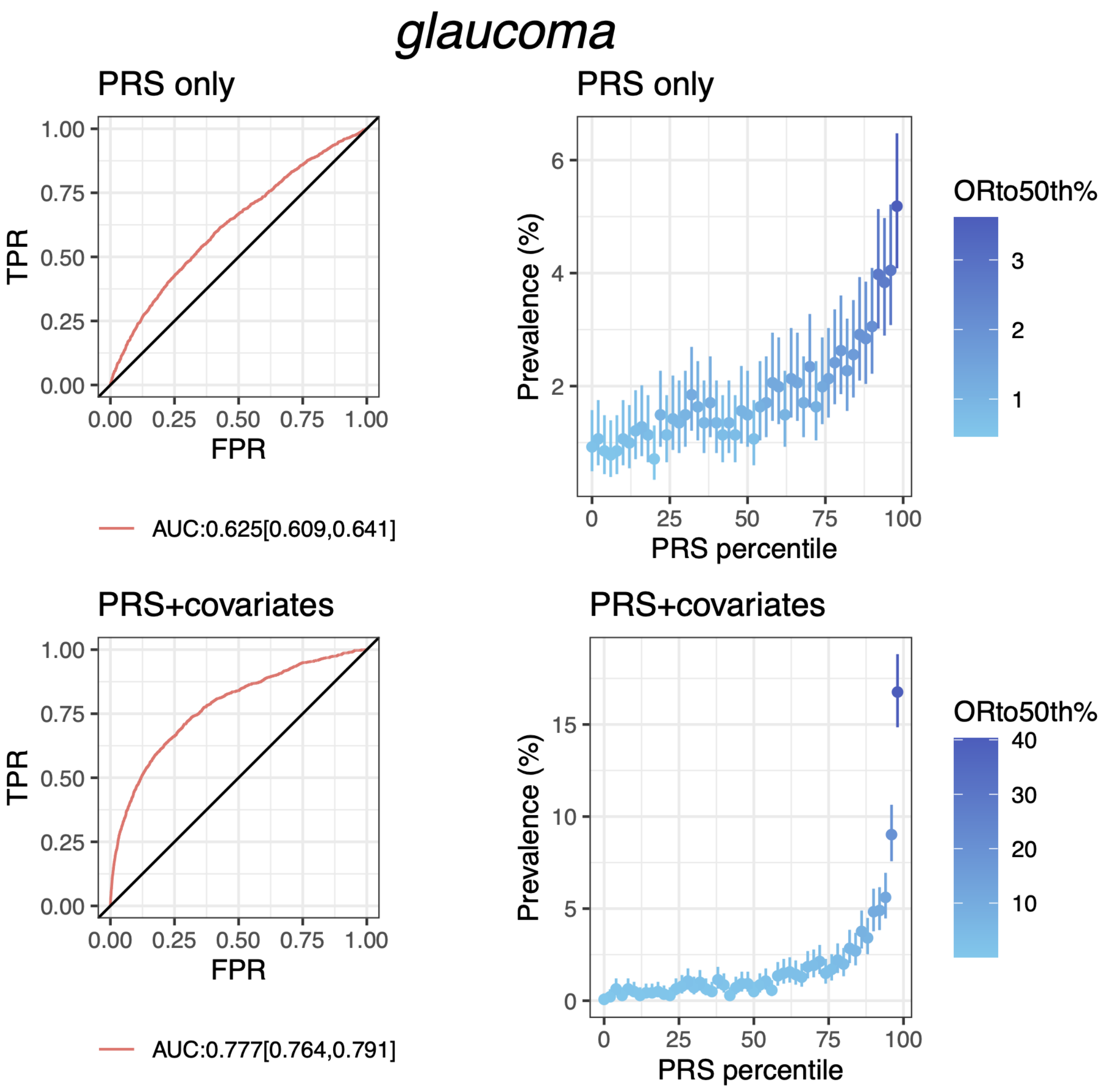


**Fig S7.G PRS performance for glaucoma in testing data from discovery dataset (Latine population)**

Plots on the left column show the receiver operating characteristics curve and AUC with 95% CI; plots on the right column show the disease prevalence in groups binned according to PRS (or PRS+covariates) percentile, where ORto50th% shows the odd ratio of each bin over the 25th bin (50 bins in total). PRS+covariates is the model with PRS, age, sex and PCs.

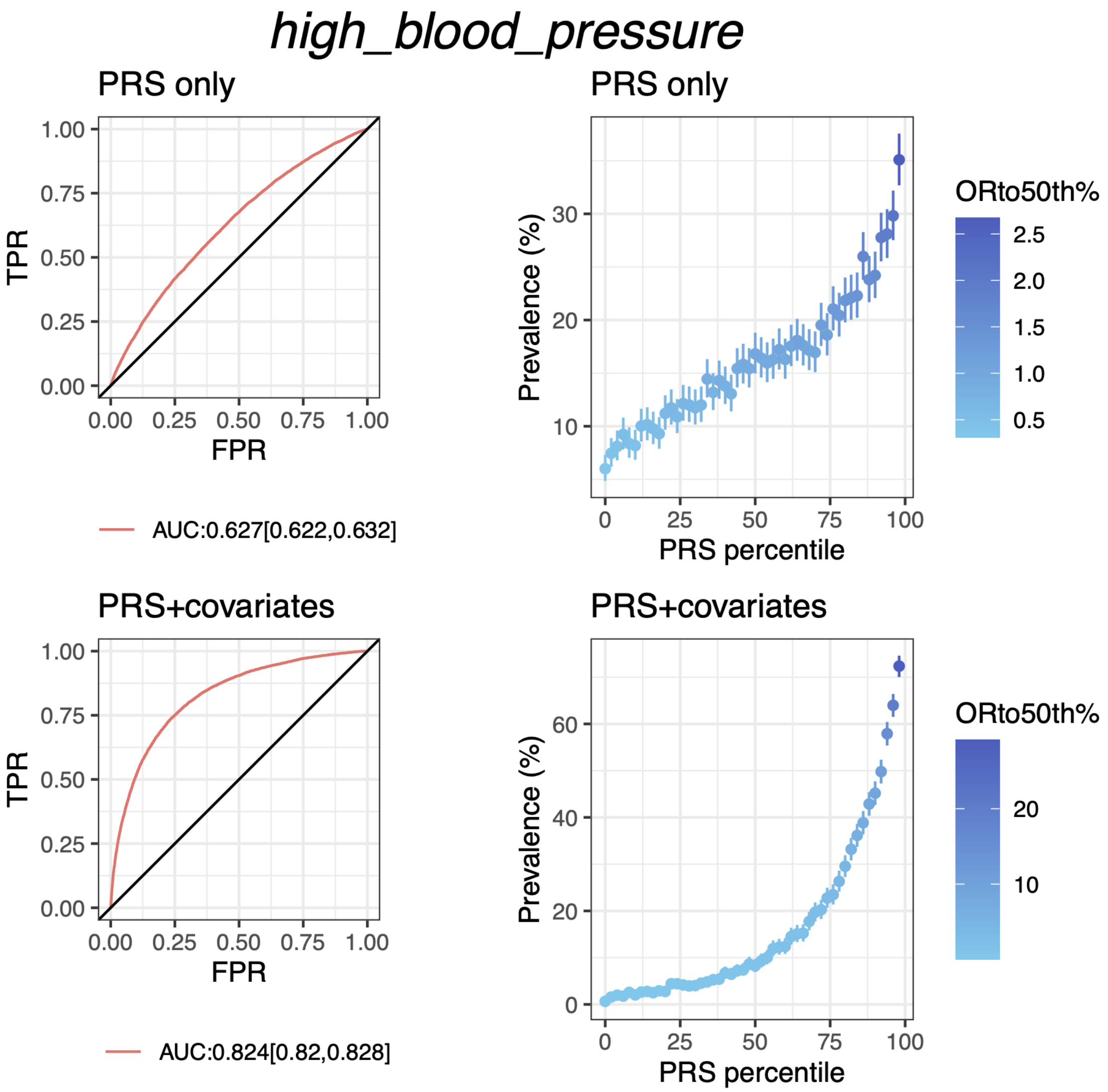


**Fig S7.H PRS performance for high blood pressure in testing data from discovery dataset (Latine population)**

Plots on the left column show the receiver operating characteristics curve and AUC with 95% CI; plots on the right column show the disease prevalence in groups binned according to PRS (or PRS+covariates) percentile, where ORto50th% shows the odd ratio of each bin over the 25th bin (50 bins in total). PRS+covariates is the model with PRS, age, sex and PCs.

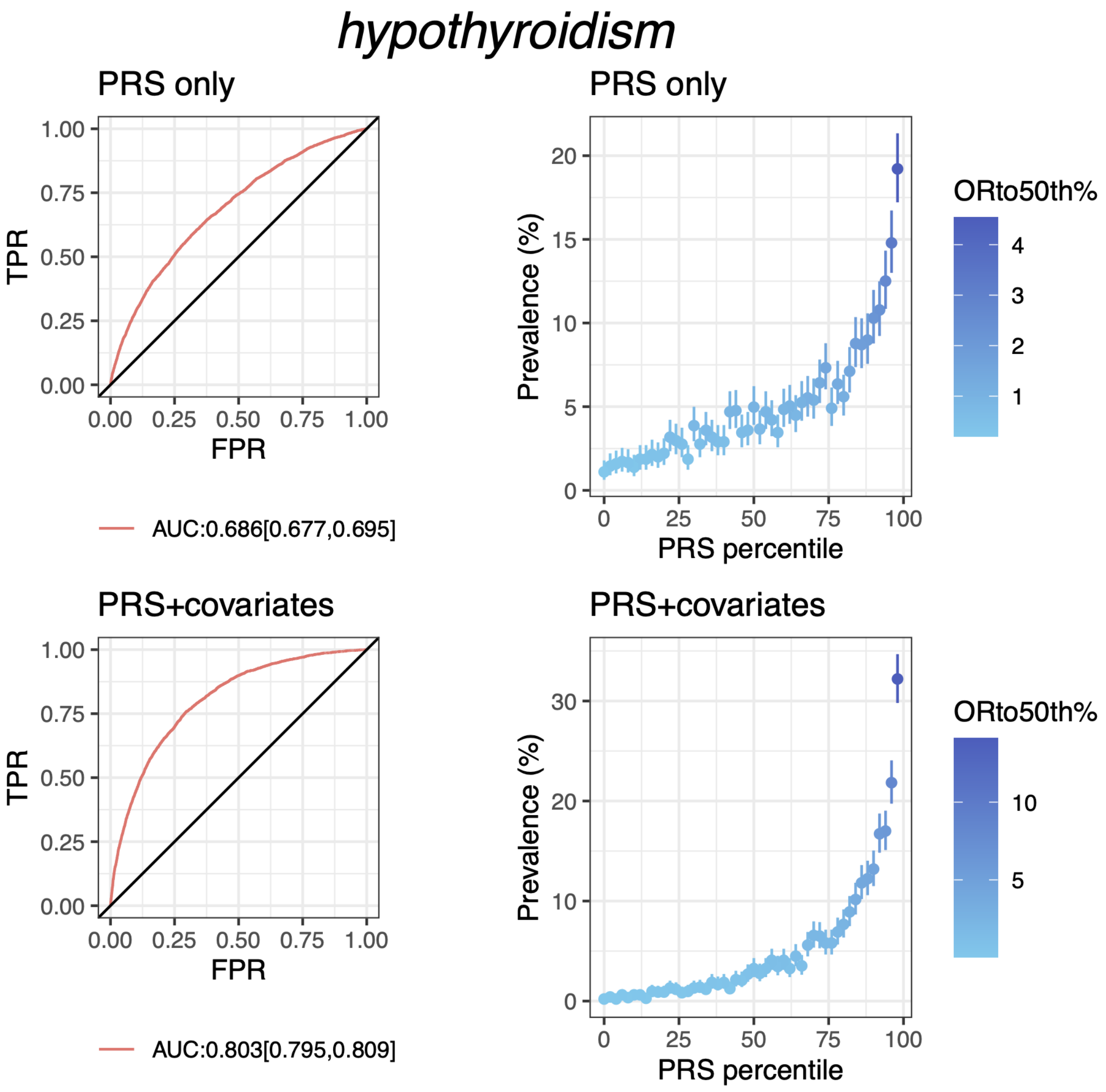


**Fig S7.I PRS performance for hypothyroidism in testing data from discovery dataset (Latine population)**

Plots on the left column show the receiver operating characteristics curve and AUC with 95% CI; plots on the right column show the disease prevalence in groups binned according to PRS (or PRS+covariates) percentile, where ORto50th% shows the odd ratio of each bin over the 25th bin (50 bins in total). PRS+covariates is the model with PRS, age, sex and PCs.

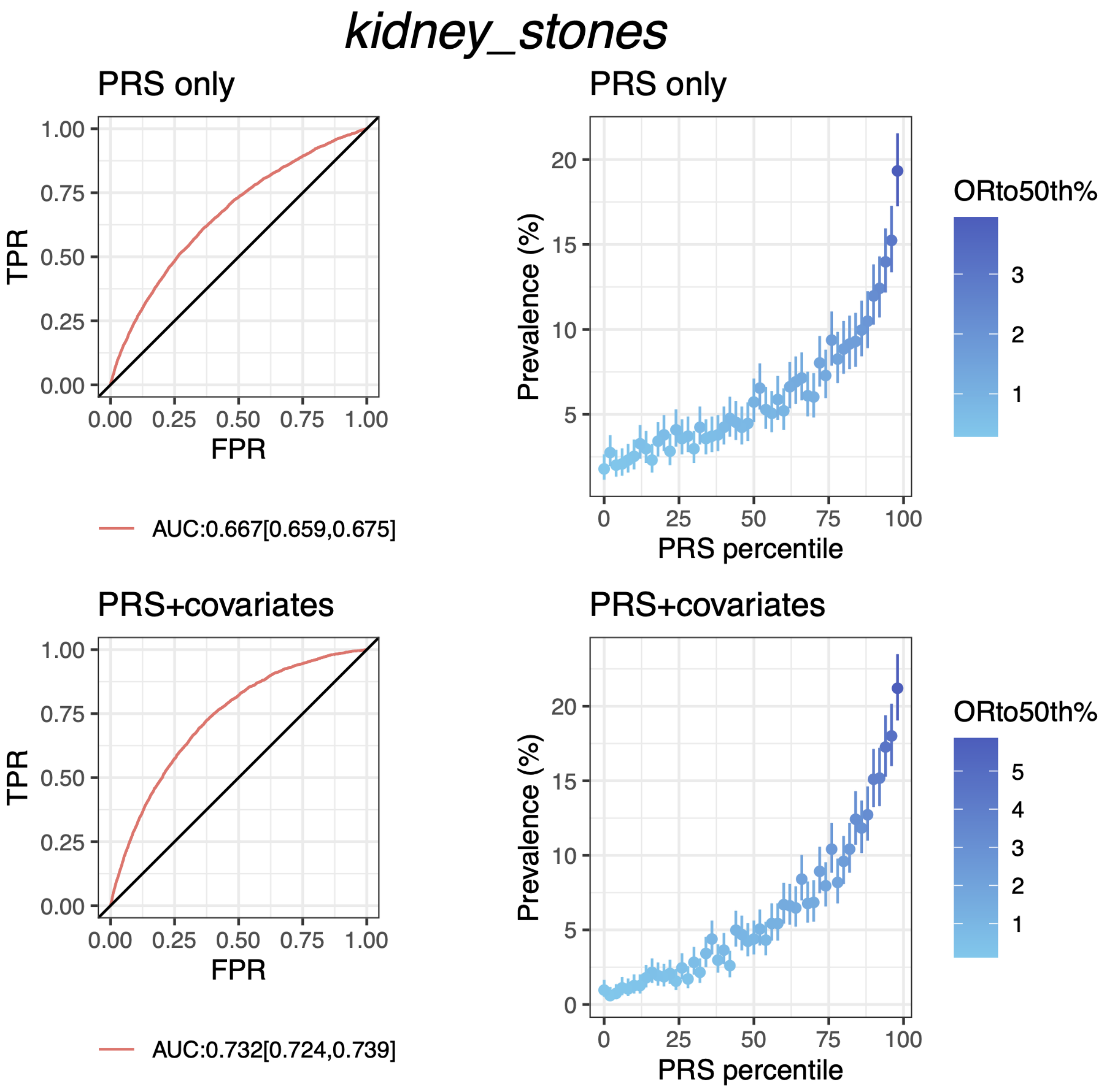


**Fig S7.J PRS performance for kidney stones in testing data from discovery dataset (Latine population)**

Plots on the left column show the receiver operating characteristics curve and AUC with 95% CI; plots on the right column show the disease prevalence in groups binned according to PRS (or PRS+covariates) percentile, where ORto50th% shows the odd ratio of each bin over the 25th bin (50 bins in total). PRS+covariates is the model with PRS, age, sex and PCs.

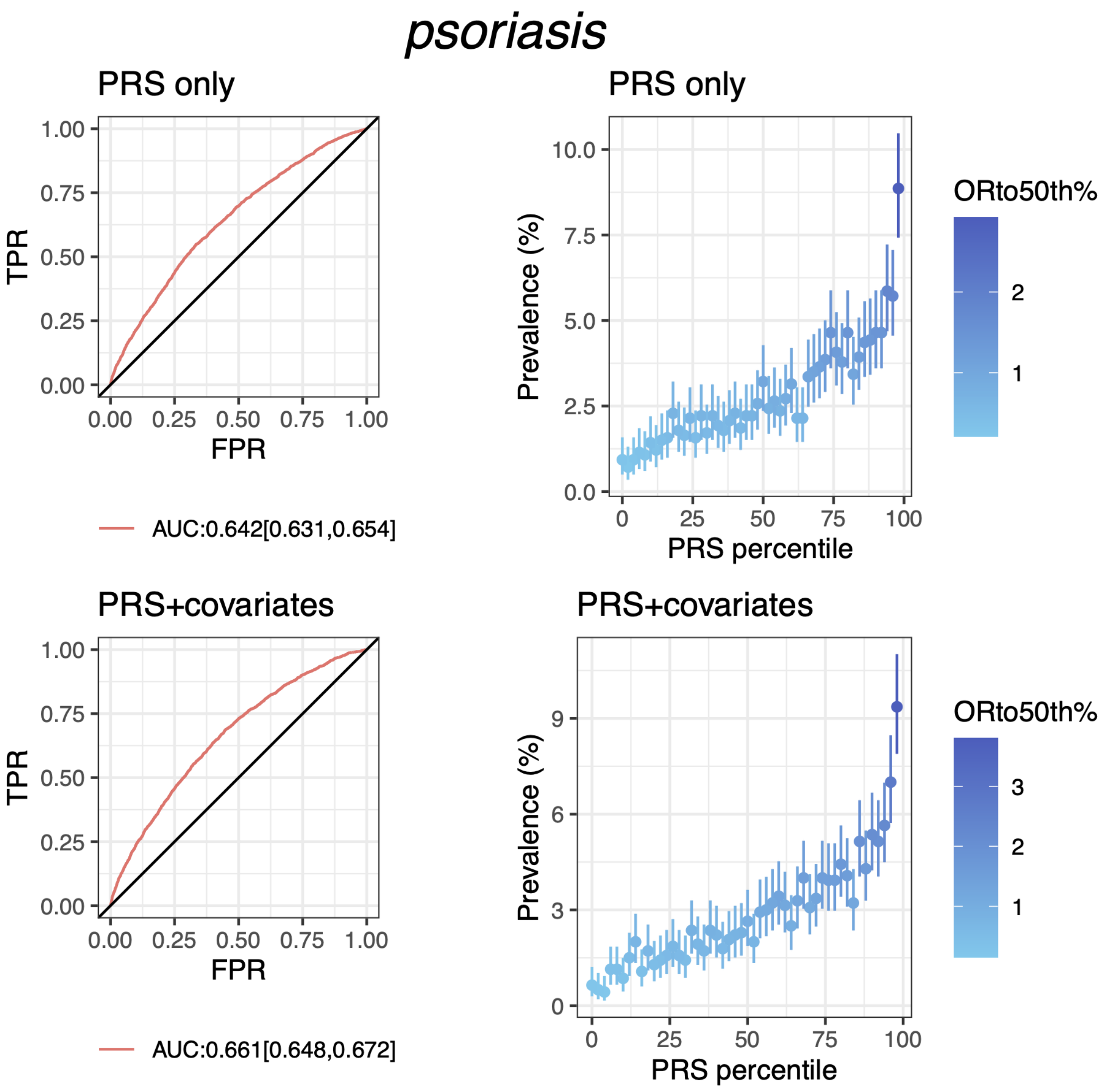


**Fig S7.K PRS performance for psoriasis in testing data from discovery dataset (Latine population)**

Plots on the left column show the receiver operating characteristics curve and AUC with 95% CI; plots on the right column show the disease prevalence in groups binned according to PRS (or PRS+covariates) percentile, where ORto50th% shows the odd ratio of each bin over the 25th bin (50 bins in total). PRS+covariates is the model with PRS, age, sex and PCs.

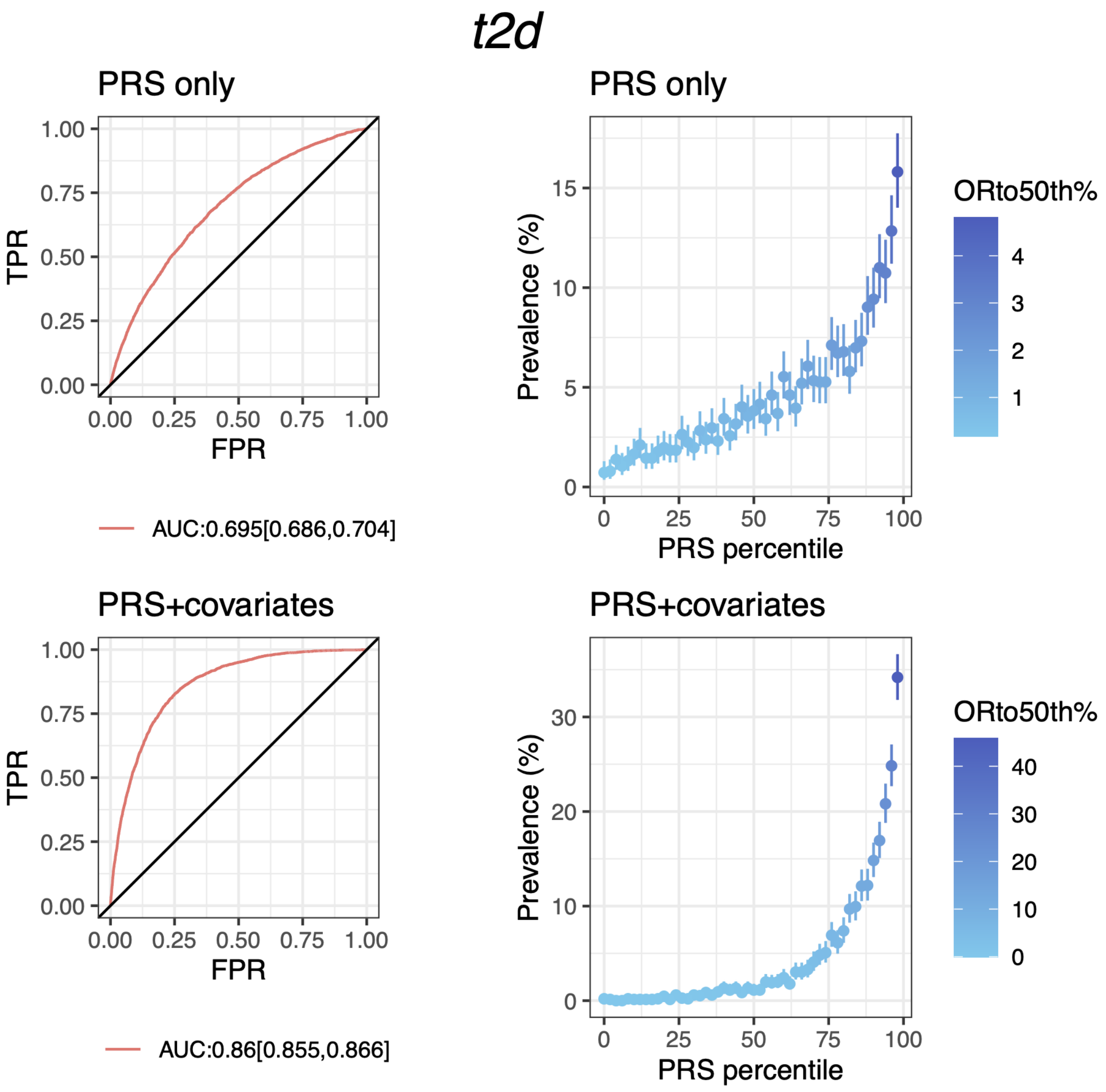


**Fig S7.L PRS performance for type 2 diabetes in testing data from discovery dataset (Latine population)**

Plots on the left column show the receiver operating characteristics curve and AUC with 95% CI; plots on the right column show the disease prevalence in groups binned according to PRS (or PRS+covariates) percentile, where ORto50th% shows the odd ratio of each bin over the 25th bin (50 bins in total). PRS+covariates is the model with PRS, age, sex and PCs.

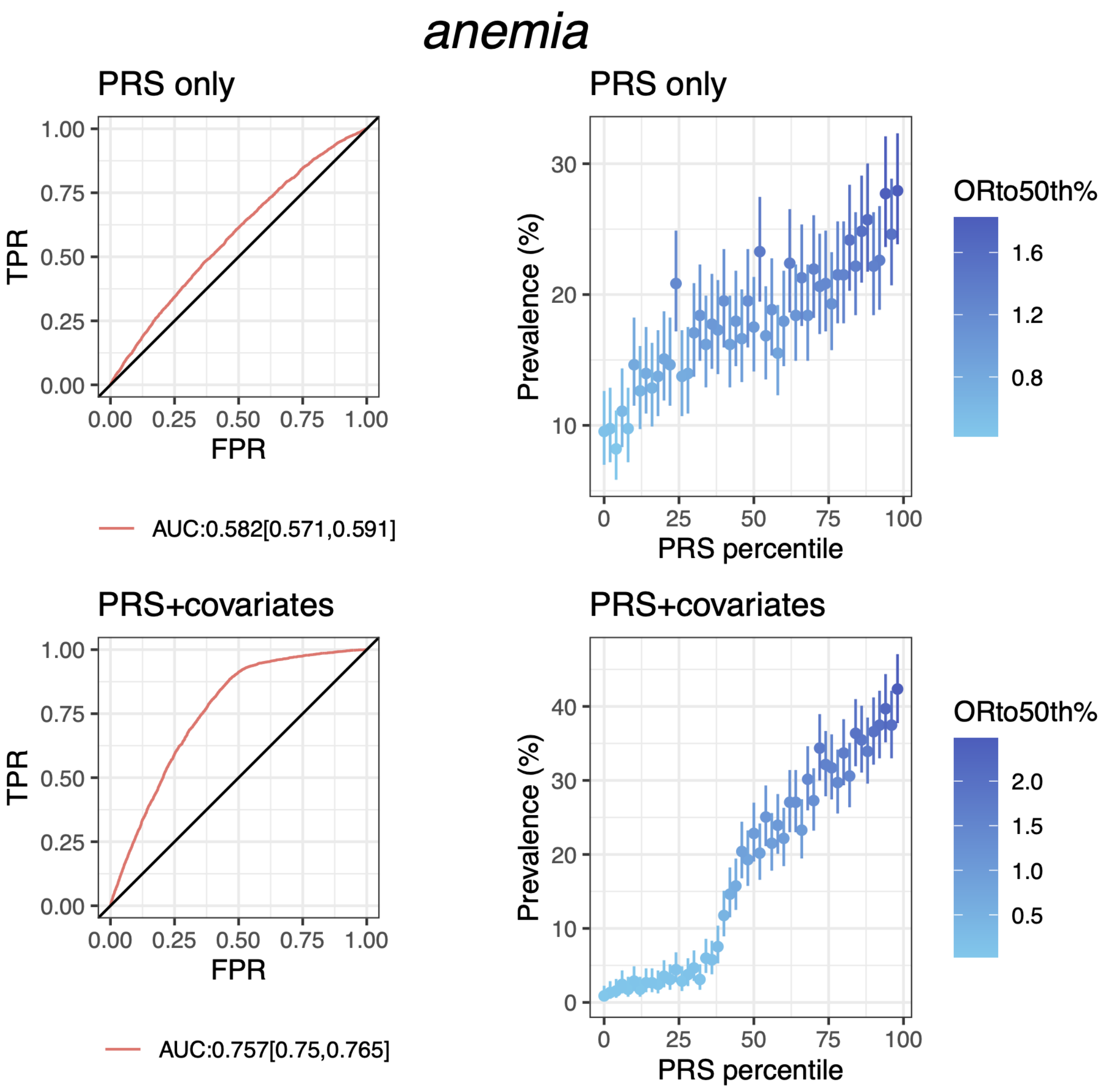


**Fig S8.A PRS performance for anemia in testing data from discovery dataset (African American population)**

Plots on the left column show the receiver operating characteristics curve and AUC with 95% CI; plots on the right column show the disease prevalence in groups binned according to PRS (or PRS+covariates) percentile, where ORto50th% shows the odd ratio of each bin over the 25th bin (50 bins in total). PRS+covariates is the model with PRS, age, sex and PCs.

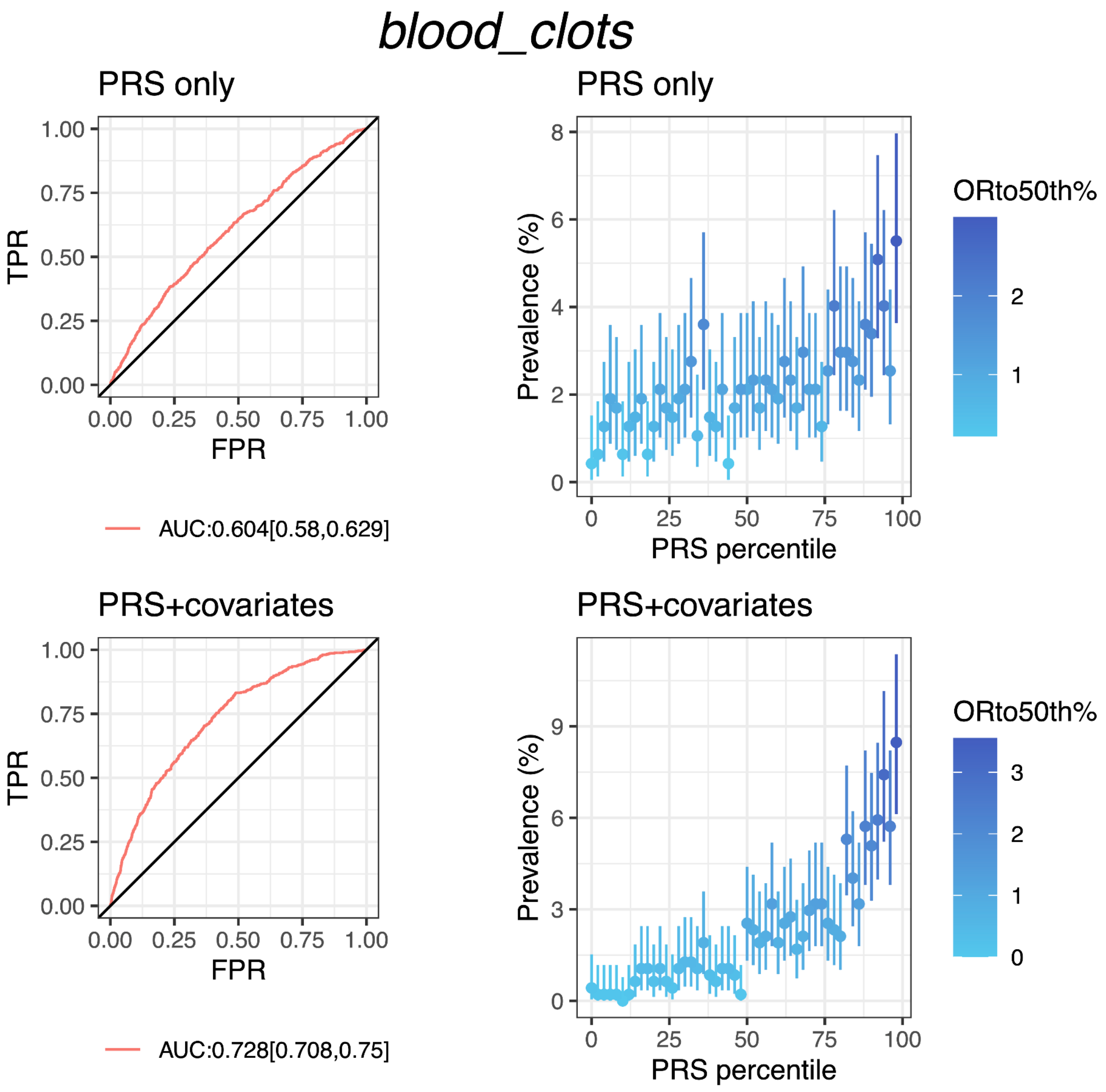


**Fig S8.B PRS performance for blood clots in testing data from Discovery dataset (African American population)**

Plots on the left column show the receiver operating characteristics curve and AUC with 95% CI; plots on the right column show the disease prevalence in groups binned according to PRS (or PRS+covariates) percentile, where ORto50th% shows the odd ratio of each bin over the 25th bin (50 bins in total). PRS+covariates is the model with PRS, age, sex and PCs.

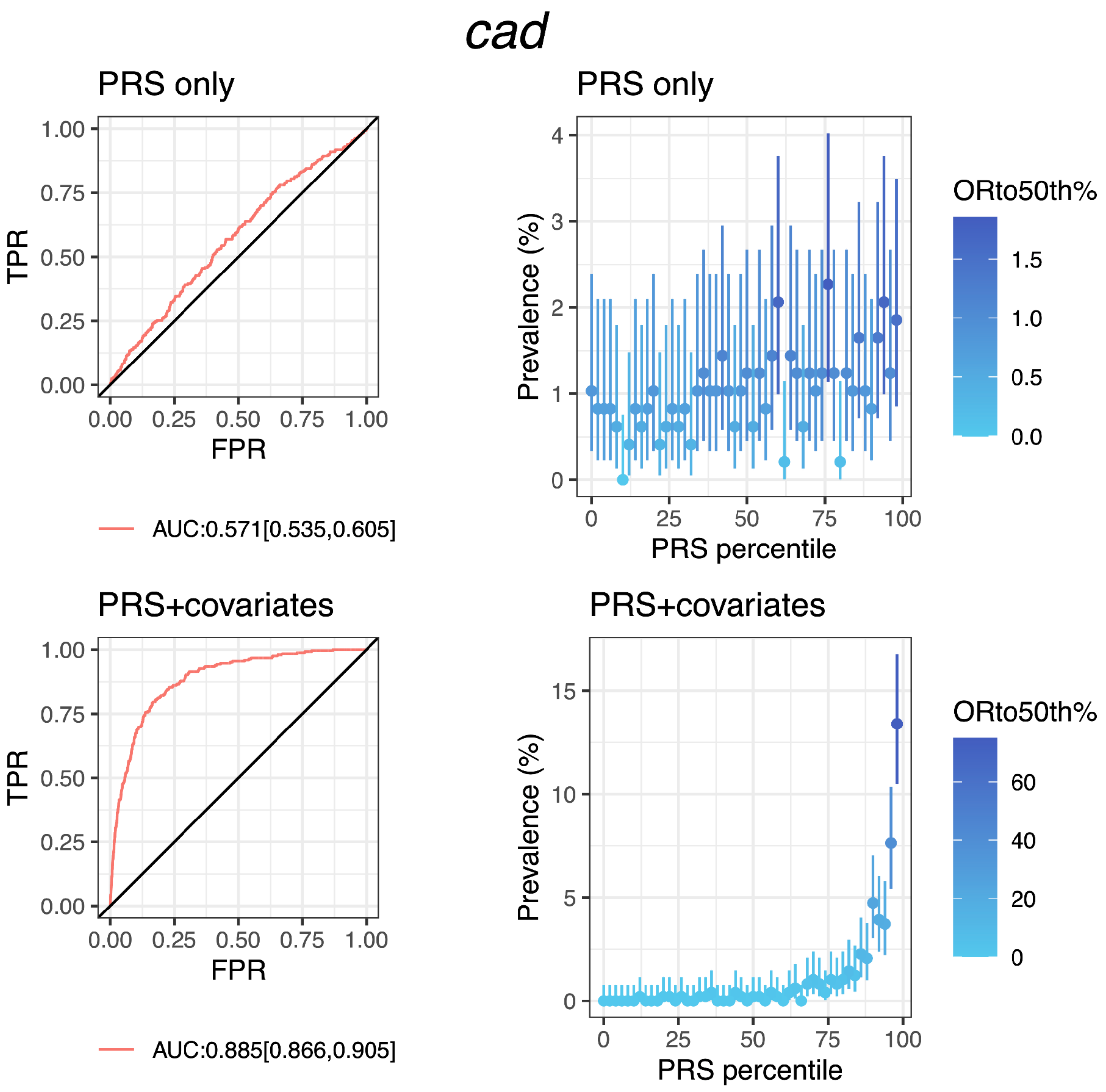


**Fig S8.C PRS performance for coronary heart disease in testing data from discovery dataset (African American population)**

Plots on the left column show the receiver operating characteristics curve and AUC with 95% CI; plots on the right column show the disease prevalence in groups binned according to PRS (or PRS+covariates) percentile, where ORto50th% shows the odd ratio of each bin over the 25th bin (50 bins in total). PRS+covariates is the model with PRS, age, sex and PCs.

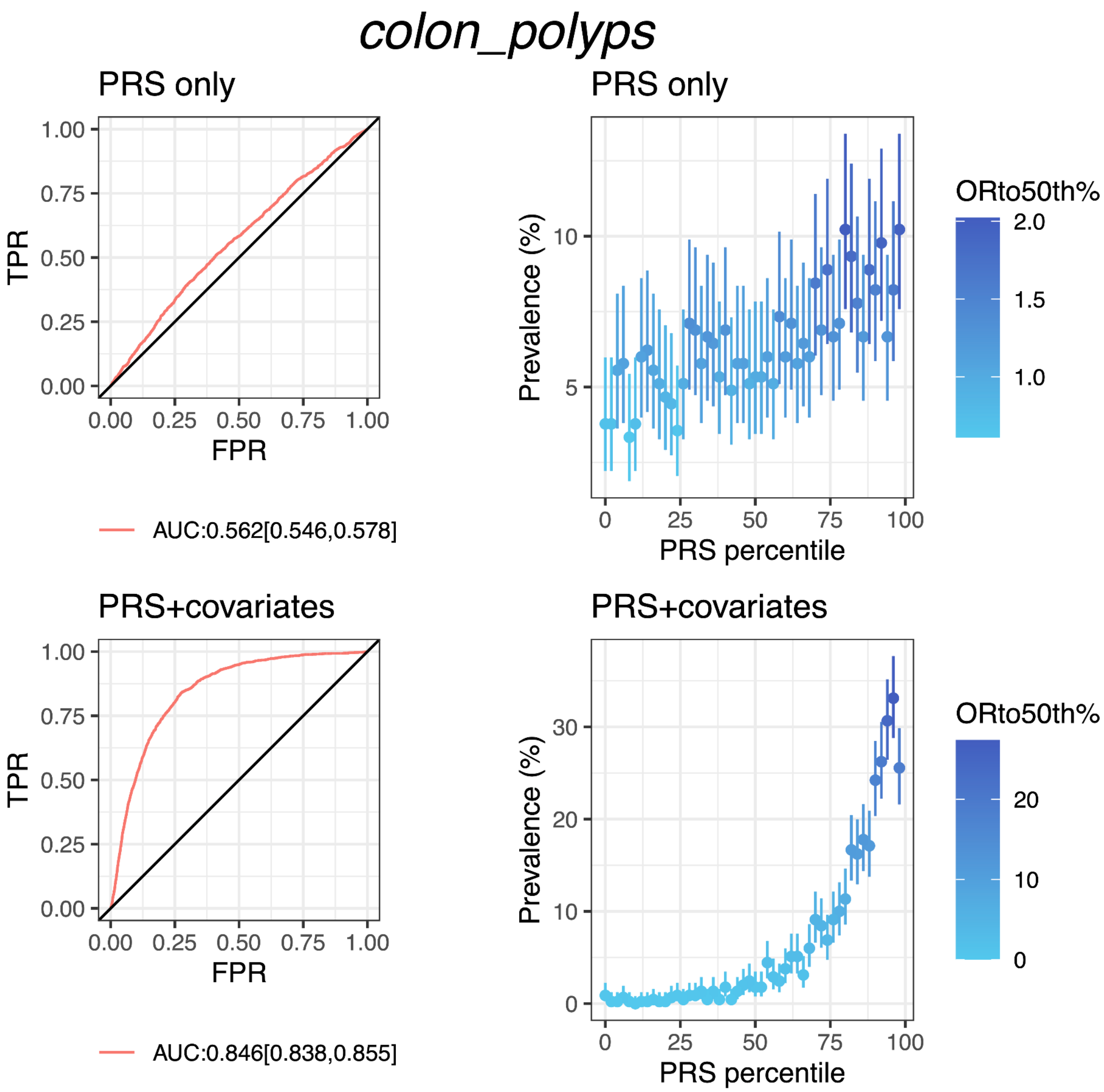


**Fig S8.D PRS performance for colon polyps in testing data from discovery dataset (African American population)**

Plots on the left column show the receiver operating characteristics curve and AUC with 95% CI; plots on the right column show the disease prevalence in groups binned according to PRS (or PRS+covariates) percentile, where ORto50th% shows the odd ratio of each bin over the 25th bin (50 bins in total). PRS+covariates is the model with PRS, age, sex and PCs.

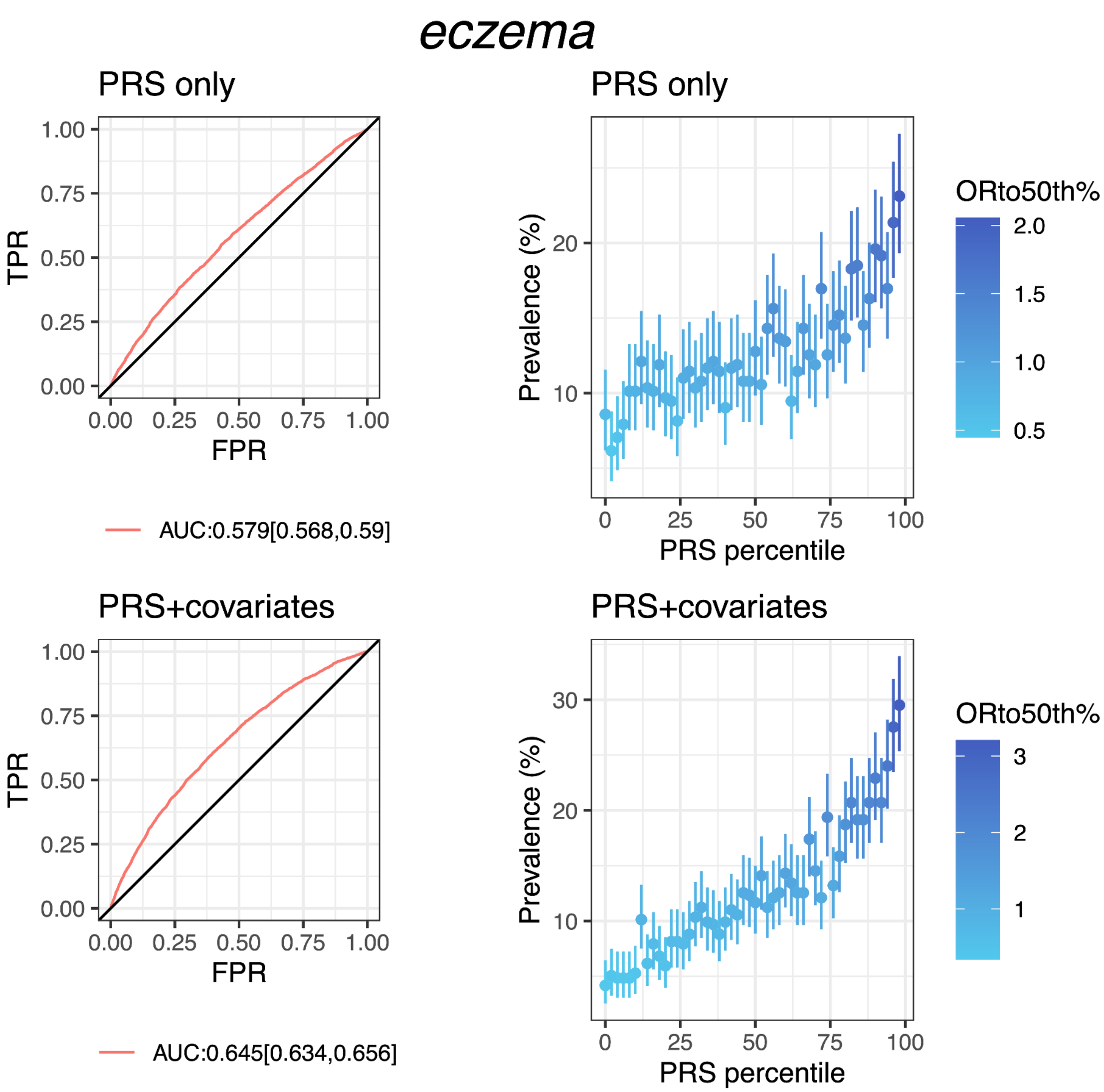


**Fig S8.E PRS performance for eczema in testing data from discovery dataset (African American population)**

Plots on the left column show the receiver operating characteristics curve and AUC with 95% CI; plots on the right column show the disease prevalence in groups binned according to PRS (or PRS+covariates) percentile, where ORto50th% shows the odd ratio of each bin over the 25th bin (50 bins in total). PRS+covariates is the model with PRS, age, sex and PCs.

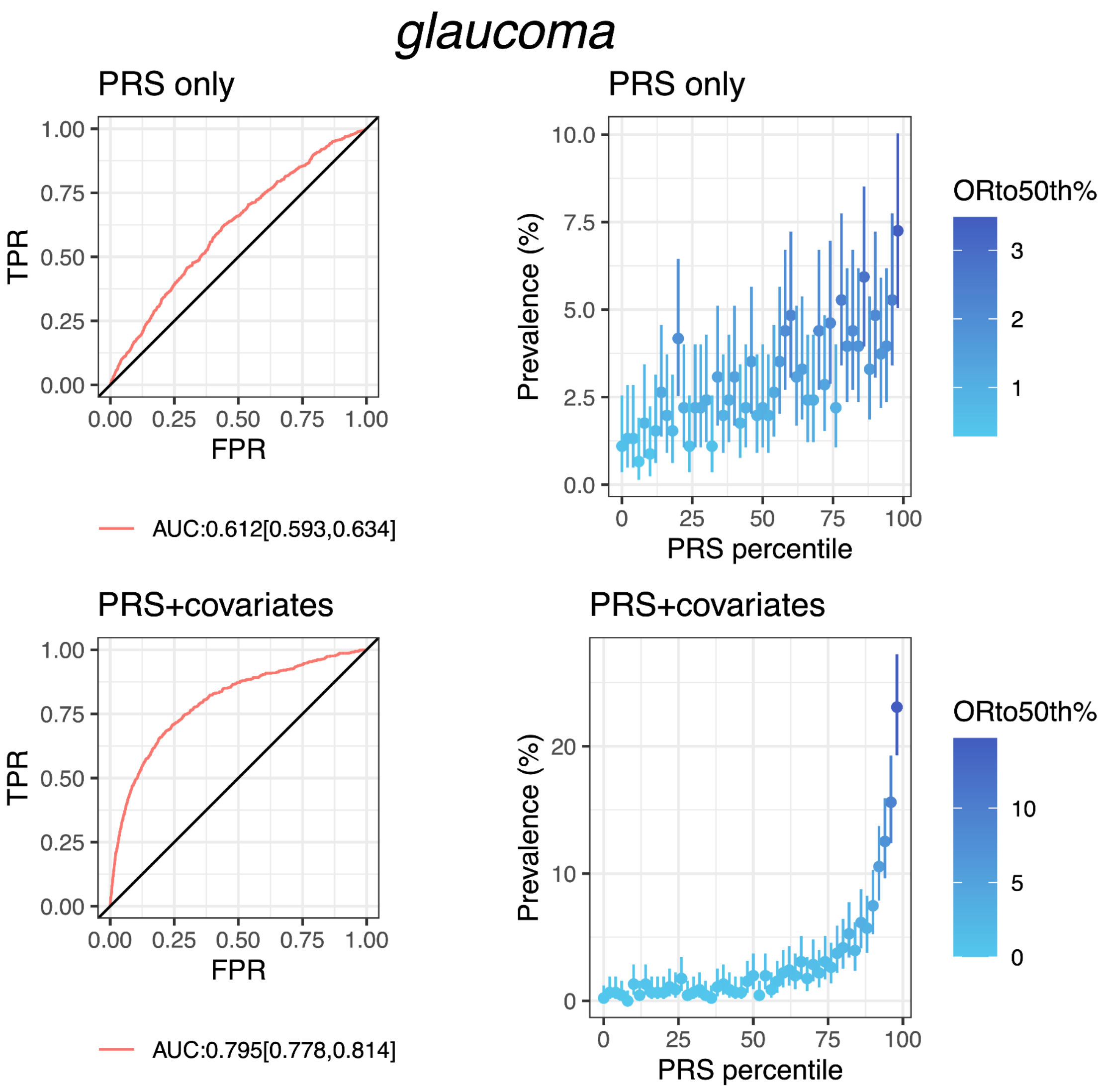


**Fig S8.F PRS performance for glaucoma in testing data from discovery dataset (African American population)**

Plots on the left column show the receiver operating characteristics curve and AUC with 95% CI; plots on the right column show the disease prevalence in groups binned according to PRS (or PRS+covariates) percentile, where ORto50th% shows the odd ratio of each bin over the 25th bin (50 bins in total). PRS+covariates is the model with PRS, age, sex and PCs.

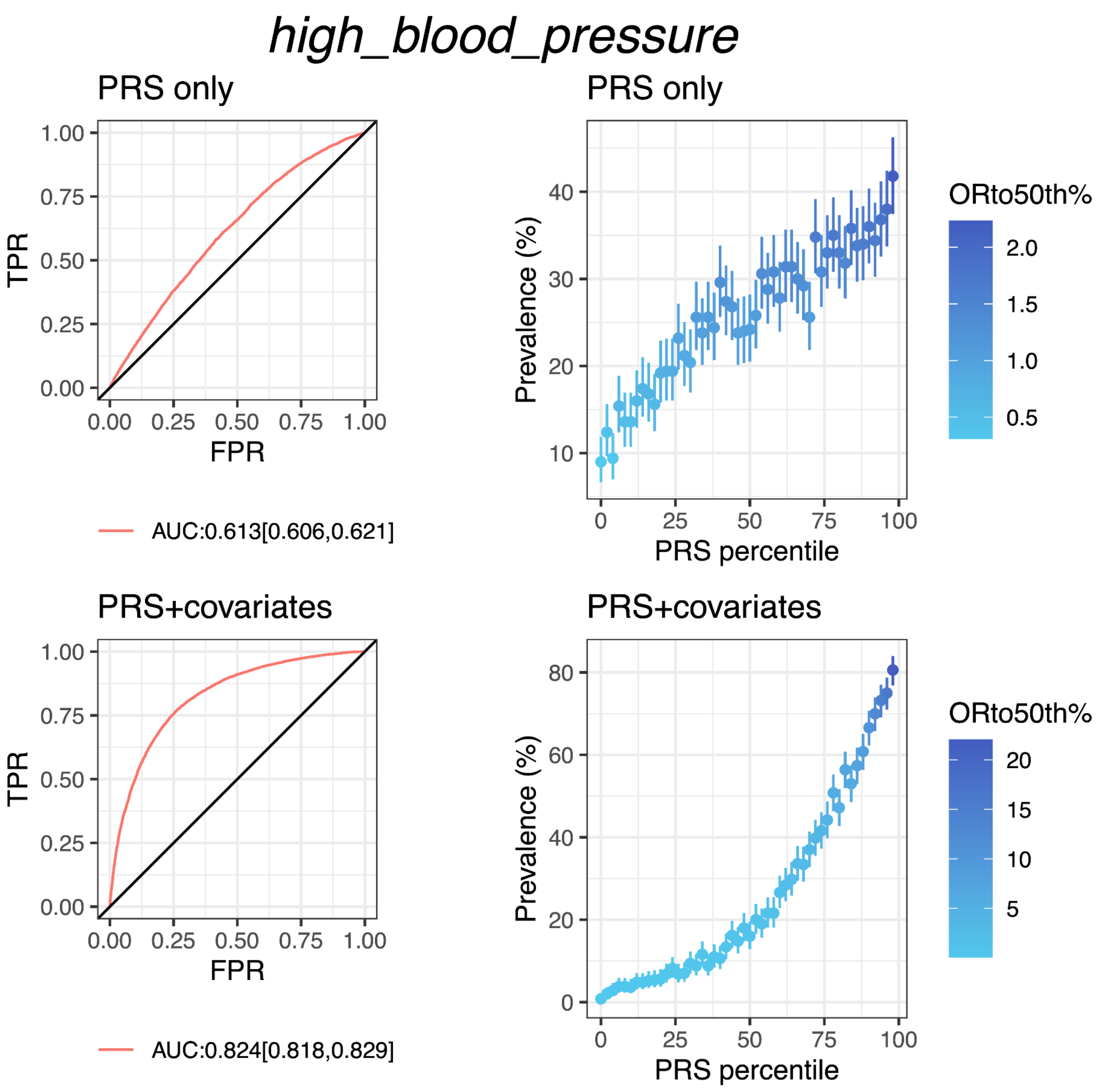


**Fig S8.G PRS performance for high blood pressure in testing data from Discovery dataset (African American population)**

Plots on the left column show the receiver operating characteristics curve and AUC with 95% CI; plots on the right column show the disease prevalence in groups binned according to PRS (or PRS+covariates) percentile, where ORto50th% shows the odd ratio of each bin over the 25th bin (50 bins in total). PRS+covariates is the model with PRS, age, sex and PCs.

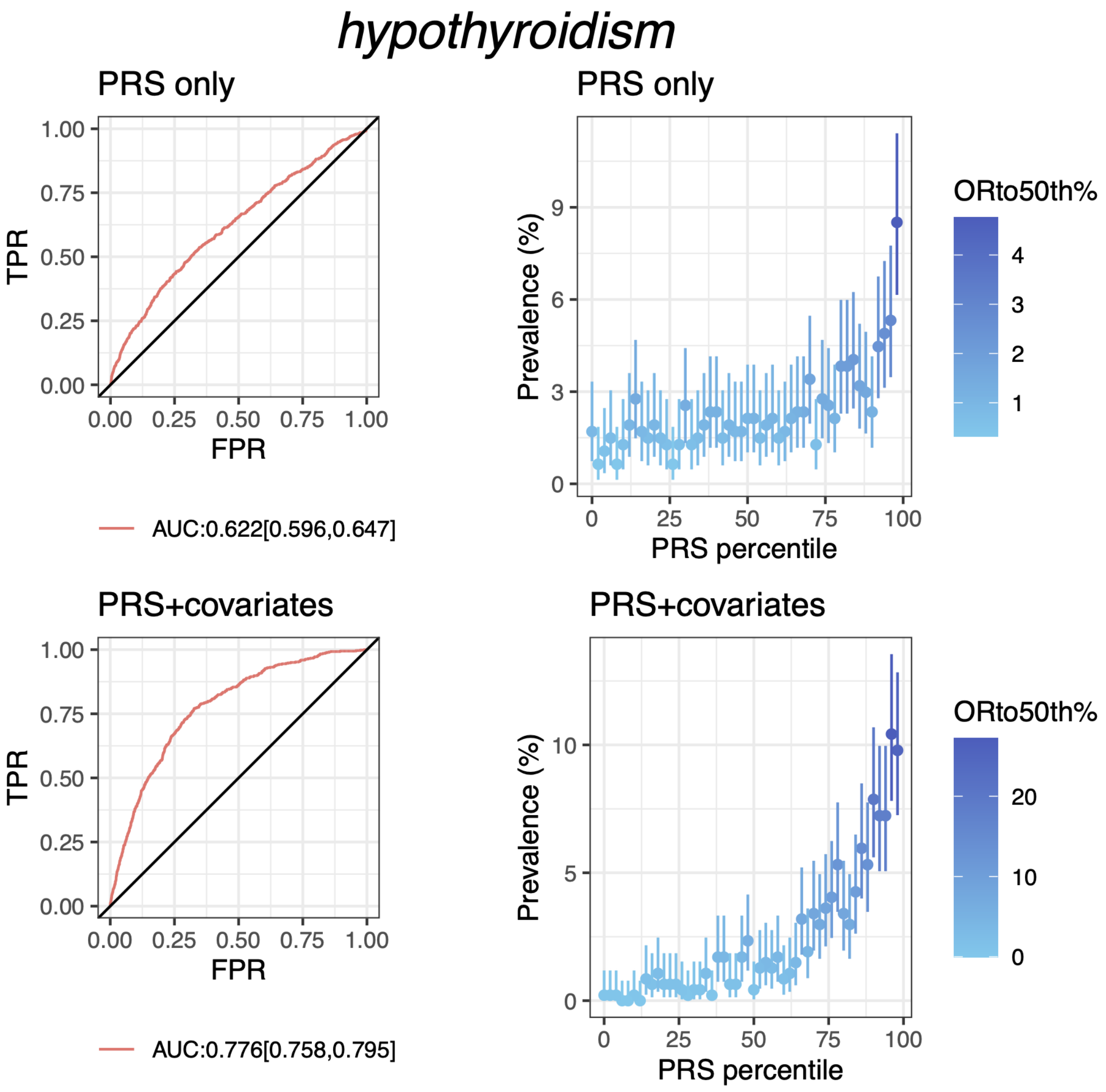


**Fig S8.H PRS performance for hypothyroidism in testing data from Discovery dataset (African American population)**

Plots on the left column show the receiver operating characteristics curve and AUC with 95% CI; plots on the right column show the disease prevalence in groups binned according to PRS (or PRS+covariates) percentile, where ORto50th% shows the odd ratio of each bin over the 25th bin (50 bins in total). PRS+covariates is the model with PRS, age, sex and PCs.

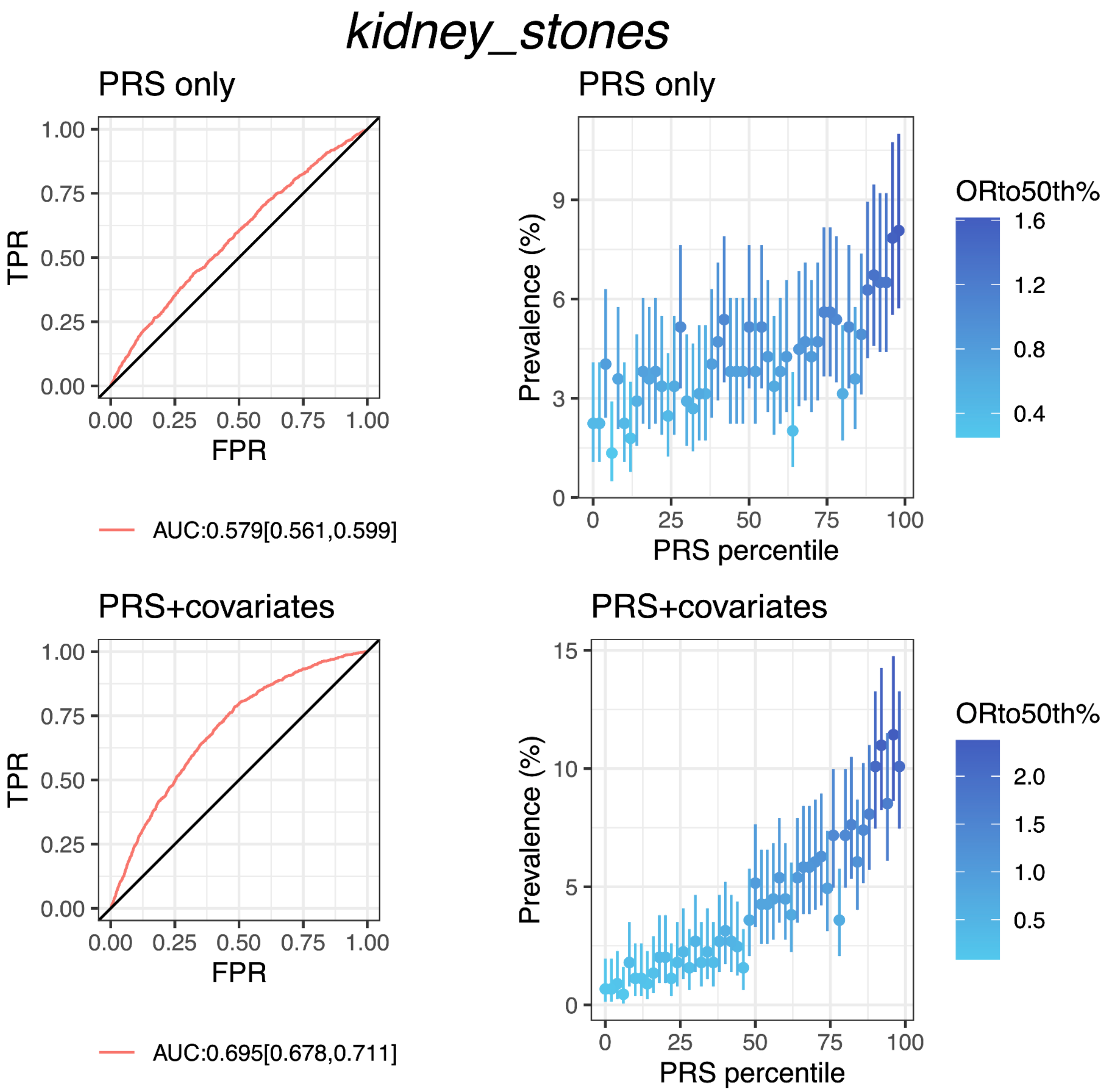


**Fig S8.I PRS performance for kidney stones in testing data from discovery dataset (African American population)**

Plots on the left column show the receiver operating characteristics curve and AUC with 95% CI; plots on the right column show the disease prevalence in groups binned according to PRS (or PRS+covariates) percentile, where ORto50th% shows the odd ratio of each bin over the 25th bin (50 bins in total). PRS+covariates is the model with PRS, age, sex and PCs.

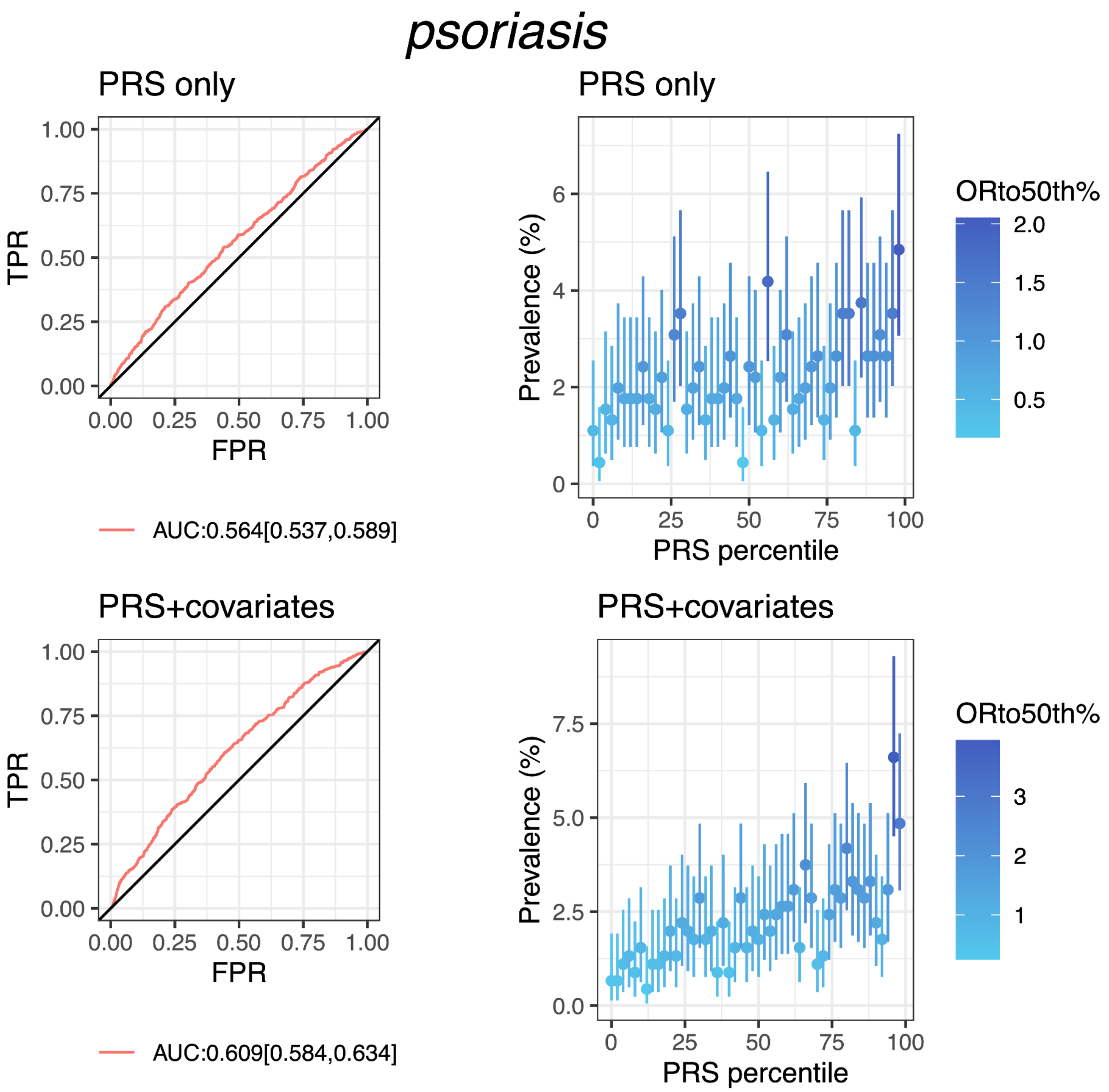


**Fig S8.J PRS performance for psoriasis in testing data from discovery dataset (African American population)**

Plots on the left column show the receiver operating characteristics curve and AUC with 95% CI; plots on the right column show the disease prevalence in groups binned according to PRS (or PRS+covariates) percentile, where ORto50th% shows the odd ratio of each bin over the 25th bin (50 bins in total). PRS+covariates is the model with PRS, age, sex and PCs.

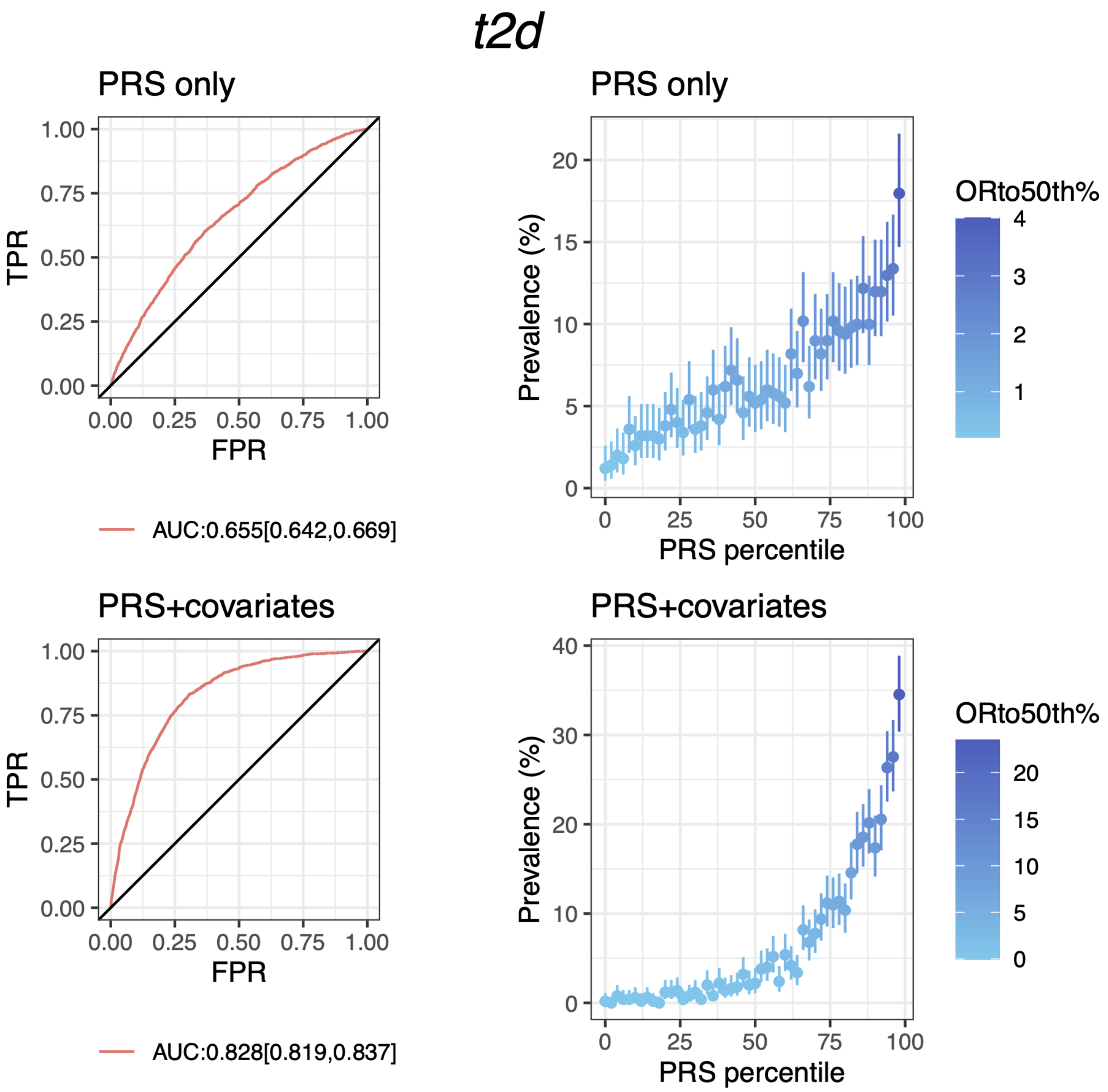


**Fig S8.K PRS performance for type 2 diabetes in testing data from discovery dataset (African American population)**

Plots on the left column show the receiver operating characteristics curve and AUC with 95% CI; plots on the right column show the disease prevalence in groups binned according to PRS (or PRS+covariates) percentile, where ORto50th% shows the odd ratio of each bin over the 25th bin (50 bins in total). PRS+covariates is the model with PRS, age, sex and PCs.

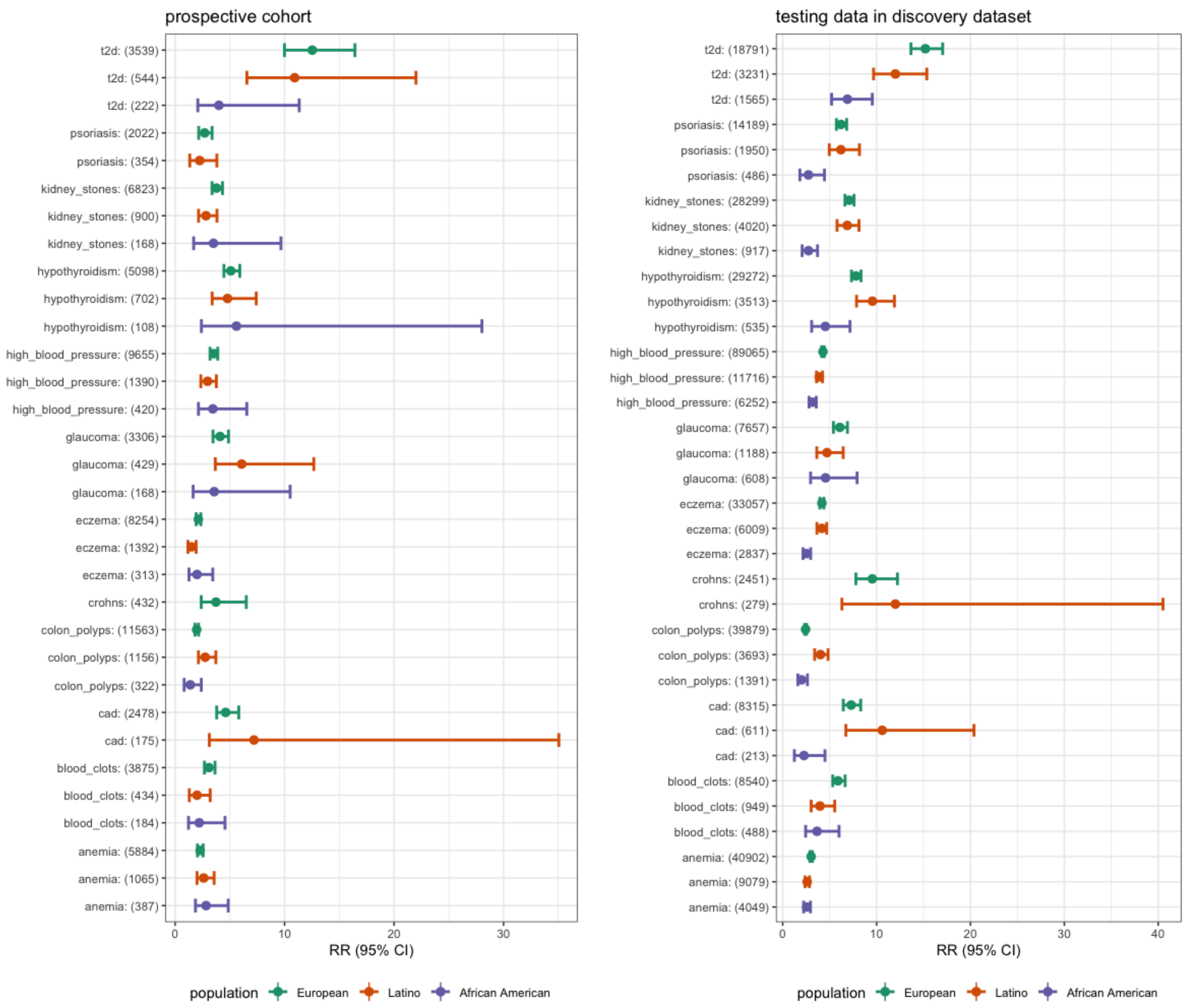


**Fig S9. PRS performance for diseases across multiple ancestries.**
Estimates represent the relative risk of disease comparing top 10% to bottom 10% of each PRS. The prospective cohort on the left is the dataset in which individuals reported new disease within the past year. The testing data represents prevalent cases and controls in the hold-out testing data for prediction performance assessment in the discovery dataset.

### Supplementary Tables

**Table S1. Clinical endpoint definitions in the discovery dataset**

| Phenotype name | Survey name | Questions |
|---|---|---|
| CHD | 1, Health_intake / Health profile.<br>2, Health Update<br>3, COVID-19 (covid_19_baseline) / COVID-19 Monthly Follow-Up<br>4, Pulmonary Fibrosis<br>5,Idiopathic Pulmonary Fibrosis<br>6, Research Snippet(hh_coronary_artery_dis) | 1 "Have you ever been diagnosed by a doctor with any of the following cardiovascular conditions?"/"Coronary artery disease/heart disease".<br>2, same as 1; and "In the past 12 months, have you been newly diagnosed with any of the following conditions by a medical professional?<br>Irregular heartbeat (arrhythmia)<br>Coronary artery disease (CAD) or coronary heart disease (CHD)<br>Anemia<br>A bleeding disorder or bleeding tendency (such as hemophilia)" and "Were you diagnosed with coronary artery disease (CAD) or coronary heart disease (CHD)?"<br>3, same as 1.<br>4, same as 1.<br>5, same as 1.<br>6, "Have you ever been diagnosed with coronary artery disease?". |
| T2D | 1, Health_intake / Health profile<br>2, Research Snippet (hh_type_2_diabetes)<br>3, COVID-19 (covid_19_baseline) / COVID-19 Monthly Follow-Up<br>4, Health Update<br>5, Hypertrophic Cardiomyopathy Survey | 1, "Have you ever been diagnosed with or treated for any of the following conditions?"/"Type 2 Diabetes", and "What types of diabetes were you diagnosed with? Please select all that apply." : "Type 2 diabetes"<br>2, "Have you ever been diagnosed with type 2 diabetes?".<br>3, "What types of diabetes were you diagnosed with? Please select all that apply." : "Type 2 diabetes". |

| | | |
|---|---|---|
| | | 4, "In the past 12 months, have you been newly diagnosed with any of the following conditions by a medical professional?<br>Lupus<br>Diabetes or high blood sugar<br>Any type of thyroid disease, not thyroid cancer<br>High blood pressure" and "What types of diabetes were you diagnosed with? Please select all that apply." : "Type 2 diabetes"<br>5, "Have you ever been diagnosed with, or treated for, diabetes, gestational diabetes (diabetes during pregnancy), or high blood sugar?". |
| High blood pressure | 1, Health_intake / Health profile<br>2, COVID-19 (covid_19_baseline) / COVID-19 Monthly Follow-Up<br>3, Hypertrophic Cardiomyopathy Survey (hcm)<br>4, Research Snippet (hh_high_blood_pressure)<br>5, Lupus - Year End (sle_final)<br>6, Health Update | 1, "Have you ever been diagnosed by a doctor with any of the following cardiovascular conditions?"/"High blood pressure".<br>2, "Have you ever been diagnosed with, or treated for, high blood pressure?"<br>3, same as 2.<br>4, "Have you ever been diagnosed with high blood pressure?"<br>5, same as 4.<br>6, "In the past 12 months, have you been newly diagnosed with any of the following conditions by a medical professional?<br>Lupus<br>Diabetes or high blood sugar<br>Any type of thyroid disease, not thyroid cancer<br>High blood pressure" and "Were you diagnosed with high blood pressure?" |
| Anemia | 1, Health_intake / Health profile<br>2, Bleeding and Blood Clots | 1, "Have you ever been diagnosed by a doctor with any of the following blood conditions?"/"Anemia" and "Did your doctor ever diagnose you with anemia |

| | | |
|---|---|---|
| | (bleeding_blood_clots)<br>3, Research Snippet (anemia_gh)<br>4, Parkinson's Prevention Survey<br>5, Health Update | related to your chronic kidney disease?".<br>2, "Have you ever been diagnosed with or treated for anemia?".<br>3, "Have you ever been diagnosed with anemia?".<br>4, same as 3.<br>5, "In the past 12 months, have you been newly diagnosed with any of the following conditions by a medical professional?<br>Irregular heartbeat (arrhythmia)<br>Coronary artery disease (CAD) or coronary heart disease (CHD)<br>Anemia<br>A bleeding disorder or bleeding tendency (such as hemophilia)" and "Were you diagnosed with anemia?". |
| Blood clots | 1, Health_intake / Health profile<br>2, Research Snippet (hh_deep_vein_thrombosis)<br>3, Lupus - Year End<br>4, Bleeding and Blood Clots (bleeding_blood_clots)<br>5, Health Update | 1, "Have you ever been diagnosed by a doctor with any of the following blood conditions?"/"Blood clots"; "Have you ever been diagnosed with or treated for any of the following conditions?"/"A blood clot in your arms or legs (deep vein thrombosis or DVT)";<br>2, "Have you ever been diagnosed with deep vein thrombosis?".<br>3, "Have you ever been diagnosed with or treated for any of the following conditions?".<br>4, "Have you ever experienced any of the following blood clotting events? Please select all that apply." : "Deep vein thrombosis or venous thrombosis": "Deep vein thrombosis or venous thrombosis", "Pulmonary embolism", "Arterial thrombosis", "Superficial thrombophlebitis".<br>5, "In the past 12 months, have you experienced any of the following health problems or events? Your best guess is fine."/"A blood clot". |

| | | |
|---|---|---|
| Hypothyroidism | 1, Health_intake / Health profile<br>2, Research Snippet (hypothyroidism, hypothyroid_medication_now, tsh_elevated)<br>3, Health Update | 1, "Have you ever been diagnosed by a doctor with any of the following thyroid conditions?"/"Hypothyroidism".<br>2, "Have you ever been diagnosed with hypothyroidism (underactive thyroid)? ", "Do you currently take medication for hypothyroidism (low thyroid hormone levels)?", "Have you ever been told by a doctor that your thyroid stimulating hormone (TSH) levels were elevated, indicating hypothyroidism? "<br>3, "In the past 12 months, have you been newly diagnosed with any of the following conditions by a medical professional?<br>Lupus<br>Diabetes or high blood sugar<br>Any type of thyroid disease, not thyroid cancer<br>High blood pressure" and "What type of thyroid condition were you diagnosed with?" : "Underactive thyroid or hypothyroidism" |
| Kidney stones | 1, Health_intake / Health profile<br>2, Research Snippet (hp_kidney_stones_v2)<br>3, Health Update | 1, "Have you ever been diagnosed with, or treated for, kidney stones?".<br>2, "Have you ever been diagnosed with, or treated for, kidney stones?".<br>3, "In the past 12 months, have you experienced any of the following health problems or events? Your best guess is fine."/"Kidney stones" |
| Crohn's disease | 1, Health_intake / Health profile<br>2, Inflammatory Bowel Disease Background<br>3, Research Snippet (crohns)<br>4, Lupus - Year End<br>5, Health Update | 1, "Have you ever been diagnosed by a doctor with any of the following intestinal conditions?"/"Crohn's disease".<br>2, "What kind of IBD do you have?" : "Crohn's disease"<br>3, "Have you ever been diagnosed with Crohn's disease?"<br>4, same as 1.<br>5, "In the past 12 months, have you been |

| | | |
|---|---|---|
| | | newly diagnosed with any of the following conditions by a medical professional?<br>Acid reflux or gastroesophageal reflux disease (GERD)<br>Irritable bowel syndrome (IBS)<br>Colon polyps<br>Crohn's disease<br>Ulcerative colitis" and "Were you diagnosed with Crohn's disease?" |
| Colon polyps | 1, Health_intake / Health profile<br>2, Research Snippet (colon_polyps_gh)<br>3, Health Update | 1, "Have you ever been diagnosed by a doctor with any of the following intestinal conditions?"/"Colon polyps".<br>2, "Have you ever been diagnosed with colon polyps?".<br>3, "In the past 12 months, have you been newly diagnosed with any of the following conditions by a medical professional?<br>Acid reflux or gastroesophageal reflux disease (GERD)<br>Irritable bowel syndrome (IBS)<br>Colon polyps<br>Crohn's disease<br>Ulcerative colitis" and "Were you diagnosed with colon polyps?". |
| Glaucoma | 1, Health_intake / Health profile<br>2, Research Snippet (exfoliation_glaucoma)<br>3, Health Update | 1, "Have you ever been diagnosed by a doctor with any of the following vision conditions?"/"Glaucoma".<br>2, "Have you ever been diagnosed with exfoliation glaucoma?"<br>3, "In the past 12 months, have you been newly diagnosed with any of the following conditions by a medical professional?<br>Macular degeneration<br>Dry eyes<br>Cataracts<br>Glaucoma" and "Were you diagnosed with glaucoma or possible glaucoma?" |
| Psoriasis | 1, Health_intake / | 1, "Have you ever been diagnosed by a |

| | | |
|---|---|---|
| | Health profile<br>2, Research Snippet (psoriasis, hh_psoriasis)<br>3, Inflammatory Bowel Disease Background<br>4, Eczema<br>5, Psoriasis<br>6, Lupus - Year End<br>7, Health Update | doctor with any of the following autoimmune conditions?"/"Psoriasis".<br>2, "Have you ever been diagnosed with psoriasis?".<br>3, "Which skin conditions have you experienced?"/"Psoriasis (dry red patches of skin covered with scales)".<br>4, "Have you ever been diagnosed with, or treated for, psoriasis?".<br>5, same as 4.<br>6, "Have you ever been diagnosed with or treated for any of the following conditions?"/"Psoriasis"<br>7, "In the past 12 months, have you been newly diagnosed with any of the following conditions by a medical professional?<br>Eczema (atopic dermatitis)<br>Psoriasis<br>Rosacea" and "Were you diagnosed with psoriasis?". |
| Eczema | 1, Health_intake / Health profile<br>2, Allergies<br>3, Inflammatory Bowel Disease Background<br>4, Research Snippet (hh_eczema)<br>5, Lupus - Year End<br>6, Health Update | 1, "Have you ever been diagnosed by a doctor with any of the following autoimmune conditions?"/"Eczema".<br>2, "Did you have any of these problems before you were 18 years old?"/"Eczema (atopic dermatitis)".<br>3, "Which skin conditions have you experienced?"/"Eczema".<br>4, "Have you ever been diagnosed with eczema?".<br>5, "Have you ever been diagnosed with or treated for any of the following conditions?"/"Eczema".<br>6, "In the past 12 months, have you been newly diagnosed with any of the following conditions by a medical professional?<br>Eczema (atopic dermatitis)<br>Psoriasis<br>Rosacea" and "Were you diagnosed with eczema (atopic dermatitis)?". |

**Table S2. Clinical endpoint definition from baseline and during follow-up**

| Phenotype name | Questions in health profile survey | Questions in health update survey |
| --- | --- | --- |
| CHD | "Have you ever been diagnosed by a doctor with any of the following cardiovascular conditions?"/"Coronary artery disease/heart disease". | "In the past 12 months, have you been newly diagnosed with any of the following conditions by a medical professional?<br>Irregular heartbeat (arrhythmia)<br>Coronary artery disease (CAD) or coronary heart disease (CHD)<br>Anemia<br>A bleeding disorder or bleeding tendency (such as hemophilia)" and "Were you diagnosed with coronary artery disease (CAD) or coronary heart disease (CHD)?" |
| T2D | "Have you ever been diagnosed with or treated for any of the following conditions?"/"Type 2 Diabetes" | "In the past 12 months, have you been newly diagnosed with any of the following conditions by a medical professional?<br>Lupus<br>Diabetes or high blood sugar<br>Any type of thyroid disease, not thyroid cancer<br>High blood pressure" and "What types of diabetes were you diagnosed with? Please select all that apply." : "Type 2 diabetes" |
| High blood pressure | "Have you ever been diagnosed by a doctor with any of the following cardiovascular conditions?"/"High blood pressure". | "In the past 12 months, have you been newly diagnosed with any of the following conditions by a medical professional?<br>Lupus<br>Diabetes or high blood sugar<br>Any type of thyroid disease, not thyroid cancer |

| | | |
|---|---|---|
| | | High blood pressure" and "Were you diagnosed with high blood pressure?" |
| Anemia | "Have you ever been diagnosed by a doctor with any of the following blood conditions?"/"Anemia" | "In the past 12 months, have you been newly diagnosed with any of the following conditions by a medical professional?<br>Irregular heartbeat (arrhythmia)<br>Coronary artery disease (CAD) or coronary heart disease (CHD)<br>Anemia<br>A bleeding disorder or bleeding tendency (such as hemophilia)" and "Were you diagnosed with anemia?". |
| Blood clots | "Have you ever been diagnosed by a doctor with any of the following blood conditions?"/"Blood clots" | "In the past 12 months, have you experienced any of the following health problems or events? Your best guess is fine."/"A blood clot". |
| Hypothyroidism | "Have you ever been diagnosed by a doctor with any of the following thyroid conditions?"/"Hypothyroidism". | "In the past 12 months, have you been newly diagnosed with any of the following conditions by a medical professional?<br>Lupus<br>Diabetes or high blood sugar<br>Any type of thyroid disease, not thyroid cancer<br>High blood pressure" and "What type of thyroid condition were you diagnosed with?" : "Underactive thyroid or hypothyroidism" |
| Kidney stones | "Have you ever been diagnosed with, or treated for, kidney stones?". | "In the past 12 months, have you experienced any of the following health problems or events? Your best guess is fine."/"Kidney stones" |
| Crohn's disease | "Have you ever been diagnosed by a doctor with any of the following intestinal | "In the past 12 months, have you been newly diagnosed with any of the following conditions by a medical professional? |

| | | |
|---|---|---|
| | conditions?"/"Crohn's disease". | Acid reflux or gastroesophageal reflux disease (GERD)<br>Irritable bowel syndrome (IBS)<br>Colon polyps<br>Crohn's disease<br>Ulcerative colitis" and "Were you diagnosed with Crohn's disease?" |
| Colon polyps | "Have you ever been diagnosed by a doctor with any of the following intestinal conditions?"/"Colon polyps". | "In the past 12 months, have you been newly diagnosed with any of the following conditions by a medical professional?<br>Acid reflux or gastroesophageal reflux disease (GERD)<br>Irritable bowel syndrome (IBS)<br>Colon polyps<br>Crohn's disease<br>Ulcerative colitis" and "Were you diagnosed with colon polyps?". |
| Glaucoma | "Have you ever been diagnosed by a doctor with any of the following vision conditions?"/"Glaucoma". | "In the past 12 months, have you been newly diagnosed with any of the following conditions by a medical professional?<br>Macular degeneration<br>Dry eyes<br>Cataracts<br>Glaucoma" and "Were you diagnosed with glaucoma or possible glaucoma?" |
| Psoriasis | "Have you ever been diagnosed by a doctor with any of the following autoimmune conditions?"/"Psoriasis". | "In the past 12 months, have you been newly diagnosed with any of the following conditions by a medical professional?<br>Eczema (atopic dermatitis)<br>Psoriasis<br>Rosacea" and "Were you diagnosed with psoriasis?". |
| Eczema | "Have you ever been diagnosed by a doctor with | "In the past 12 months, have you been newly diagnosed with any of the |

| | | |
|---|---|---|
| | any of the following autoimmune conditions?"/"Eczema". | following conditions by a medical professional?<br>Eczema (atopic dermatitis)<br>Psoriasis<br>Rosacea" and "Were you diagnosed with eczema (atopic dermatitis)?". |

**Table S3. Performance of PRS for incident endpoints across different ancestries**

| Endpoint | Ancestry | 1st year incident cases/N | AUC PRS (PRS + age + sex + 5PCs) discovery | Cumulative incidence (median age at diagnosis) | | | Top vs bottom decile PRS |
|---|---|---|---|---|---|---|---|
| | | | | All | Bottom 10% PRS | Top 10% PRS | Relative risk (95%CI) |
| Coronary heart disease | European | 2478/ 763557 | 0.66 (0.90) | 0.32% (66) | 0.14% (70) | 0.63% (64) | 4.61 [3.79, 5.81] |
| | Latine | 175/ 101865 | 0.67 (0.92) | 0.17% | 0.049% | 0.35% | 7.2 [3.12, 35.05] |
| Type 2 diabetes | European | 3539/ 739992 | 0.71 (0.84) | 0.48% (60) | 0.095% (64.5) | 1.19% (55) | 12.53 [9.99, 16.43] |
| | African American | 222/ 23849 | 0.66 (0.83) | 0.93% | 0.34% | 1.34% | 4 [2.07, 11.33] |
| | Latine | 544/ 97343 | 0.70 (0.86) | 0.56% | 0.12% | 1.35% | 10.92 [6.55, 22] |
| High blood pressure | European | 9655/ 552962 | 0.64 (0.82) | 1.75% (55) | 0.88% (63) | 3.09% (47) | 3.52 [3.19, 3.89] |

| | | | | | | | |
|---|---|---|---|---|---|---|---|
| | African American | 420/ 17517 | 0.61 (0.82) | 2.40% | 1.03% | 3.54% | 3.44 [2.13, 6.55] |
| | Latine | 1390/ 81862 | 0.63 (0.82) | 1.70% | 1.04% | 3.08% | 2.96 [2.35, 3.77] |
| Anemia | European | 5884/ 652748 | 0.60 (0.75) | 0.90% (47) | 0.58% (56) | 1.33% (43) | 2.28 [2.04, 2.55] |
| | African American | 387/ 19600 | 0.58 (0.76) | 1.97% | 1.17% | 3.32% | 2.83 [1.86, 4.85] |
| | Latine | 1065/ 84702 | 0.59 (0.75) | 1.26% | 0.72% | 1.88% | 2.61 [2, 3.56] |
| Blood clots | European | 3875/ 757294 | 0.65 (0.74) | 0.51% (61) | 0.31% (61) | 0.95% (59) | 3.09 [2.68, 3.64] |
| | African American | 184/ 25,010 | 0.60 (0.73) | 0.74% | 0.60% | 1.32% | 2.2 [1.22, 4.56] |
| | Latine | 434/ 100130 | 0.61 (0.75) | 0.43% | 0.29% | 0.58% | 2 [1.29, 3.20] |
| Hypothyroidism | European | 5098/ 690812 | 0.68 (0.80) | 0.74% (51) | 0.31% (55) | 1.58% (49) | 5.09 [4.46, 5.91] |
| | African American | 108/ 24763 | 0.62 (0.78) | 0.44% | 0.20% | 1.13% | 5.6 [2.4, 28.03] |
| | Latine | 702/ 94415 | 0.69 (0.80) | 0.74% | 0.34% | 1.62% | 4.78 [3.38, 7.41] |
| Kidney stones | European | 6823/ 705021 | 0.67 (0.71) | 0.97% (54) | 0.48% (60) | 1.80% (49.5) | 3.78 [3.37, 4.33] |
| | African American | 168/ 24,433 | 0.58 (0.70) | 0.69% | 0.33% | 1.15% | 3.5 [1.69, 9.67] |
| | Latine | 900/ 94661 | 0.67 (0.73) | 0.95% | 0.63% | 1.79% | 2.82 [2.14, 3.82] |
| Crohn’s disease | European | 432/ 779178 | 0.70 (0.71) | 0.055% (47) | 0.028% (41) | 0.11% (42.5) | 3.73 [2.38, 6.5] |

| | | | | | | | |
|---|---|---|---|---|---|---|---|
| Colon polyps | European | 11563/ 649697 | 0.58 (0.81) | 1.78% (60) | 1.31% (62) | 2.57% (59) | 1.97 [1.81, 2.14] |
| | African American | 322/ 23297 | 0.56 (0.85) | 1.38% | 1.12% | 1.55% | 1.38 [0.82, 2.39] |
| | Latine | 1156/ 93390 | 0.61 (0.85) | 1.24% | 0.71% | 1.94% | 2.74 [2.13, 3.72] |
| Glaucoma | European | 3306/ 760327 | 0.65 (0.81) | 0.43% (66) | 0.20% (66) | 0.84% (65) | 4.10 [3.46, 4.87] |
| | African American | 168/ 24,799 | 0.61 (0.80) | 0.68% | 0.28% | 1.01% | 3.57 [1.64, 10.50] |
| | Latine | 429/ 100182 | 0.63 (0.78) | 0.43% | 0.13% | 0.79% | 6.08 [3.67, 12.67] |
| Psoriasis | European | 2022/ 742756 | 0.66 (0.67) | 0.27% (50) | 0.15% (53) | 0.42% (49) | 2.70 [2.16, 3.37] |
| | Latine | 354/ 98,283 | 0.64 (0.66) | 0.36% | 0.21% | 0.48% | 2.24 [1.33, 3.81] |
| Eczema | European | 8254/ 693320 | 0.63 (0.65) | 1.19% (51) | 0.78% (51) | 1.65% (53) | 2.12 [1.92, 2.35] |
| | African American | 313/ 22,021 | 0.58 (0.65) | 1.42% | 1.04% | 2.09% | 2 [1.26, 3.44] |
| | Latine | 1,392/ 91,534 | 0.62 (0.66) | 1.52% | 1.16% | 1.74% | 1.5 [1.17, 1.91] |

**Table S4. Lifestyle traits constituting the lifestyle risk score (LRS)**

| Phenotype name | Survey questions | Completion rate |
|---|---|---|
| wb_moderate_exercise_1 | "In a typical week, do you do the following physical activities for at least 10 minutes at a | 30.2% |

| | | |
|---|---|---|
| | time, as part of your job, your house and yard work, getting from place to place, or in your leisure time?"/"Light to moderate physical activity (not including walking) that causes a moderate increase in breathing or heart rate and light perspiration"; "How many times per week on average do you do this activity?"; "For about how long each time on average do you do this activity?" | |
| wb_strengthening_exercise_1 | "In a typical week, do you do the following physical activities for at least 10 minutes at a time, as part of your job, your house and yard work, getting from place to place, or in your leisure time?"/"Strengthening physical activity involving resistance or weights to increase muscle tone and power"; "How many times per week on average do you do this activity?"; "For about how long each time on average do you do this activity?" | 30.7% |
| wb_vigorous_exercise_1 | "In a typical week, do you do the following physical activities for at least 10 minutes at a time, as part of your job, your house and yard work, getting from place to place, or in your leisure time?"/"Vigorous physical activity that causes a large increase in breathing or heart rate and significant perspiration"; "How many times per week on average do you do this activity?"; "For about how long each time on average do you do this activity?" | 30.6% |
| wb_walking_exercise_1 | "In a typical week, do you do the following physical activities for at least 10 minutes at a time, as part of your job, your house and yard work, getting from place to place, or in your leisure time?"/"Walking (for any purpose) that causes a slight increase in breathing"; "How many times per week on average do you do this activity?"; "For about how long each time on average do you do this activity?". | 30.4% |

| | | |
|---|---|---|
| iqb.wb_more_fiber_freq | "How often do you consciously choose to eat more of the following in your meals or snacks?"/"Fiber" | 32.6% |
| iqb.wb_red_meat_freq | In a typical week, how often do you eat each of the following foods? Your best guess is fine."/"Beef, lamb, or other red meat" | 33.5% |
| iqb.wb_avoid_refined_carbs_freq | "How often do you consciously choose to eat less of the following in your meals or snacks?"/"Starch or refined carbohydrates" | 32.9% |
| iqb.wb_nonworkday_hrs_sleep | "On a typical non-workday, or weekend day if you do not work, during the past 6 weeks, how many hours of sleep did you get? Your best guess is fine." | 29.6% |
| iqb.wb_avoid_sugar_freq | "How often do you consciously choose to eat less of the following in your meals or snacks?"/"Sugar" | 33.0% |
| iqb.wb_bean_freq | "In a typical week, how often do you eat each of the following foods? Your best guess is fine."/"Beans" | 33.4% |
| iqb.wb_orange_vegetable_freq | "In a typical week, how often do you eat each of the following foods? Your best guess is fine."/"Sweet or orange vegetables" | 33.4% |
| iqb.wb_nut_freq | "In a typical week, how often do you eat each of the following foods? Your best guess is fine."/"Nuts or nut butter" | 33.4% |
| iqb.wb_avoid_cooked_food_freq | "How often do you consciously choose to eat less of the following in your meals or snacks?"/"Cooked food" | 32.5% |
| iqb.hp_red_meat | "During a typical week, how often do you eat red meat?" | 68.1% |
| iqb.wb_fruit_freq | "In a typical week, how often do you eat each of the following foods? Your best guess is fine."/"Fruit" | 33.4% |

| | | |
|---|---|---|
| iqb.wb_workday_hrs_sleep | "On a typical workday, or weekday if you do not work, during the past 6 weeks, how many hours of sleep did you get? Your best guess is fine." | 30.0% |
| iqb.wb_more_supplements_freq | "How often do you consciously choose to eat more of the following in your meals or snacks?"/"Vitamins or minerals" | 32.3% |
| iqb.hp_exercise_intake | "In a typical week how many times do you participate in any physical activities or exercise such as running, calisthenics, golf, bike riding, yoga or walking for 30 minutes or more per occurrence?" | 72.2% |
| iqb.wb_sugary_drink_freq | "In a typical week, how often do you eat each of the following foods? Your best guess is fine."/"Drinks containing sugars, including juice, energy drinks, flavored tea, non-diet soda, coffee-based drinks, and other drinks containing sugars rather than artificial sweeteners" | 33.3% |
| iqb.wb_avoid_salt_freq | "How often do you consciously choose to eat less of the following in your meals or snacks?"/"Salt" | 32.9% |
| iqb.wb_more_unsaturated_fats_freq | "How often do you consciously choose to eat more of the following in your meals or snacks?"/"Unsaturated fat or oils" | 32.2% |
| iqb.wb_avoid_meat_freq | "How often do you consciously choose to eat less of the following in your meals or snacks?"/"Meat" | 32.8% |
| iqb.wb_more_leafy_greens_freq | "How often do you consciously choose to eat more of the following in your meals or snacks?"/"Leafy green vegetables" | 32.5% |
| iqb.wb_avoid_fat_freq | "How often do you consciously choose to eat less of the following in your meals or snacks?"/"Fat" | 32.9% |

| | | |
|---|---|---|
| iqb.wb_avoid_animal_derived_food_freq | "How often do you consciously choose to eat less of the following in your meals or snacks?"/"Animal-derived food" | 32.4% |
| iqb.wb_processed_meat_freq | "In a typical week, how often do you eat each of the following foods? Your best guess is fine."/"Processed, cured, or smoked meat, including bacon, pepperoni, lunch meats, or hot dogs" | 33.6% |
| iqb.hp_smoked_100_cigarettes | "Have you smoked at least 100 cigarettes in your entire life?" | 52.6% |
| iqb.hp_alcohol_2wks | "In the last two weeks, how many servings of alcohol did you drink each day? (1 serving equals 12 oz. of beer, 5 oz. of wine, or 1.5 oz. of hard alcohol)" | 64.6% |
| iqb.wb_vitamins_minerals | "In a typical week, do you regularly take any vitamin or mineral supplements?" | 32.1% |
| iqb.wb_avoid_calories_freq | "How often do you consciously choose to eat less of the following in your meals or snacks?"/"Calories" | 33.0% |
| iqb.wb_avoid_gluten_freq | "How often do you consciously choose to eat less of the following in your meals or snacks?"/"Gluten or wheat" | 33.0% |
| iqb.wb_sweets_freq | "In a typical week, how often do you eat each of the following foods? Your best guess is fine."/"Candy, sweets, pastries, cookies, ice cream or other desserts" | 33.4% |
| iqb.wb_workday_hrs_sit | "On a typical workday, or weekday if you do not work, how many hours do you usually spend sitting?" | 30.7% |
| iqb.wb_green_vegetable_freq | "In a typical week, how often do you eat each of the following foods? Your best guess is fine."/"Green vegetables" | 33.5% |

| | | |
|---|---|---|
| iqb.wb_avoid_sat_fat_freq | "How often do you consciously choose to eat less of the following in your meals or snacks?"/"Saturated fat" | 24.8% |
| iqb.wb_fish_freq | "In a typical week, how often do you eat each of the following foods? Your best guess is fine."/"Fish" | 33.6% |
| iqb.wb_poultry_freq | "In a typical week, how often do you eat each of the following foods? Your best guess is fine."/"Chicken, turkey, or other poultry" | 33.5% |
| iqb.wb_nonworkday_hrs_sit | "On a typical non-workday, or weekend day if you do not work, how many hours do you usually spend sitting?" | 30.3% |
| iqb.wb_mindfulness_practices.yoga_tai_chi | "In a typical week, do you practice any of the following mind-body techniques for stress relief or health? Please check all that apply." : "Yoga or tai chi" | 29.4% |
| iqb.wb_pork_freq | "In a typical week, how often do you eat each of the following foods? Your best guess is fine."/"Pork" | 33.6% |

### References


1. McCarthy, S. *et al.* A reference panel of 64,976 haplotypes for genotype imputation. *Nat. Genet.* **48**, 1279–1283 (2016).
2. Durand, E. Y., Do, C. B., Mountain, J. L. & Michael Macpherson, J. Ancestry Composition: A Novel, Efficient Pipeline for Ancestry Deconvolution. *bioRxiv* 010512 (2014) doi:10.1101/010512.
3. Henn, B. M. *et al.* Cryptic distant relatives are common in both isolated and cosmopolitan genetic samples. *PLoS One* **7**, e34267 (2012).
4. Purcell, S. *et al.* PLINK: A Tool Set for Whole-Genome Association and Population-Based Linkage Analyses. *The American Journal of Human Genetics* vol. 81 559–575 Preprint at https://doi.org/10.1086/519795 (2007).
5. Privé, F., Vilhjálmsson, B. J., Aschard, H. & Blum, M. G. B. Making the Most of Clumping and Thresholding for Polygenic Scores. *Am. J. Hum. Genet.* **105**, 1213–1221 (2019).
6. Chen, T. & Guestrin, C. XGBoost. *Proceedings of the 22nd ACM SIGKDD International Conference on Knowledge Discovery and Data Mining* Preprint at https://doi.org/10.1145/2939672.2939785 (2016).
